# Integrating *de novo* and inherited variants in over 42,607 autism cases identifies mutations in new moderate risk genes

**DOI:** 10.1101/2021.10.08.21264256

**Authors:** Xueya Zhou, Pamela Feliciano, Tianyun Wang, Irina Astrovskaya, Chang Shu, Jacob B. Hall, Joseph U. Obiajulu, Jessica Wright, Shwetha Murali, Simon Xuming Xu, Leo Brueggeman, Taylor R. Thomas, Olena Marchenko, Christopher Fleisch, Sarah D. Barns, LeeAnne Green Snyder, Bing Han, Timothy S. Chang, Tychele N. Turner, William Harvey, Andrew Nishida, Brian J. O’Roak, Daniel H. Geschwind, The SPARK Consortium, Jacob J. Michaelson, Natalia Volfovsky, Evan E. Eichler, Yufeng Shen, Wendy K. Chung

## Abstract

Despite the known heritable nature of autism spectrum disorder (ASD), studies have primarily identified risk genes with *de novo* variants (DNVs). To capture the full spectrum of ASD genetic risk, we performed a two-stage analysis of rare *de novo* and inherited coding variants in 42,607 ASD cases, including 35,130 new cases recruited online by SPARK. In the first stage, we analyzed 19,843 cases with one or both biological parents and found that known ASD or neurodevelopmental disorder (NDD) risk genes explain nearly 70% of the genetic burden conferred by DNVs. In contrast, less than 20% of genetic risk conferred by rare inherited loss-of-function (LoF) variants are explained by known ASD/NDD genes. We selected 404 genes based on the first stage of analysis and performed a meta-analysis with an additional 22,764 cases and 236,000 population controls. We identified 60 genes with exome-wide significance (p < 2.5e-6), including five new risk genes (*NAV3, ITSN1, MARK2, SCAF1*, and *HNRNPUL2)*. The association of *NAV3* with ASD risk is entirely driven by rare inherited LoFs variants, with an average relative risk of 4, consistent with moderate effect. ASD individuals with LoF variants in the four moderate risk genes *(NAV3, ITSN1, SCAF1, and HNRNPUL2*, n = 95) have less cognitive impairment compared to 129 ASD individuals with LoF variants in well-established, highly penetrant ASD risk genes (*CHD8, SCN2A, ADNP, FOXP1, SHANK3)* (59% vs. 88%, p= 1.9e-06). These findings will guide future gene discovery efforts and suggest that much larger numbers of ASD cases and controls are needed to identify additional genes that confer moderate risk of ASD through rare, inherited variants.

## Introduction

Autism spectrum disorder (ASD) is a neurodevelopmental condition characterized by impaired social communication and repetitive behaviors^1^. Previous studies in ASD utilized family-based designs to focus on *de novo* variants (DNVs) identified from parent-offspring trios^2-8^. Over one-hundred high confidence ASD genes enriched with likely deleterious DNVs have been identified^8^, most of which are also enriched for DNVs in other neurodevelopment disorders (NDDs)^9-11^. Statistical modeling suggests there are ∼1000 genes with DNV variants in ASD^12,13^. However, despite the large effect size of individual pathogenic DNVs, all DNVs together only explain ∼ 2% of variance in liability for ASD^8,14^.

On the other hand, ASD is highly heritable (estimated heritability over 0.5)^14-16^. Previous studies estimated that common variants explain up to half of the heritability^14^, although only five genome-wide significant loci have been identified^17^. The role of inherited coding variants has been evaluated using familial segregation of loss-of-function (LoF) variants (stop-gain, splice site and frameshift variants) carried by parents without ASD diagnoses or intellectual disability. Rare LoF variants only in genes intolerant of variation^9,18^ are over-transmitted to probands compared with siblings without ASD^7,8,19-22^.However, identification of the individual risk genes enriched by such inherited variants has remained elusive.

We have created a large longitudinal research cohort, SPARK (SPARKForAutism.org^23^) to advance research on the genetic, behavioral, and clinical features associated with ASD. SPARK represents the largest ASD cohort in the world, with over 100,000 individuals with ASD enrolled.

Rare, LoF variants are enriched in developmental disorders including ASD^22,24^, but LoF variants in the general population are also enriched for sequencing and annotation artefacts^25^, which present technical challenges in large sequencing studies. Methods to distinguish between high and low confidence LoF variants^18,26,27^ have been used to quantify gene level LoF intolerance^18,26,28,29^ and to refine the role of *de novo* LoF variants in NDDs^20^.

Here we present an integrated analysis of *de novo* and inherited coding variants in over 42,607 ASD cases, including cases from previously published ASD cohorts and 35,130 new cases from SPARK. To our knowledge, this analysis is the largest sequencing study of ASD to date. In our two-stage design, we first characterized the contribution of DNVs and rare inherited LoF variants to ASD risk. Results from the first stage informed the second stage, in which we conducted a meta-analysis of 404 genes. By combining evidence from DNVs, over-transmission, and case-control comparison, we identified 60 ASD risk genes with exome-wide significance, including five new genes not previously implicated in neurodevelopmental conditions. Finally, we estimated the effect sizes of known and newly significant genes and used them for power calculations to inform the design of future studies.

## Results

### Overview of data and workflow

We aggregated exome or whole genome sequencing (WGS) data of 35,130 new cases from the SPARK study and 7,665 cases from published ASD studies (ASC^3,8^, MSSNG^6^, and SSC^2,30^) (**Supplementary Table S1**) and performed a two-stage analysis (**Figure 1)**. In the first stage, we analyzed *de novo* coding variants (DNVs) in 16,877 ASD trios and assessed transmission of rare LoF variants in 20,491 parents without ASD diagnoses or intellectual disability to offspring with ASD (including 9,504 trios and 2,966 single-parent-proband duos). For DNVs, we characterized the enrichment pattern in known and candidate risk genes, mutation intolerance (ExAC pLI^18^ and gnomAD metrics^26^) and performed gene-based burden tests of *de novo* LoF and missense variants by DeNovoWest^11^. For rare inherited LoFs, we estimated the over-transmission from parents without an ASD diagnosis to ASD cases in all genes and gene sets predefined by functional genomic data or results from DNV analysis. Based on DNV enrichment and over-transmission patterns in gene sets, we selected 404 genes for meta-analysis in stage 2 utilizing 22,764 new cases with exome or WGS data. In stage 2, we applied DeNovoWEST on DNVs, conducted transmission-disequilibrium tests on inherited LoFs in trios or duos, performed burden tests on rare LoFs in cases compared with population controls (104,068 subjects from gnomAD exome, non-neuro subset v2.1.1 and 132,345 TOPMed subjects), and combined the p-values to estimate a final p-value for each of the 404 genes. Finally, we performed a mega-analysis of rare LoFs in all cases and controls to estimate the effect sizes of known or new candidate ASD genes to inform future studies.

**Figure 1.**
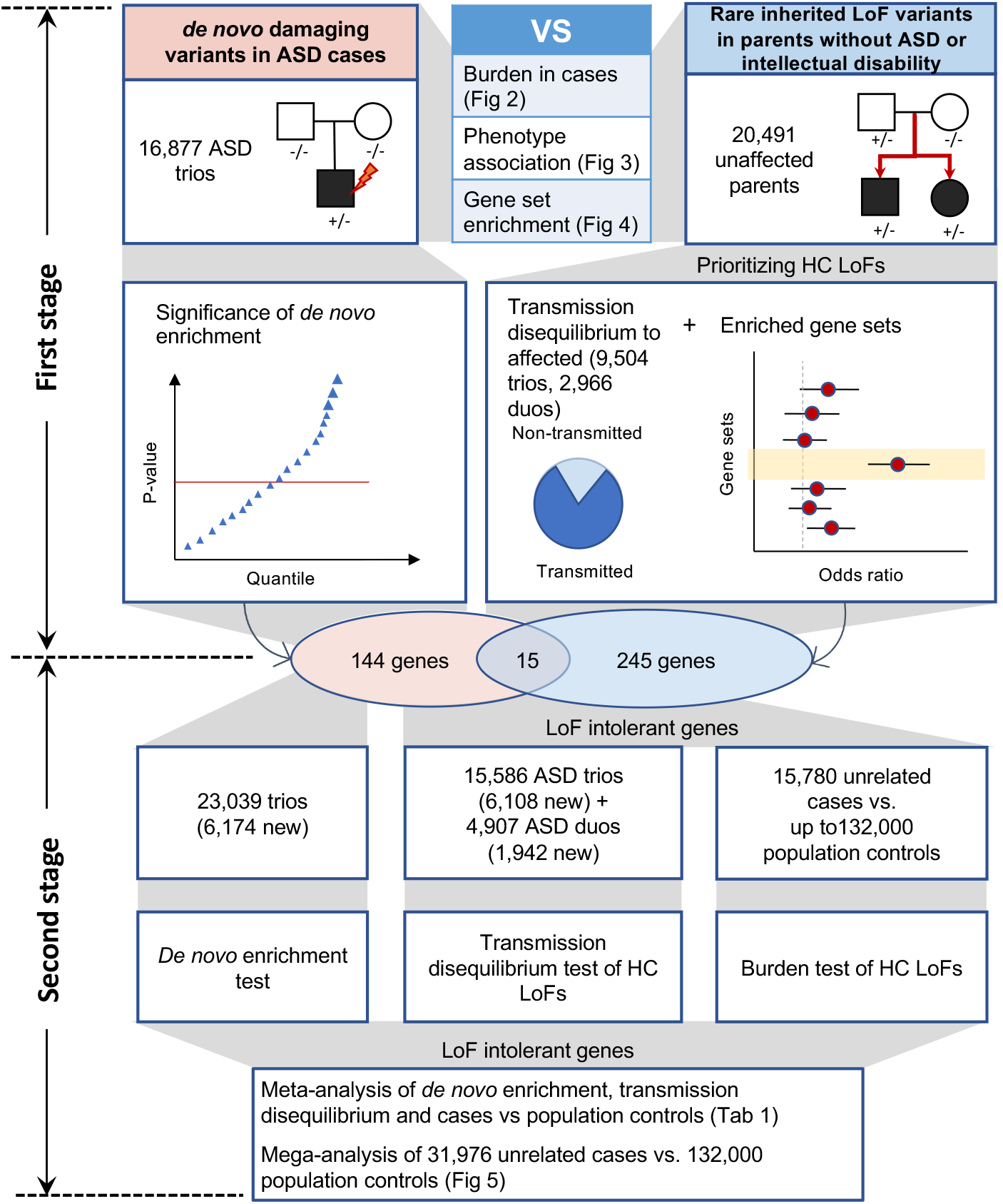
Analysis workflow. In the discovery stage, we identified *de novo* variants in 16,877 ASD trios and rare LoF variants in 20,491 parents without ASD diagnoses and intellectual disability. We compared properties of *de novo* and rare variants to identify rare LoFs that contribute to genetic risk in individuals with ASD. We also evaluated their associations with cognitive impairment and enriched gene sets. We performed an initial exome-wide scan of genes enriched by *de novo* variants or showing transmission disequilibrium (TD) of rare LoFs to affected offspring and selected a total of 404 genes for further replication, including 159 *de novo* enriched genes and 260 prioritized TD genes from enriched gene sets (15 genes were in both). In the meta-analysis stage, we first evaluated evidence from *de novo* enrichment and TD of rare, inherited LoFs in an expanded set of family-based samples including over 6,000 additional ASD trios and around 2000 additional duos. The *de novo* variants in ASD were combined with those from additional 31,565 NDD trios to refine the filters of high confidence (HC) LoFs in *de novo* LoF enriched genes. We also constructed an independent dataset of LoF variants of unknown inheritance from 15,780 cases that were not used in *de novo* or transmission analysis. We compared LoF rates in cases with two population-based sets of controls (n ∼104,000 and ∼132,000, respectively). For 367 LoF intolerant genes on autosomes, the final gene level evidence was obtained by meta-analyzing p-values of *de novo* enrichment, TD of HC rare, inherited LoFs, and comparison of HC LoFs from cases and controls not used in the de novo or transmission analysis. We also performed a mega-analysis that analyzed HC LoFs identified in all 31,976 unrelated ASD cases and compared their rates with population-based controls.

### Known ASD or NDD risk genes explain two-thirds of population attributable risk of de novo coding variants in ASD

In the first stage, we combined data from four large-scale ASD cohorts, resulting in 16,877 unique ASD trios and 5,764 unaffected trios (**Supplementary Table S1**). The four cohorts show similar exome-wide burden of DNVs in simplex families. The burden of *de novo* LoF variants in cases with a family history of ASD is significantly lower than those without a reported family history (p=1.1e-4 by Poisson test), whereas the burden of predicted *de novo* damaging missense (D-mis, defined by REVEL score^31^>=0.5) and synonymous variants are similar (**Supplementary Figure S1**).

Compared to unaffected offspring, the excess of damaging DNVs (*de novo* LoF and D-mis variants) in individuals with ASD is concentrated in LoF-intolerant genes, defined as genes with a probability of being LoF intolerant (pLI)^18^ >=0.5 in the Exome Aggregation Consortium (ExAC). Using LoF observed/expected upper-bound fraction (LOEUF), a recently developed gene constraint metric^26^, the burden of damaging DNVs is highest among genes ranked in the top 20% of LOEUF scores (**Figure 2A**). Overall, the population attributable risk (PAR) from damaging DNVs is about 10%. We assembled 618 previously established dominant (“known”) ASD or NDD risk genes (**Supplementary Table S2**). These genes explained about 2/3 of the PAR from damaging DNVs. Excluding these genes, the fold enrichment of damaging DNVs was greatly attenuated (**Figure 2A**).

**Figure 2.**
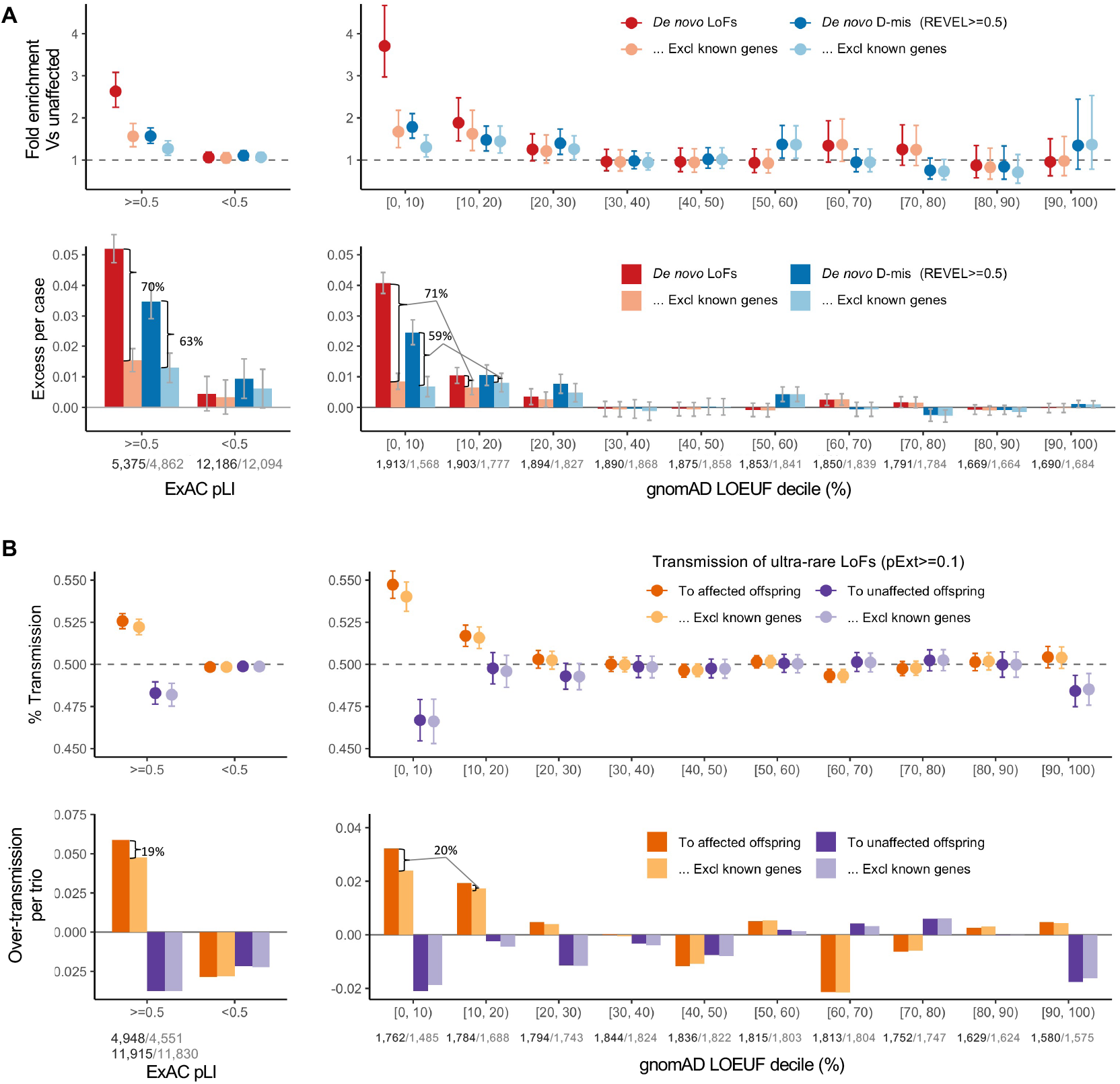
Comparison of burden between *de novo* damaging variants and rare, inherited LoFs in ASD. (A) The burden of *de novo* variants was evaluated by the rate ratio and rate difference between 16,877 ASD and 5,764 unaffected trios. The exome-wide burden of *de novo* LoF and Dmis (REVEL>=0.5) variants are concentrated in constrained genes (ExAC pLI>=0.5) and in genes with the highest levels of LoF-intolerance in the population— defined by the top two deciles of gnomAD LOEUF scores. Burden analysis was repeated after removing known ASD/NDD genes. The number of genes before and after removing known genes in each constraint bin is shown below the axis label. Among constrained genes (ExAC pLI>=0.5 or the top 20% of gnomAD LOEUF scores), close to two thirds of case-control rate differences of *de novo* LoF and Dmis variants can be explained by known genes. (B) The burden of inherited LoFs was evaluated by looking at the proportion of rare LoFs in 20,491 parents without ASD diagnoses or intellectual disability that are transmitted to affected offspring in 9,504 trios and 2,966 duos and show evidence of over-transmission of LoFs per ASD trio. As a comparison, we also show the transmission disequilibrium pattern to unaffected offspring in 5,110 trios and 129 duos. Using ultra-rare LoFs with pExt>=0.1, exome-wide signals of transmission disequilibrium of rare, inherited LoF variants also concentrate in constrained genes (ExAC pLI>=0.5) and in genes within the top two deciles of gnomAD LOEUF scores. Analysis was restricted to autosomal genes and repeated after removing known ASD/NDD genes (number of genes in each constrained bin before and after removing known genes is shown below the axis label). Among all constrained genes, only one-fifth of over-transmission of LoFs to ASD trios can be explained by known ASD/NDD genes.

To assess the evidence of DNVs in individual genes, we applied DeNovoWEST^11^, which integrates DNV enrichment with clustering of missense variants in each gene. The initial DeNovoWEST scan of DNVs in 16,877 ASD trios identified 159 genes with p<0.001 (**Supplementary Table S3**).

### Rare inherited LoF variants contribute to ASD risk mostly through unknown risk genes

To analyze the contribution of rare inherited LoF variants to ASD risk, we evaluated transmission disequilibrium in ultra-rare (allele frequency < 1e-5) high-confidence (by LOFTEE^26^ and pext^27^; see Methods and Supplementary Note) LoF variants from parents without ASD diagnoses or intellectual disability to affected offspring with ASD in 9,504 trios and 2,966 duos from the first stage (**Supplementary Table S4**). For a given set of genes, we quantified transmission disequilibrium using the number of over-transmitted (excess in transmission over non-transmission) LoF variants per trio; parent-offspring duos were considered half-trios.

Among autosomal genes, the overall transmission disequilibrium signal of ultra-rare LoF variants is enriched in LoF intolerant genes (ExAC pLI>=0.5) and in genes within the top 20% of LOEUF scores (**Figure 2B**), similar to the burden of damaging DNVs. We observed both over-transmission to affected and under-transmission to unaffected offspring, especially in genes within the top 10% of LOEUF scores. However, known ASD/NDD genes only explain ∼20% of over-transmission of LoF variants to affected offspring (**Figure 2B**). On the X chromosome, we only considered transmission from mothers without ASD diagnoses to 9,883 affected sons and 2,571 affected daughters (**Supplementary Table S4**). Rare LoF variants in mothers without ASD diagnoses only show significant over-transmission to affected sons but not affected daughters and remain significant after removing known ASD/NDD genes (**Supplementary Figure S2**).

Together, these data suggest that most genes conferring inherited ASD risk are yet to be identified. Autosomal rare D-mis variants also show evidence of transmission disequilibrium to affected offspring, although the signal is much weaker and dependent on gene set, D-mis prediction method, pExt and allele frequency filters (**Supplementary Figure S3**).

To characterize the properties of genes contributing to ASD risk through rare inherited variants, we defined 25 gene sets from five categories representing both functional and genetic evidence relevant to ASD (**Supplementary Table S5 and Supplementary Figure S4**). We limited the genes to 5,754 autosomal constrained genes (ExAC pLI>=0.5 or top 20% of LOEUF scores) and performed TDT (**Supplementary Table S6**). For each gene set, we tested if high-confidence rare LoF variants show a higher frequency of transmission to ASD offspring than the remaining genes in the overall constrained gene set. As a comparison with DNVs, we also tested if the same set of genes are more frequently disrupted by damaging DNVs than the rest of the genes in ASD trios using the framework of dnEnrich^32^.

We first considered functional gene sets derived from the neuronal transcriptome, proteome, or regulome. We confirmed significant enrichment in damaging DNVs (p<0.005 by simulation) in the gene sets that were previously suggested to be enriched for ASD risk genes including expression module M2/3^33^, RBFOX1/3 targets^34^, FMRP targets^35^, and CHD8 targets^36^. However, this enrichment can be largely explained by known ASD/NDD genes (**Supplementary Figure S5**). For ultra-rare inherited LoF variants, we found the proportion of transmission to ASD individuals in most functional gene sets is close to all genes in the background; only RBFOX targets show a weak enrichment but can be largely explained by known genes (**Figure 3**). We also applied two recently developed machine learning methods to prioritize ASD risk genes: forecASD^37^ that integrates brain expression, gene network, and other gene level metrics, and A-risk^38^ that uses cell-type specific expression signatures in developing brain. Although enrichment of DNVs in genes predicted by these methods are mainly explained by known genes, genes prioritized by A-risk are significantly enriched with inherited LoFs that cannot be explained by known genes. Using A-risk>=0.4 (recommended threshold), 30% of constrained genes (n=1,464) were prioritized and explain 64% of the over-transmission of LoF variants to ASD offspring (p=2.6e-5 by chi-squared test). The enrichment is even higher than genes prioritized by the LOEUF score: 33% of genes (N=1,777) in the top decile of LOEUF account for 55% over-transmission (P=3.5e-4 by chi-squared test) (**Figure 3**).

**Figure 3.**
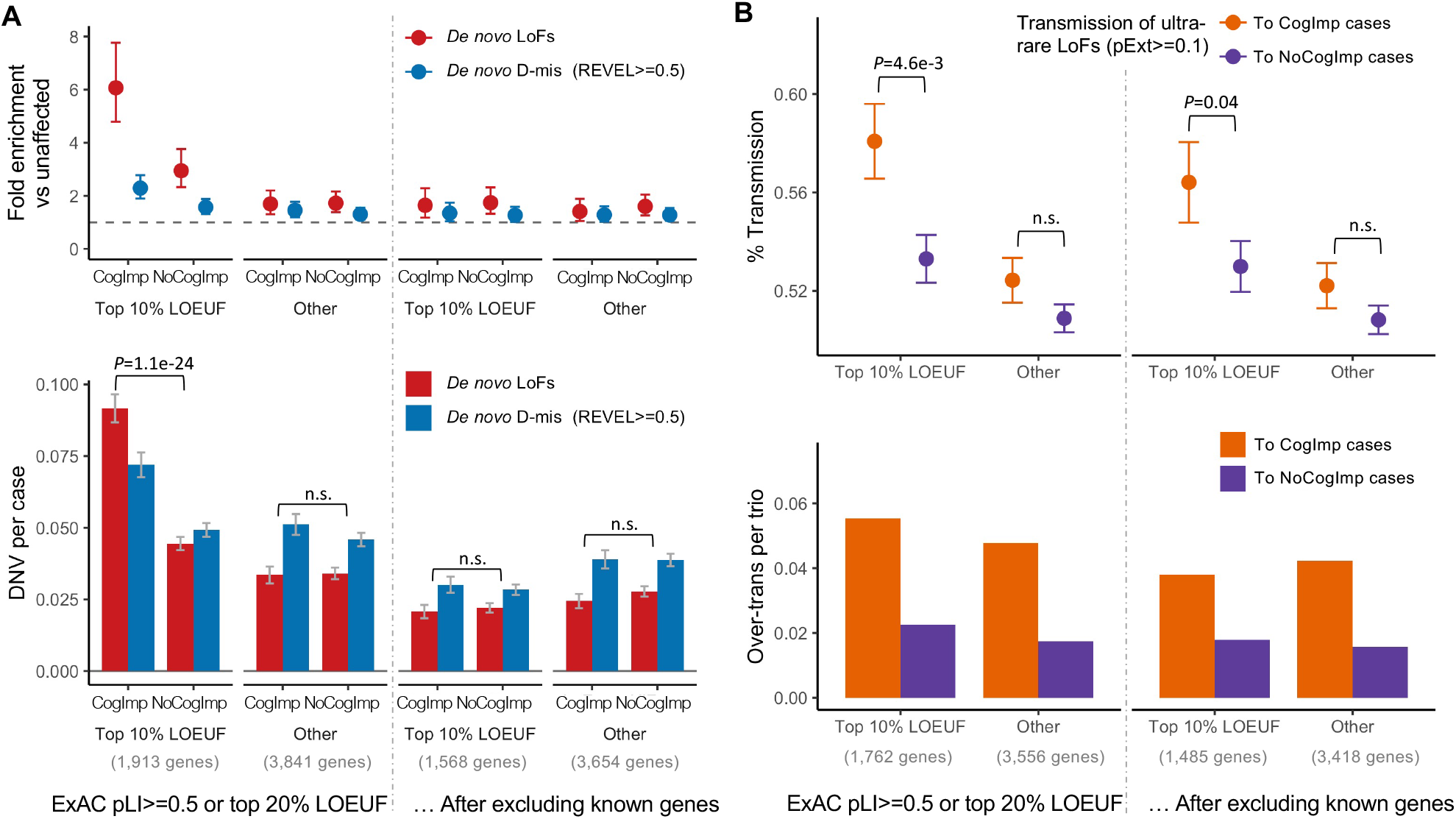
Association of rare, inherited LoFs with cognitive impairment in ASD cases. Ultra-rare inherited LoFs with pExt>=0.1 in genes with the top 10% gnomAD LOEUF scores also show a higher proportion of transmission and a higher over-transmission rate to ASD offspring with cognitive impairment than those without. Rare LoFs in other constrained genes are not significantly associated with phenotypic severity. The increased burden of inherited LoFs in cases with cognitive impairment remains significant after removing known ASD/NDD genes.

We also considered gene sets that have evidence of genetic association with DNVs. Genes nominally enriched by DNVs (P<0.01 by DeNovoWEST; N=300) in ASD from the current study have a significantly higher over-transmission rate than other constrained genes (Odds ratio=1.39, p=3.0e-5 by chi-squared test) (**Figure 3**), although these genes only account for 21% of the over-transmission. Genes nominally enriched by DNVs in other NDDs^11^ are also significantly enriched by DNVs in ASD and weakly enriched by inherited LoFs in ASD; however, both can be largely explained by known genes (**Figure 3**). This suggests that a subset of ASD genes increase risk by both *de novo* and inherited variants, and new genes can be identified by integrating evidence from DNV enrichment and TDT.

### DNVs and a subset of rare inherited LoFs are associated with cognitive impairment

To evaluate the association of genotypes with phenotype in ASD, we used self-reported cognitive impairment in SPARK, a Vineland score of <70 in the SSC or the presence of intellectual disability in ASC. Damaging DNVs in genes ranked within the top 10% of LOEUF scores show a higher burden (p=1.1e-24, by chi-squared test) in ASD cases with evidence of cognitive impairment than other cases, consistent with previous results^2,8^ (**Figure 4A**). Once known ASD/NDD genes were excluded, the residual burden of damaging DNVs in genes at the top 10% LOEUF is greatly reduced and not significantly associated with cognitive phenotype in ASD (**Figure 4A**). Over-transmission of rare LOFs in genes within the top 10% of LOEUF genes to ASD cases with cognitive impairment is about 2.7 times higher than to the cases without cognitive impairment (p=4.6e-3 by chi-squared test) and is still 2x higher (p=0.04 by chi-squared test) once known ASD/NDD genes were excluded (**Figure 4B**). However, rare LoFs in genes prioritized by A-risk, in which there is significant over-transmission to all cases overall, are not associated with cognitive impairment (**Supplementary Figure S6**). Taken together, these results suggest that rare variants in the top 10% of LOEUF genes—most of which are already known to be ASD/NDD risk genes—are associated with cognitive impairment. However, a subset of rare, inherited variants, particularly those prioritized by A-risk, are not associated with cognitive impairment.

**Figure 4.**
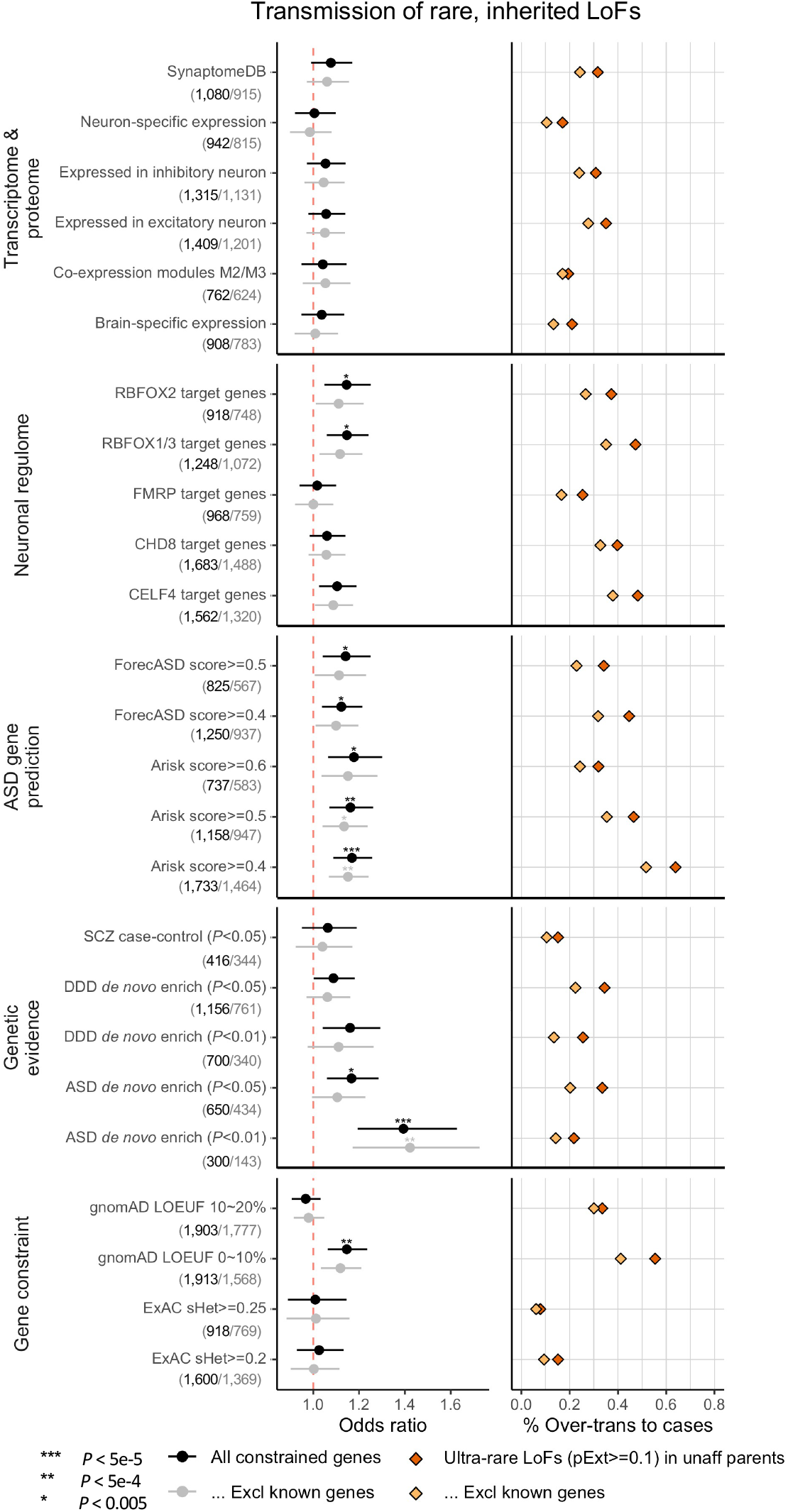
Enrichment of rare LoF variants in ASD cases across gene sets. Gene sets were defined and grouped by transcriptome proteome, neuronal regulome, ASD gene prediction scores, genetic evidence from neuropsychiatric diseases, and gene level constraint. Analyses were repeated after removing known ASD/NDD genes. (Number of genes in each set before and after removing known genes are shown in bracket below gene set.) Dots represent fold enrichment of DNVs or odds ratios for over-transmission of LoFs in each set. Horizontal bars indicate the 95% confidence interval. For each gene set, we show the percentage of over-transmission of rare LoFs to cases. Enrichment of rare, inherited LoFs was evaluated by comparing the transmission and non-transmission of ultra-rare LoFs with pExt>=0.1 in the gene set versus those in all other constrained genes using a 2-by-2 table. *P*-values were given using the chi-squared test.

### Meta-analysis of de novo and rare inherited LoF variants identifies 5 new risk genes with exome-wide significance

Based on results from the first stage of analysis, 404 genes showed plausible evidence of contributing to ASD risk, including: 1) 260 genes with evidence of TDT (TDT statistic^39^>=1) and in gene sets enriched with rare inherited LoFs (top 10% LOEUF or within top 20% LOEUF and A-risk>=0.4) (**Supplementary Table S6**) and 2) 159 genes with p<0.001 from the DeNovoWEST analysis of DNVs (with 15 genes by both) (**Supplementary Table S3**). We performed a meta-analysis on the 367 autosomal genes with all data from Stage 1 and Stage 2, which includes 6,174 new ASD trios, 1,942 new duos, 15,780 unrelated cases (see Methods), and 236,000 population controls.

In the meta-analysis, we used Fisher’s method^40^ to combine 3 p-values that estimate independent evidence of DNVs, TDT, and case-control comparison: (1) DeNovoWEST with DNVs from both Stage 1 and 2 (n=23,039 trios, **Supplementary Table S1**) using the parameters estimated in Stage 1, (2) TDT with rare LoF variants in parents without ASD diagnoses or intellectual disability with affected offspring in 15,586 trios and 4,907 duos (**Supplementary Table S4**), and (3) unrelated cases (**Supplementary Table S7**) compared to population controls using a binomial test. We used two sets of controls: gnomAD exome v2.1.1 non-neuro subset (n=104,068) and TOPMed WGS (freeze 8, n=132,345). We performed a case-control burden test using the two sets separately and input the larger p-value for the Fisher’s method. This approach avoids any sample overlap and provides sensitivity analysis to ensure that significant genes are not dependent on the choice of population reference. Although population reference data were processed by different bioinformatics pipelines, the cumulative allele frequencies (CAFs) of high-confidence (HC, see Methods) LoF variants are similar between internal pseudo-controls (see Methods) and the two population references after applying the same LoF filters (**Supplementary Figure S7**). Previous population genetic simulations predict that for genes under moderate to strong selection (selection coefficient>0.001), deleterious variants are expected to arise within 1,000 generations and population demographic histories do not confound the CAFs of deleterious alleles in these genes^41^. For 367 selected autosomal genes, the point estimates of selection coefficient under mutation-selection balance model^42^ are all greater than 0.01 (**Supplementary Figure S8**). Consistent with the theoretical predictions, most HC LoF variants in these genes are ultra-rare (**Supplementary Figure S9**) and the CAFs of HC LoF variants in European and non-European population samples are highly correlated (**Supplementary Figure S10**). Thus, we included population samples across all ancestries as controls. To make use of all genetic data collected, we also included rare variants of unknown inheritance from autism cases that were analyzed in the first stage. These variants come from cases that are part of parent-autism duos; such variants were either inherited from the parent not participating in the study or occurred *de novo*. Therefore, these data represent data independent of the transmission disequilibrium testing, even though the same cases were included in TDT.

We identified 60 genes with exome-wide significance (p<2.5e-6). Figure 5 summarizes the distribution of LoF variants (with different modes of inheritance) in genes that reached experimental-wide significance by DNV enrichment (**Figure 5A**) and other significant genes by meta-analysis (**Figure 5B, Supplementary Figure S11**). Genes that are significant only in meta-analysis tend to harbor more inherited LoF variants than *de novo* variants, consistent with their lower penetrance for ASD or NDD.

**Figure 5.**
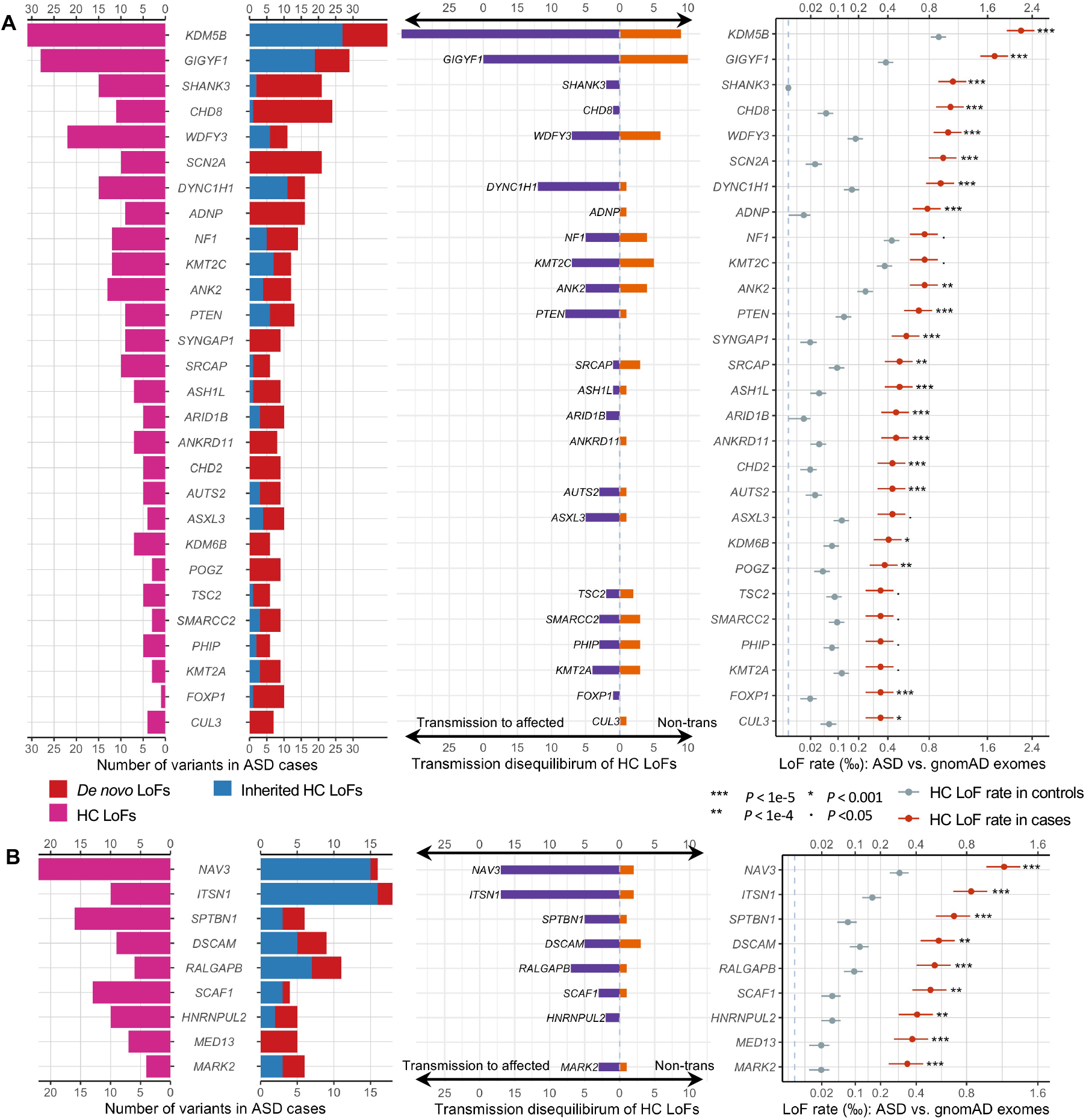
Distribution of *de novo* and inherited LoF variants in known and novel ASD genes in cases and population controls. From left to right: pyramid plots summarizing the number of *de novo* LoF variants in 15,857 ASD trios, inherited HC LoFs in 18,720 unrelated offspring included in transmission analysis, and HC LoFs in 15,780 unrelated cases; bar plot of transmission vs. non-transmission for rare HC LoFs identified in parents without ASD diagnoses or intellectual disability; three plots comparing the HC LoF rate in 31,976 unrelated ASD cases with gnomAD exomes (non-neuro subset, 104,068 individuals). Horizontal bars indicate standard errors. (A) The upper panel shows 28 known ASD/NDD genes in which LOEUF scores are in the top 30% of gnomAD, have a p-value for enrichment among all DNVs (p <9e-6) in 23,039 ASD trios, and have more than 10 LoFs. (B) The lower panel shows 9 additional ASD risk genes that achieved a p-value of <9e-6 in Stage 2 of this analysis. The majority of genes in the lower panels harbor more inherited LoFs than *de novo* variants. All five novel genes (**Error! Reference source not found**.) are shown in the lower panel. Note that the x-axes of LoF rates are in the squared root scale.

Although most significant genes were previously known, we identified five new genes that are exome-wide significant regardless of the choice of population reference: *NAV3, MARK2, ITSN1, SCAF1*, and *HNRNPUL2* (**Table 1**). As expected, most supporting variants are ultra-rare, and results are robust to the allele frequency filter. These five new genes together explain 0.27% population attributable risk ratio (PAR) (**Supplementary Table S8**). *NAV3* has a similar PAR as *CHD8* and *SCN2A* (∼0.095%). *ITSN1* is similar to *PTEN* (∼0.065%).

**Table 1:**
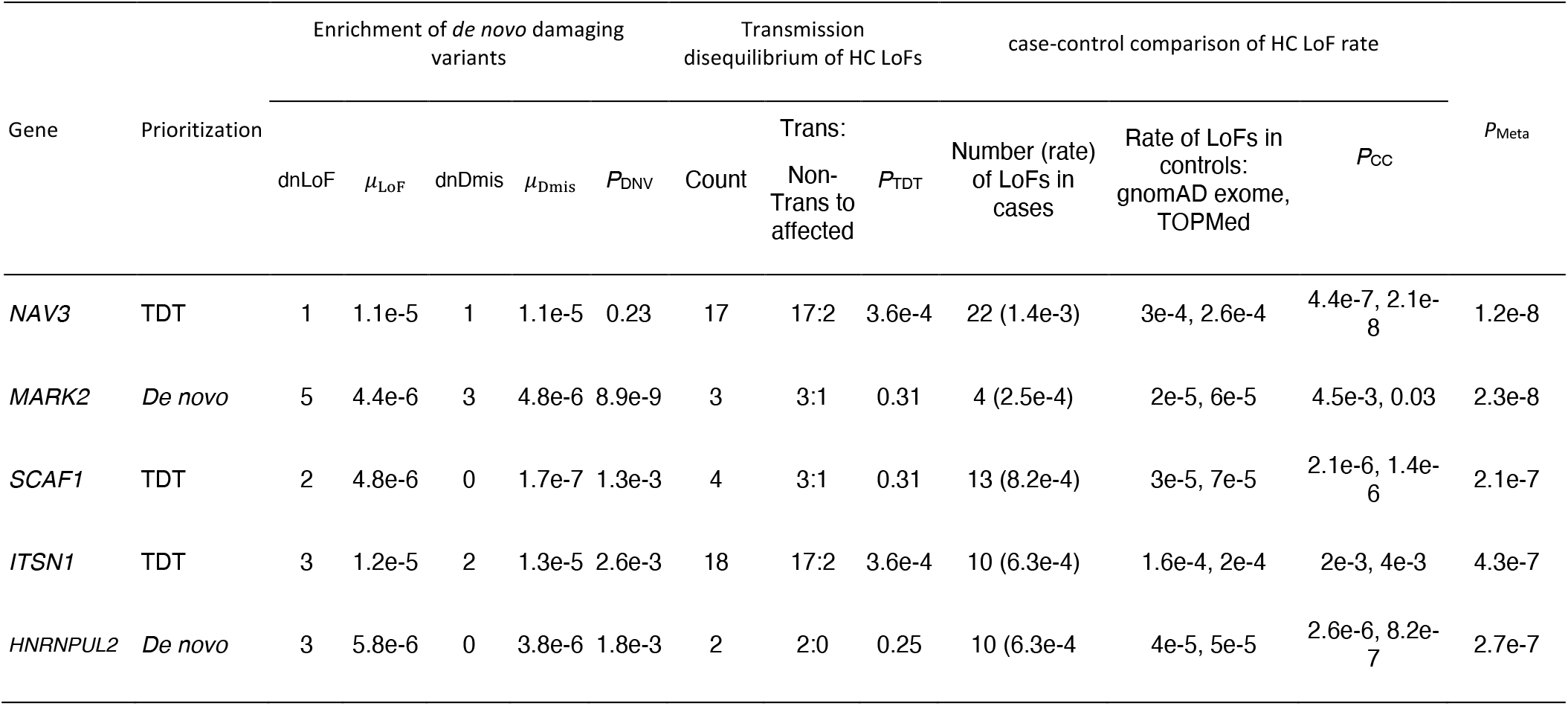
Statistical evidence for the five novel exome-wide significant ASD risk genes identified in this study. Control HC LoF rates are estimated from two population-based reference panels: gnomAD exome (v2.1.1, non-neuro subset, 104,068 individuals), and TopMed (freeze 8, 132,345 individuals). Meta-analysis is done by combining p-values from *de novo*, TDT and pseudo case-control analysis using Fisher’s method. For pseudo case-control, we conservatively took the largest p-value for meta-analysis. *P*_DNV_: One-sided p-value for enrichment of all DNVs in 23,053 ASD trios, *P*_TDT_: One-sided p-value of over-transmission of HC LoFs to affected offspring in 28,556 trios and 4,526 duos, *P*_CC_: One-side p-value for increased HC LoF rate in 15,811 unrelated cases compared with population controls (showing two p-values from comparison with gnomAD exome and TOPMed data respectively).

The association of *NAV3* with ASD risk is entirely driven by rare inherited variants (**Table 1**). *NAV3* harbors a single HC *de novo* LoF variant in an unaffected sibling in the SSC and was previously included in the negative training set by A-risk^38^. Despite this, NAV3 still has a high A-risk score, suggesting NAV3’s expression pattern is highly similar to known ASD genes (**Supplementary Data 1**)^7,43^. *NAV3* has high expression in inner cortical plate of developing cortex ^33^, and in pyramidal neurons (hippocampus CA1 and somatosensory cortex) and cortical interneurons, consistent with the signatures of known ASD genes ^44^ (**Supplementary Figure S12**).

The association of *MARK2* with ASD risk is primarily driven by DNVs. *MARK2* is also associated with other NDDs^11^ (P=2.7e-5 by DeNovoWEST) including Tourette syndrome^45^ and epilepsy^46^. We find that 3/8 of autistic offspring with variants in *MARK2* report epilepsy, 2/8 report Tourette syndrome and 7/8 have evidence of cognitive impairment (**Supplementary Table S9**).

The remaining three novel genes have support from both DNVs and rare LoFs. Two genes have suggestive evidence from other NDD studies. *ITSN1* and *SCAF1* shows nominal significance of DNV enrichment in 31,058 NDD trios^11^ (P<0.05 by DeNovoWEST). *SCAF1* was among the top 50 genes from gene-based burden test in a recent schizophrenia case-control study (P=0.0027 by burden test)^47^. Both *ITSN1* and *NAV3* have moderate effect sizes (point estimate of relative risk 3∼6, **Supplementary Table S8**). *ITSN1* has been highlighted in our previous study with evidence of enriched inherited LoFs^7^. *ITSN1* and *NAV3* also show increased CAF of LoF variants in a recent study by ASC^8^ although the association was not significant. We also assessed deletions in these new genes. For both *ITSN1* and *NAV3*, we identified four partial or whole gene deletions in 33,083 parents without ASD diagnoses or intellectual disability that also show transmission disequilibrium to affected offspring (**Supplementary Figure S13**).

While both *de novo* and rare inherited LoFs in the most constrained genes are strongly associated with intellectual disability (ID) in ASD (**Figure 4**), the association of such variants in individual genes is heterogenous, as suggested by the lack of association of rare inherited variants in genes with high A-risk (**Supplementary Figure S5**). We calculated the burden of cognitive impairment (see **Methods**) in 87 ASD individuals with HC LoF variants in the four novel moderate risk genes and compared it to 129 individuals with HC LoF in the well-established ASD risk genes *CHD8, SCN2A, SHANK3, ADNP* and *FOXP1* as well as 8,731 individuals with ASD in SPARK (**Supplementary Figure S14**). Although most individuals with variants in well-established ASD risk genes have some evidence of cognitive impairment (88%,) individuals with LoF variants in the moderate risk genes had significantly lower burden (56%, p=4.5e-7 by chi-squared test). Individuals with HC LOFs in the moderate risk genes did not have a significantly different burden of cognitive impairment than 8,731 individuals with ASD in SPARK (56% vs. 50%, p = n.s.). Individuals with LoF variants in the moderate risk genes also had a similar male: female (4:1) ratio compared to the larger cohort whereas individuals with variants in the well-established ASD risk genes showed significantly less male bias (1.6: 1, p= 0.009 by chi-squared test) **(Supplementary Figure S14)**, as previously reported^2^. We also predicted full-scale IQ on all participants based on parent-reported data using a machine learning method^48^. Carriers of rare LoFs in three (*NAV3, SCAF1*, and *HNRNPUL2*) of the four new genes with substantial contribution from rare inherited variants have similar IQ distribution as the overall SPARK cohort (**Figure 6A**), which is substantially higher than heterozygotes with rare LoFs in well-established, highly-penetrant genes that contribute to ASD primarily through *de novo* variants (“DN genes”), such as *CHD8, SHANK3*, and *SCN2A*. In fact, both novel and established genes with significant contribution from rare inherited LoFs are less associated with ID than DN genes (**Figure 6B**). Across these genes, there is a significant negative correlation (r=0.78, p=0.001) of estimated relative risk of rare LoFs with average predicted IQ of the individuals with these variants (**Figure 6C**). These genes could be associated with other neurobehavioral phenotypes.

**Figure 6.**
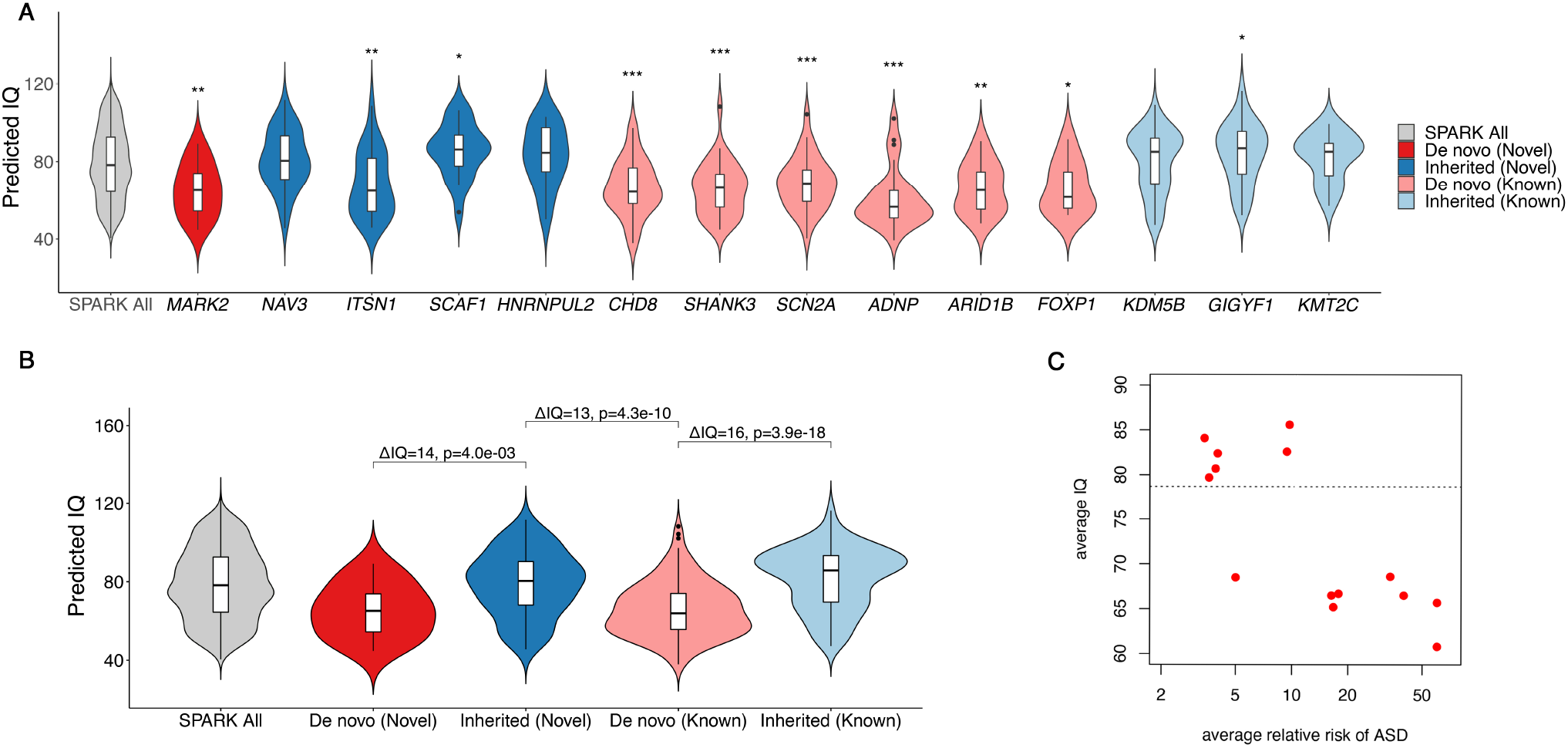
Predicted full-scale IQ (FSIQ) in individuals with pathogenic variants in inherited or *de novo* genes in SPARK. We examined the distribution of predicted IQ by a machine learning method^48^ for individuals with ASD with a LoF mutation in one of the five novel exome-wide significant genes (*MARK2, NAV3, ITSN1, SCAF1, HNRNPUL2*) and nine known ASD genes (*CHD8, SHANK3, SCN2A, ADNP, ARID1B, FOXP1, KDM5B, GIGYF1, KMT2C*), compared with 2,545 SPARK participants with ASD and known IQ scores. We denote the genes contributing to ASD primarily through *de novo* LoF variants in our analysis as “*De novo*” (in red), and the genes primarily through inherited LoF variants as “Inherited” (in blue). (A) Distribution of predicted IQ between individuals with ASD with LoF mutations in the five novel genes, 9 known genes and all participants with ASD and known IQ scores in SPARK (n =2,545). We compared the mean predicted IQ between participants with LoF mutations in ASD genes and all participants by two-sample t-test. Significance level is denoted by the star sign above each violin plot (*: 0.01 ≤ p<0.05, **: 0.001≤p<0.01, ***: p<0.001). Individuals with pathogenic variants in de novo risk genes have significantly lower predicted IQ than overall SPARK participants with ASD and known IQ scores, while individuals with LoF variants in moderate risk, inherited genes with show similar predicted IQ as the overall SPARK participants, with the exception of *ITSN1*. (B) Distribution of predicted IQ between individuals with ASD gene grouped by both inheritance status (“De novo” or “Inherited”) and whether the ASD genes are novel (“Novel” or “Known”). We compared the mean predicted IQ between individuals with pathogenic variants in “De novo” genes and “Inherited” genes among our five novel genes and nine known genes. Overall, people with LoF mutations in “De novo” genes have an average of 13-16 points lower predicted IQ than individuals with LoF mutations in “Inherited” genes, regardless of whether the ASD genes are novel or known. (C) Average relative risk of ASD and average predicted IQ among different groups. Each dot shows the average of individuals with rare LoFs of a gene selected in panel A. The relative risk is estimated from mega analysis and capped at 60. Pearson correlation between average IQ and log relative risk is - 0.78 (p=0.001). The horizontal line represents the average IQ (IQ=79) of all SPARK individuals with predicted IQs. *ITSN1* is an outlier at the bottom left corner.

Most known ASD/NDD genes that are enriched by *de novo* LoF variant harbor more *de novo* than inherited HC LoF variants in ∼16,000 unrelated ASD trios (**Figure 5A and Supplementary Figure S15**), consistent with their high penetrance for ASD/NDD phenotypes and strong negative selection. Using population exome or WGS data, we calculated a point estimate of selection coefficient (*ŝ*)^49^ of LoFs in each gene (**Supplementary Table S8**) and found that the fraction of *de novo* LoFs in ASD genes is higher in genes with large *ŝ*, and smaller in genes with small *ŝ* (**Supplementary Figure S7B**), consistent with population genetic theory^50^. We also estimated average effect size of rare LoFs in ASD genes by comparing cumulative allele frequency (CAF) in 31,976 unrelated cases and population exome or WGS data. As expected, known and newly significant ASD genes with higher risk to ASD are under stronger selection (larger *ŝ*) (**Supplementary Figure S16**).

### Functional similarity of new genes with known ASD genes

To better appreciate the probable functional implications of the new exome-wide significant genes that confer inherited risk for ASD, we integrated mechanistic (STRING^102^) and phenotypic (HPO^103^) data into a single embedding space (six dimensions, one for each archetype coefficient) using a combination of canonical correlation analysis and archetypal analysis. This embedding space serves as an interpretive framework for putative ASD risk genes (N=1,776). Six functional/phenotypic archetypes were identified (**Figure 7**) that represent pathways that are well-understood to play a role in ASD: neurotransmission (archetype 1 or A1), chromatin modification (archetype 2 or A2), RNA processing (archetype 3 or A3), membrane trafficking and protein transport (archetype 4 or A4), extracellular matrix, motility, and response to signal (archetype 5 or A5), and KRAB domain and leucine-rich region proteins (archetype 6 or A6), also enriched for intermediate filaments. These archetypes organize risk genes in a way that jointly maximizes their association with mechanisms (STRING clusters) and phenotypes (HPO terms). For instance, A1 genes (neurotransmission) are enriched for the STRING cluster CL:8435 (ion channel and neuronal system) and are also associated with seizure and epileptic phenotypes. A2 genes (chromatin modifiers) are enriched for nuclear factors and genes linked to growth and morphological phenotypes (**Supplementary Table S10**). We call genes that strongly map to an archetype (i.e., > 2x the next highest-ranking archetype) “archetypal” and “mixed” if this criterion is not met (see methods). Archetypal genes are generally less functionally ambiguous than “mixed” genes. Of the five novel inherited risk genes, two are archetypal (suggesting function within known risk mechanisms): *NAV3* (A6: KRAB domain & LRR) and *ITSN1* (A4: membrane trafficking and protein transport). *SCAF1, MARK2*, and *HNRNPUL2* are mixtures of the identified archetypes, largely A4 and A5. That these new genes did not resolve clearly into archetypes (that were defined by known and suspected autism risk genes) suggests that they may operate in potentially novel or under-appreciated mechanisms. To elucidate these possibilities, we constructed an *ad hoc* “archetype,” defined by the centroid between *SCAF1, MARK2*, and *HNRNPUL2* (see Figure 7C). Cell-cell junction (CL:6549) was the STRING cluster most associated with this centroid (p =4.12 × 10^−14^ by the K-S test, Fig. 7D), which fits with its location between A4 (membrane trafficking) and A5 (ECM).

**Figure 7.**
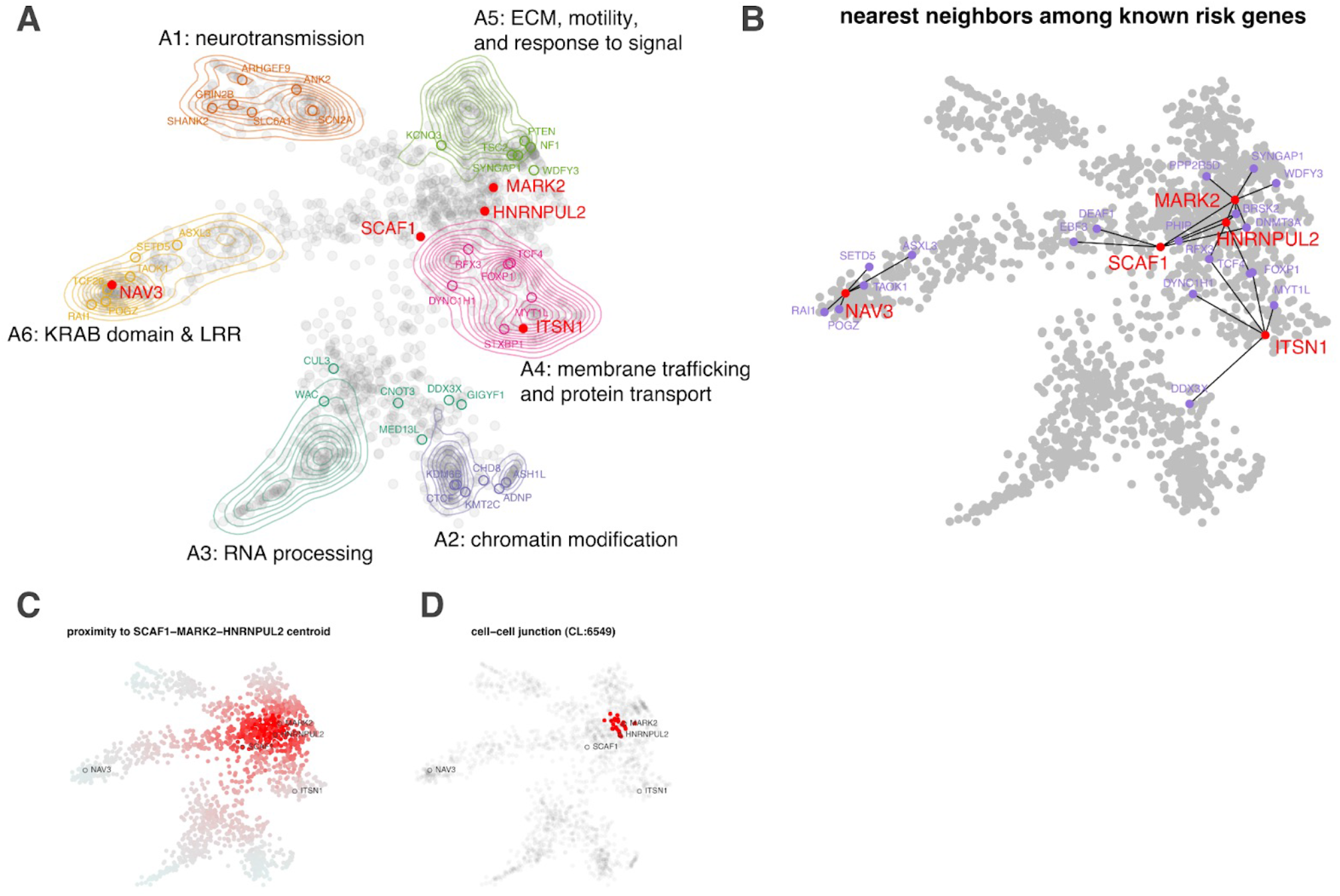
Functional/phenotypic embedding of ASD risk genes. Using a combination of archetypal analysis and canonical correlation analysis, putative autism risk genes were organized into *k=*6 archetypes that represent distinct mechanistic (STRING) and phenotypic (HPO) categorizations (A; neurotransmission, chromatin modification, RNA processing, transport, extracellular matrix, motility and response to signal, and leucine-rich repeat/KRAB domain containing genes). Genes implicated by our meta-analysis are indicated by their label, with novel genes indicated in red. For each of the five novel genes, we identified the five nearest neighbors in the embedding space among the 62 meta-analysis genes (B). *SCAF1, MARK2*, and *HNRNPUL2* were identified as “mixed” rather than “archetypal” in their probable risk mechanisms. To gain further insight into possible risk mechanisms, we calculated the embedding distance to the centroid of these three genes (C), which was then used as an index variable to perform gene set enrichment analysis. A STRING cluster (CL:6549) containing genes related to cell-cell junctions and the gap junction was identified as being highly localized in this region of the embedding space (p =4.12 × 10^−14^ by the KS test) (D). This may suggest that these genes confer autism risk through dysregulation of processes related to cell adhesion and migration.

### Power analysis

The power of identifying risk genes with rare or *de novo* variants monotonically increases with increasing effect size or expected CAF under the null. New ASD genes to be discovered are likely to have smaller effect size than known ASD genes, as suggested by our results. Additionally, known ASD genes are biased toward longer genes with higher background mutation rate of damaging variants (“long genes”) (**Supplementary Figure S17**). Even though longer genes are more likely to be expressed in brain and relevant to ASD/NDD^51^, among most constrained genes, long genes (LoF mutation rate^52,53^ above 80% quantile) and short genes (below 80%) have similar enrichment of damaging *de novo* variants and rare inherited LoFs (**Supplementary Figure S18**). Notably, for small genes, known genes have virtually no contribution to over-transmitted HC LoFs to affected offspring (**Supplementary Figure S18B**). It suggests that many smaller genes contributing to ASD risk remain to be identified. We focus on the power of detecting new ASD genes with a moderate effect size and the full range of background mutation rate.

We use a published framework^41^ to analyze power based on case-control association of rare variants. For rare variants in genes under strong selection, CAF is largely determined by mutation rate and selection coefficient^41^. We therefore modeled power of discovering risk genes as a function of relative risk and selection coefficient. With about 5,500 constrained genes, the power of the current study was calculated for 31,976 unrelated cases and experiment-wise error rate of 9e-6 (**Supplementary Figure S19**).

We inversed the power calculation to determine required sample size to achieve 90% power under the same assumptions (**Supplementary Figure S20**). For genes at median LoF mutation rate across all genes, we estimated that it requires about 96,000 cases (three times the current sample size) to identify genes with similar effect size as *NAV3* (RR=4.5) and *ITSN1* (RR=5), about 64,000 (twice the current sample size) to find genes with similar effect sizes as *SCAF1* (RR=8) and *HNRNPUL2* (RR=9). We note that it requires 10 and 5 times the current sample size to detect these types of genes by *de novo* variants alone.

## Discussion

In this study, we assembled the largest sequencing data set of individuals with ASD to date, including 35,130 ASD cases and their family members collected by SPARK. We characterized the contribution of rare inherited variants to ASD risk and identified five new ASD risk genes by both *de novo* and rare inherited coding variants. We identified rare LoF variants in new ASD risk genes with modest effect size that are not strongly associated with ID. This finding represents a difference in phenotypic association with ID compared with other well-established, highly penetrant ASD genes. To find new risk genes with relative risks of 2-5 (comparable to the low relative risk genes from this study: *NAV3* and *ITSN1*) in the 50-percentile for gene-wide LoF mutation rate (2e-6) and the 50-percentile for selection among known risk genes (0.2), our power analysis suggests that 52,000, 73,000, 116,000 or 227,000 total ASD cases are necessary, respectively (cf. eq 1 from power calculation in Supplementary material). Larger ASD cohorts with phenotypic data will be necessary to identify new ASD risk genes and may help to understand the biology of core symptoms of ASD in individuals without ID.

Our results suggest that identification of new risk genes with rare inherited variants can substantially improve genetic diagnostic yield. We found that rare inherited LoF variants account for 6% of PAR, similar to *de novo* LoF variants. Over two thirds of the PAR from *de novo* coding variants are explained by known ASD or NDD genes. In contrast, less than 20% of PAR from rare inherited LoFs variants is explained by known genes, suggesting most genes contributing to ASD risk through rare inherited variants are yet to be discovered. These unknown risk genes are still largely constrained to LoFs in the general population and/or have similar expression profiles in developing brains to known ASD risk genes. Combining evidence from both *de novo* and rare inherited variants, we identified 60 genes associated with ASD with exome-wide significance, including five novel genes. Rare LoFs in these five new genes account for a PAR of 0.27%, about half of the PAR of the 5 most common highly penetrant ASD genes (*KDM5B, GIGYF1, CHD8, SCN2A, SHANK3*).

*NAV3*, to our knowledge, is the first autosomal ASD risk gene discovered by association of solely rare inherited variants. Carriers of rare LoFs in *NAV3* have an average predicted IQ of 81, slightly above the SPARK cohort average (79). The prevalence of ID among *NAV3* heterozygotes is similar to the SPARK cohort average. This is distinctly different from established ASD risk genes (e.g., *CHD8, SHANK3, SCN2A*), nearly all identified by highly penetrant *de novo* variants, associated with ID in ASD cohorts^2^. The absence of ID is also observed in other genes (e.g., *SCAF1, HNRNPUL2, GIGYF1, KDM5B, KMT2C*) with substantial contribution from rare inherited variants and modest effect size. Nevertheless, the data show that variants in these new ASD genes have effects on core symptoms of ASD, cognition, and other behaviors including schizophrenia, Tourette syndrome, ADHD and other behavioral conditions. Detailed phenotyping of individuals carrying these rare inherited variants is needed to understand the phenotypic effects of each gene. Such strategies should include a genetic and phenotypic assessment of family members who also carry the rare variant but may not have an ASD diagnosis. Since all individuals consented in SPARK are re-contactable, such studies will enable a more complete picture of the broad phenotypic effects of these variants without the bias of clinical ascertainment. Overall, these risk genes with modest effect size may represent a different class of ASD genes that are more directly associated with core symptoms of ASD and/or neuropsychiatric conditions rather than global brain developmental and ID.

The approaches employed in this study made full use of rare variation, and this analytical method is generalizable to many conditions. In particular, the multiple methods used to reduce noise in LoF alleles present in control samples were particularly effective in assessing the signal within the novel genes of moderate effect. We also leveraged gene expression profiles informed by machine learning methods to help prioritize genes for the meta-analysis stage of our analysis^38^. Future studies that leverage additional multi-omic data such as dGTEx may further improve signal to noise.

Our archetypal analysis provides some clues as to the potential risk mechanisms of the five newly identified risk genes. *ITSN1* was unambiguously mapped to A4: membrane trafficking and protein transport and has a role in coordinating endocytic membrane traffic with the actin cytoskeleton^53,54^ *NAV3* (A6: KRAB domain and LRR), is associated with both axon guidance^55^ and malignant growth and invasion^56^ and is thought to regulate cytoskeletal dynamics. Indeed, A6 is enriched for processes related to intermediate filaments (**Supplementary Table S10**) a known determinant of cell motility and polarity^57^. Although *MARK2, SCAF1*, and *HNRNPUL2* were not identified as archetypal (potentially suggesting divergence from well-known autism risk mechanisms) a search for functional enrichment of this interstitial region between A4 and A5 found that their roles in developmental risk may be most relevant at the cell-cell junction, particularly as it relates to migration (see **Figure 7D**).

Taken together, our results suggest that a continued focus on *de novo* variants for ASD gene-discovery may yield diminishing returns. By contrast, studies designed to identify genomic risk from rare and common inherited variants will not only yield new mechanistic insight but help explain the high heritability of ASD. SPARK is designed to recruit individuals across the autism spectrum, without relying on ascertainment at medical centers. As a result, SPARK may be better suited to identify genes with transmitted variants that have lower penetrance and to identify the genetic contributions to the full spectrum of autism. The strategies employed by SPARK — to recruit and assess large numbers of individuals with autism across the spectrum and their available family members without costly, in-depth clinical phenotyping — is necessary to achieve the required sample size to fully elucidate genetic contributions to ASD. SPARK’s ability to recontact and follow all participants will also be critical to deeply assess the phenotypes associated with the newly discovered genes and to develop and test novel treatments.

## Methods

We performed an integrated analysis of coding variants in over 35,130 new ASD cases in SPARK and additional cases from previously published autism cohorts (ASC^3,8^, MSSNG^6^, and SSC^2,30^), using a two-stage analysis workflow (**0 1**). In the first stage, we analyzed over 10,000 ASD cases from family-based samples and systematically compared damaging DNVs and rare, inherited LoF variants. Then we performed an exome-wide scan of genes enriched by DNVs in ASD cases and prioritized genes with suggestive evidence of DNV enrichment. We filtered for high-confidence (HC) LoF variants and searched for genes enriched by inherited HC LoFs using a transmission disequilibrium test (TDT)^54^. In the second stage, we added 22,764 ASD cases and used meta-analysis to further assess the prioritized genes for enrichment of DNVs and TDT of HC LoFs. For LoF intolerant genes, we compared frequency of HC LoF variants in unrelated cases, population controls, and pseudo-controls in ASD families. Finally, we performed a case-control analysis of ASD cases vs population controls to estimate effect sizes for known and newly significant genes and used them for power calculations to estimate sample sizes needed for future studies.

### ASD Cohorts

#### SPARK

We established SPARK (Simons Foundation Powering Autism Research for Knowledge) cohort to facilitate genotype driven research of ASD at scale^23^. Eligibility criteria for SPARK study is residence in the United States and a professional diagnosis of ASD or a family member of a proband in SPARK. SPARK has recruited over 50,000 re-contactable families with ASD cases at 31 different clinical centers across the United States as well as through social and digital media. Individuals with known genetic diagnoses and individuals with and without a family history of autism are included. Whenever possible, parents and family members with or without autism were enrolled and included in the genetic analysis.

Saliva was collected using the OGD-500 kit (DNA Genotek) and DNA was extracted at PreventionGenetics (Marshfield, WI). The samples were processed with custom NEB/Kapa reagents, captured with the IDT xGen capture platform, and sequenced on the Illumina NovaSeq 6000 system using S2/S4 flow cells. Samples were sequenced to a minimum standard of >85% of targets covered at 20X. 97% of samples have at least 20x coverage in >95% of region (99% of samples — in 89% of regions). Pending sample availability, any sample with 20X coverage below 88% was re-processed and the sequencing events were merged to achieve sufficient coverage. The Illumina Infinium Global Screening Array v1.0 (654,027 SNPs) was used for genotyping. The average call rate is 98.5%. Less than 1% of samples have a call rate below 90%.

In the first stage of analysis, we included 28,649 SPARK individuals including 10,242 ASD cases from over 9,000 families with exome sequencing data that passed QC (**Error! Reference source not found**.). A subset of 1,379 individuals was part of the previously published pilot study^7^. To replicate prioritized genes from the discovery stage, we performed a second stage analysis that included an additional 39,926 individuals with 16,970 ASD cases from over 20,000 families with exome or whole genome sequencing (WGS) data available after of the analysis in discovery cohort was completed. For new samples in this study, exome sequences were captured by IDT xGEN research panel and sequenced on the Illumina NovaSeq system. DNA samples were also genotyped for over 600K SNPs by Infinium Global Screening Array.

We used KING^55^ to calculate statistics for pairwise sample relatedness from genotypes of known biallelic SNPs, and validated participant-reported familial relationships (**Supplementary Figure S21A-B**). The relatedness analysis also identified cryptically related families that are connected by unreported parent-offspring or full sibling pairs. Pedigrees were reconstructed manually from inferred pairwise relationships and validated by PRIMUS^56^ and we used inferred pedigree for all analyses. Sample sex was validated by normalized sequencing depths or array signal intensities of X and Y chromosomes which also identified X and Y chromosome aneuploidies (**Supplementary Figure S21C-D**). To infer genetic ancestry, we first performed principal component (PC) analysis on SNP genotypes of non-admixed reference population samples from 1000 Genomes Projects^57^ (Africans, Europeans, East Asians and South Asians) and Human Genome Diversity Project^58,59^ (Native Americans), then projected SPARK samples onto PC axes defined by the five reference populations using EIGNSOFT^60^ (**Supplementary Figure S22**). The projected coordinates on first four PC axes were transformed into probabilities of five population ancestries using the method of SNPweights^61^. The inferred ancestral probabilities show general concordance with self-reported ethnicities (**Supplementary Figure S22B**). Samples were predicted from a reference population if the predicted probability was >=0.85.

The phenotypes of participants are based on self- or parent-report provided at enrollment and in a series of questionnaires from the Simons Foundation Autism Research Initiative database, SFARI Base. We used SFARI Base Version 4 for the discovery cohort and Version 5 for the replication cohort. In the discovery cohort, information about self-reported cognitive impairment (or intellectual disability/developmental delay) was available for 99.2% of ASD cases and 83.5% of other family members at recruitment or from the Basic Medical Screening Questionnaire available on SFARIbase. For phenotype-genotype analyses in individuals with variants in specific ASD risk genes, we defined an individual as having cognitive impairment if 1) there was self- or parent-report of cognitive impairment at registration or in the Basic Medical Screening Questionnaire, 2) the participant was at or over the age of 6 at registration and was reported to speak with less than full sentences or the participant was at or above age 4 at registration and reported as non-verbal at that time, 3) the parent reported that cognitive abilities were <u>significantly</u> below age level, 4) the reported IQ or the estimated cognitive age ratio (ratio IQ^62,63^) was <80 or 5) the parent reported unresolved regression in early childhood without language returning and the participant does not speak in full sentences. The continuous full-scale IQ was imputed based on a subset of 521 samples with full scale IQ and phenotypic features by the elastic net machine learning model^48^. In a subset of cases for which full-scale IQ data or standardized Vineland adaptive behavior scores (version 3) was available, we found self-reported cognitive impairment shows higher correlation with Vineland score than full-scale IQ (**Supplementary Figure S23**). ASD cases with self-reported cognitive impairment were defined as Cognitively Impaired cases, and other cases as Not Cognitively Impaired cases. Other non-ASD family members were considered as unaffected if they were also not indicated to have cognitive impairment. In total of 18.5% families, proband has at least one first-degree relative with ASD who was recruited in the study and/or reported by a family member. Those families were referred to as multiplex, and other families with only a single ASD individual as simplex. The majority (>85%) of affected relative pairs in multiplex families were siblings. Multiplex families have slightly lower male-to-female ratio and lower proportion of cognitive impairment among affected offspring (**Supplementary Figure S24A-B**). In comparison, only 1% of parents in the discovery cohort are affected of which two thirds are females and less than 3% have cognitive impairment (**Supplementary Figure S24A-B**). In addition, non-ASD family members in multiplex families show significantly higher frequency of self-reported cognitive impairment, learning/language disorders, other neuropsychiatric conditions, and other types of structural congenital anomalies (**Supplementary Figure S24C**). Non-ASD parents in multiplex families also have lower educational attainment (**Supplementary Figure S24D**).

##### SSC

SSC (Simon Simplex Collection) collected over 2,500 families with only one clinically confirmed ASD cases who have no other affected first or second degree relatives as an effort to identity *de novo* genetic risk variants for ASD^64^. SSC data have been published before^2,19,30,65^. Here we included 10,032 individuals including 2,633 cases with exome or WGS data available and passed QC (**Error! Reference source not found**.). The data were reprocessed using the same pipeline as SPARK. For 91 trios that are not available or incomplete, we collected coding DNVs from published studies^2,30^. In analysis to associate genetic variants with phenotype severity, we used standardized Vineland adaptive behavior score to group affected cases because it shows higher correlation than full-scale IQ with self-reported cognitive impairment in SPARK (**Supplementary Figure S23**). Cases with cognitive impairment in SSC were defined by Vineland score<=70, and cases with no cognitive impairment by score>70.

##### ASC

ASC (Autism Sequencing Consortium) is an international genomics consortium to integrate heterogenous ASD cohorts and sequencing data from over 30 different studies^66^. Individual level genetic data are not available. So we included 4,433 published trios (4,082 affected and 351 unaffected) merged from two previous studies^3,8^ for DNV analysis. To define low and high functioning cases, we used binary indicator of intellectual disability which was available for 66% of cases. Families with multiple affected trios are considered multiplex, others are simplex.

##### MSSNG

The MSSNG initiative aims to generate WGS data and detailed phenotypic information of individuals with ASD and their families^6^. It comprehensively samples families with different genetic characteristics in order to delineate the full spectrum of risk factors. We included 3,689 trios in DB6 release with whole genome DNV calls are available and passed QC in DNV analysis, of which 1,754 trios were published in the previous study^6^. A total of 3,404 offspring with a confirmed clinical diagnosis of ASD were included as cases. Among individuals without a confirmed ASD diagnosis, 222 who did not show broader or atypical autistic phenotype or other developmental disorders were used as part of controls. Multiplex families were defined as families having multiple affected siblings in sequenced trios or in phenotype database.

Information about cognitive impairment was not available at the time of analysis.

###### Variant calling and quality control

Supplementary Table S11 describes software version and parameter settings for each analysis below.

###### Data processing

Sequencing reads were mapped to human genome reference (hg38) using bwa-mem^67^ and stored in CRAM format^68^. Duplicated read pairs in the same sequencing library of each individual were marked up by MarkupDuplicates of Picard Tools^69^. Additional QC metrics for GC bias, insert size distribution, hybridization selection were also calculated from mapped reads by Picard Tools^69^. Mosdepth^70^ was used to calculate sequencing depth on exome targets (or 500 bp sliding windows for WGS) and determine callable regions at 10X or 15X coverage. Cross-sample contamination was tested by VerifyBamID^71^ using sequencing only mode. Samples were excluded if it has insufficient coverage (less than 80% targeted region with >=20X), shows evidence of cross-sample contamination (FREEMIX>5%), or discordant sex between normalized X and Y chromosome depth and self/parent reports that cannot be explained by aneuploidy.

Variants for each individual were discovered from mapped reads using GATK HaplotypeCaller^72^, weCall^73^, and DeepVariant^74^. Individual variant calls from GATK and weCall were stored in gVCF format and jointly genotyped across all samples in each sequencing batch using GLnexus^75^.

Variants were also jointly discovered and genotyped for individuals of the same family using GATK HaplotypeCaller^72^ and freebayes^76^, and then read-backed phased using WhatsHap^77^. To verify sample relatedness, identify overlapping samples with other cohorts, and verify sample identity with SNP genotyping data, genotypes of over 110,000 known biallelic SNPs from 1000 Genomes or HapMap projects that have call rate >98% and minor allele frequency (MAF) >1% in the cohort were extracted from joint genotyping VCFs. SNP array genotypes were called by Illumina GenomeStudio. We kept samples with >90% non-missing genotype calls and used genotypes of over 400,000 known SNPs that have call rate >98% and MAF>0.1 for relatedness check and ancestry inference.

###### De novo variants

We identified candidate *de novo* SNVs/indels from SPARK and SSC cohorts from per-family VCFs generated by GATK and freebayes and cohort-wide population VCF by weCall using a set of heuristic filters that aim to maximize the sensitivity while minimizing false negatives in parents^7^. We then reevaluated the evidence of all de novo candidates from all input sources. Candidate was removed if there was contradictory evidence against from any input source (“contradiction filters”, see **Supplementary Table S11**). Further, we only kept candidates if they can be called by DeepVariant in offspring but have no evidence of variant in parents. For candidates that were identified in multiple offspring (recurrent), we only kept the ones that passed DeepVariant filter in all trios. For candidates that were shared by siblings in the same family, we only kept the ones with de novo quality estimated by triodenovo higher than 8 (or 7 for SNVs in CpG context). Before creating the final cleaned call set, we selected subsets of variants (see **Supplementary Table S11**) for manual evaluation by IGV to filter out candidates with failed review. Finally, we merged nearby clustered de novo coding variants (within 2bp for SNVs or 50bp for indels) on the same haplotype to form multi-nucleotide variants (MNVs) or complex indels. We removed variants located in regions known to be difficult for variant calling (HLA, mucin, and olfactory receptors). DNVs in the final call set follow a Poisson distribution with an average 1.4 coding DNVs per affected and 1.3 per unaffected offspring (**Supplementary Figure S25**). The proportion of different types of DNVs, the mutation spectrum of SNVs, and indel length distributions were similar between SPARK and SSC (**Supplementary Figure S25**). A small fraction of variants in the final call set are likely post-zygotic mosaic mutations (**Supplementary Figure S26**).

###### Rare variants

Rare variant genotypes were filtered from cohort-wide population VCFs with QC metrices collected from individual and family VCFs (**Supplementary Figure S27A**). Briefly, we initially extracted high quality genotypes for each individual for variants that appear in less than 1% of families in the cohort. Evidence for the variant genotypes were re-evaluated by DeepVariant from aligned reads and collapsed over individuals to create site level summary statistics including fraction of individual genotypes that passed DeepVariant filter and mean genotype quality over all individuals. For variant genotypes extracted from GLnexus VCFs, we re-examined variant genotype from per-family VCFs by GATK to collect GATK site level metrics (including QD, MQ, SOR, etc.) then took read-depth weighted average over families to create cohort-wide site metrics. For variant genotypes extracted GATK joint genotyping VCFs, these site metrics were directly available directly from INFO fields.

Variant site level QC filters were calibrated using familial transmission information, assuming that false positive calls are more likely to show Mendelian inheritance error (**Supplementary Figure S27B**). Briefly, we first applied a baseline site level filter that favors high sensitivity, then optimized thresholds for filters with additional QC metrics. The selected QC metrics were reviewed first to determine a small number of optional thresholds. Then the final set of QC parameters were optimized from a grid search over the combinations of available thresholds such that: 1. presumed neutral variants identified from parents (silent variants or variants in non-constrained genes) shows equal transmission and non-transmission to offspring; 2. rates of neutral variants are similar in different sample groups from the same population ancestry; 3. vast majority variants identified in trio offspring are inherited from parents. In case when multiple sets of QC thresholds give similar results, priority will be given to the set that also recovers maximum number of DNV calls in trio offspring. The optimized filtering parameters were used in final QC filters to generate analysis-ready variants.

For a rare coding variant initially annotated as LoF (including stop gained, frameshift, or splice site), we searched for nearby variants on the same haplotype (within 2bp for SNVs or 50bp for indels). If nearby variants can be found, they were merged to form MNVs or complex indel and re-annotated to get the joint functional effect. If the joint effect was not LoF, then the original variant was removed from LoF analysis.

###### Variant annotations

The genomic coordinates of QC passed variants were lifted over to hg19 and normalized to the leftmost positions^78^. Functional effects of coding variants were annotated to protein coding transcripts in GENCODE V19 Basic set^79^ using variant effect predictor^80^. The gene level effect was taken from the most severe consequences among all transcripts (based on the following priority: LoF>missense>silent>intronic). pExt for each variant can be operationally defined as the proportion of expression levels of transcripts whose variant effects are the same as gene effect over all transcripts included in the annotation^27^. We used transcript level expressions in prenatal brain development from Human Developmental Biology Resource^81^ to calculate pExt. Missense variants were annotated by pathogenicity scores of REVEL^31^, CADD^82^, MPC^83^ and PrimateAI^84^. Population allele frequencies were queried from gnomAD^26^ and ExAC^18^ using all population samples. All rare variants were defined by cohort allele frequency <0.001 (or <0.005 for X chromosome variants). To filter for ultra-rare variants, we keep variants with cohort allele frequency <1.5e-4 (or allele count=1) and population allele frequency <5e-5 in both gnomAD^26^ and ExAC^18^.

LoF variants on each coding transcript were further annotated by LOFTEE^26^ (v1.0, default parameters). We also annotated splice site variants by SpliceAI^85^, and removed low confidence splice site variants with delta score <0.2 from LoF variants. pExt for LoF variants was calculated by the proportion of expression level of transcripts that harbor HC LoFs evaluated by LOFTEE over all transcripts included in the analysis. Thus, the pExt filter for LoFs already incorporated LOFTEE annotations. The baseline filter to analyze rare, inherited LoFs and LoFs of unknown inheritance is pExt>=0.1. To refine gene-specific pExt threshold in the second stage, we selected 95 known ASD/NDD genes plus a newly significant DNV enriched gene *MARK2* which harbor at least four *de novo* LoF variants in combined ASD and other NDD trios, and for each gene choose the pExt threshold from {0.1,0.5,0.9} that can retain all *de novo* LoF variant with pExt>=0.1 (Supplementary Table S1).

###### Copy number variants

Copy number variants (CNVs) were called from exome read depth using CLAMMS^86^. CNV calling windows used by CLAMMS were created from exome targets after splitting large exons into equally sized windows of roughly 500bp. Calling windows were annotated by average mappability score^87^ (100mer) and GC content assuming average insert size of 200. Depths of coverage for each individual on the windows were calculated using Mosdepth^70^ and then normalized to control for GC-bias and sample’s overall average depth. Only windows with GC content between 0.3 and 0.75 and mappability >=0.75 were included in further analyses. For each given sample, we used two approaches to reduce the dimension of sample’s coverage profile and automatically selected 100 nearest neighbors of the sample under analysis as reference samples. The first approach used seven QC metrics calculated by Picard Tools from aligned reads as recommended by the CLAMMS developer^86^, we further normalized those metrics in the cohort by its median absolute deviation in the cohort. The second approach used singular value decomposition of the sample by read-depth matrix to compute the coordinates of the first 10 principal components for each sample.

Model fitting and CNV calling for each individual using custom reference samples were performed using default parameters. From raw CNV calls, neighboring over-segmented CNVs of the same type were joined if joined CNVs include over 80% of the calling windows of original calls. For each sample, we kept CNV calls made from one set of reference samples that have smaller number of raw CNV calls. Outliers with excessive raw CNV calls (>400) were removed. For each CNV, we counted the number of CNVs of the same type in parents that overlap >50% of the calling windows. High-quality rare CNVs were defined as <1% carrier frequency among parents and have Phred-scaled quality of CNV in the interval >90. We queried high-quality rare copy number deletions to look for additional evidence to support new genes.

##### Genetic analysis

###### De novo variants analysis

In the discovery stage analysis, the DNV call sets of SPARK and SSC were merged with published DNVs from ASC^3,8^ and MSSNG^6^ and additional SSC trios of which we did not have sequencing data. To infer likely samples overlaps with published trios of which we do not have individual level data, we tallied the proportion of shared DNVs between all pairs of trios. For a pair of trios, let *N*_1_ and *N*_2_ be the number of coding DNVs and *O* the number of shared DNVs between pair. To account for mutation hotspots, if a DNV is a SNV within CpG context or a known recurrent DNVs identified in SPARK and SSC, it contributes 0.5 to the count. Likely overlapping samples were identified if 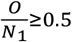 or 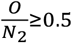 and they have identical sex.

To determine the expected number of DNVs in the cohort, we used a 7-mer mutation rate model^52^ in which the expected haploid mutation rate of each base pair (bp) depends on the 3bp sequence context on both sides. The per-base mutation rates were adjusted by the fraction of callable trios at each base pair which was the fraction of trios with >=10X coverage in parents and >=15X coverage in offspring. For published trios, we used an inhouse WGS data of 300 trios with average 36X coverage to approximate the callable regions. Gene level haploid mutation rates for different classes of DNVs were calculated by summing up the depth-adjusted per-base mutation rate of all possible SNVs of the same class. The rate for frameshift variants was presumed to be 1.3 times the rate of stop gained SNVs^53^. Mutation rates in haploid X chromosome regions were adjusted for the observed male-female ratio (4.2) assuming mutation rates in spermatogenesis is 3.4 times higher than oogenesis^9^. The exome-wide rate of synonymous DNVs closely matches the observed number of DNVs (**Supplementary Figure S12**). We also observed similar fold enrichment of damaging DNVs (vs. expected rate) in ASD cases across four cohorts after accounting for samples with family history (**Supplementary Figure S12**).

To perform gene-based test of DNVs, we applied DeNovoWEST^11^ a simulation-based approach to test the enrichment of weighted sum of different classes of DNVs compared to the expected sum based on per-base mutation rates in each gene. We used empirical burden of DNVs to derive weights for different variant classes in constrained genes (ExAC pLI>=0.5) and non-constrained genes separately based on positive predictive values (PPV) (**Supplementary Table S13**). For ASD, we defined *de novo* D-mis variants by REVEL score >=0.5, and the rest of *de novo* missense variants are taken as benign missense (B-mis). For other NDDs, we defined two classes of *de novo* D-mis variants by MPC score>=2 or MPC<=2 and CADD score>=25, and the remaining *de novo* missense variants are B-mis. We first ran DeNovoWEST to test the enrichment of all nonsynonymous DNVs (pEnrichAll). To account for risk genes that harbor only missense variants, we ran DenovoWEST to test the enrichment of *de novo* missense variants only and applied a second test for spatial clustering of missense variants using DenovoNear^9^, then combined evidence of missense enrichment and clustering (pCombMis). The minimal of pEnrichAll and pCombMis was used as the final p-value for DeNovoWEST. The exome-wide significance threshold was set to 1.3e-6 (=0.05/(18,000 genes*2 tests)) to account for the two tests. The analysis on replication cohort used the same weights as derived from discovery cohort. Compared with the original publication^11^, our implementation of DeNovoWEST used different ways to stratify genes, determine variant weights, and calculate per-base mutation rates. We applied our DeNovoWEST implementation on 31,058 NDD trios and compared with published results on the same data set. The p-values from re-analysis show high overall concordance with published results (**Supplementary Figure S28**). We used p-values from our re-analysis on other NDD trios in comparative analysis with ASD.

Gene set enrichment analysis of DNVs was performed by DnEnrich framework^32^. We included all *de novo* LoF and D-mis variants in 5,754 constrained genes from 16,877 ASD and 5,764 control trios. For each gene set, we calculated the fraction of weighted sums of damaging DNVs in the set using PPV weights of constrained genes (**Supplementary Table S13**) for cases and controls respectively. The test statistics for each gene set is the ratio of such fractions in cases over controls. To determine the distribution of test statistic under the null hypothesis, we randomly placed mutations onto the exome of all constrained genes, while held the number of mutations, their tri-nucleotide context and functional impact to be the same as observed in cases and controls separately. Note that by conditioning on the observed number of damaging DNVs in cases and controls, we tested enriched gene sets in cases that are not due to an increased overall burden. At each round of simulation, the permuted test statistic in each gene set was calculated. Finally, the p-value was calculated as number of times the permuted statistic is greater than or equal to observed statistic. Fold enrichment (FE) was calculated as the ratio of between observed and average of test statistics over all permutations. We also approximated 95% confidence interval for FE by assuming log(FE) follows normal distribution with mean 0 and standard deviation determined by the p-value.

In all DNV analyses above, DNVs shared by full or twin siblings represent single mutational events and were counted only once. When an individual carry multiple DNVs within 100bp in the same gene, only one variant with most severe effects was included in the analysis.

##### Transmission disequilibrium analysis

The effect of inherited LoF variants was analyzed using TDT in each individual genes or in gene sets. Rare LoF variants were first identified in parents without ASD diagnoses or intellectual disability who have at least one offspring, then for each parent-offspring pair, the number of times the LoF variant was transmitted from parents to offspring was tallied. For variants in (non-PAR part of) X chromosome, we only used rare LoF variants carried by mothers without ASD diagnoses or intellectual disability and analyzed transmission in different types of mother-offspring pairs. For TDT analysis of rare, inherited missense variants in selected gene sets, different D-mis definitions and allele frequency cutoffs were used (**Supplementary Figure S3**).

The over-transmission of LoFs to affected offspring was evaluated by a binomial test assuming transmission equilibrium under the null hypothesis of 50% chance of transmission. In the discovery stage, ultra-rare LoFs with pExt>=0.1 were used in exome-wide transmission disequilibrium and gene set enrichment analysis. For gene-based test, all rare LoFs with pExt>=0.1 were also used, and TDT statistic^39^ for each gene was calculated by 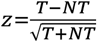, where *T*(*NT*) is the number of times LoF variants were transmitted (not transmitted) to affected offspring. When offspring include monozygotic twin pairs, only one was kept in the transmission analysis. We prioritized 244 autosomal genes with *z*>1 in top 10% LOEUF or in top 20% LOEUF and A-risk>=0.4. In the second stage gene-based test, if a gene-specific pExt threshold is available, we used HC LoF variants passed the gene-specific pExt filter.

In gene set enrichment analysis of inherited LoFs, the rate of transmission to affected offspring in each gene set was compared with the transmission rate in rest of the genes in the background using chi-squared test.

##### Case control analysis

Pseudo-controls are constructed from parents without ASD diagnoses or intellectual disability in simplex families, using alleles that were not transmitted to affected offspring. Each parent without ASD diagnoses or intellectual disability contributes sample size of 0.5 to pseudo-controls. Rare LoFs in ASD cases whose parent data are not available and from other cases that were not utilized in DNV enrichment or TDT analysis were analyzed in this stage. Specifically, for each ASD case, we found out all his/her most recent unaffected ancestors without ASD diagnoses or intellectual disability in the pedigree and calculated the contributing sample size as 1 minus the summation of kinship coefficients with these ancestors. If the contributing sample size is greater than 0, then the sample was included in pseudo-cases after removing alleles that were observed in any unaffected ancestors without ASD diagnoses or intellectual disability used in TDT and alleles included in DNV analysis if any. Examples of such rare LoFs in cases and their contributing sample sizes are given in Supplementary Figure S29.

Rare LoFs in cases and controls for X chromosome were categorized separately for males and females. For male controls, because fathers do not transmit X chromosomes to sons, male controls include all fathers. In contrast, male cases only include those whose mothers do not have ASD diagnoses or intellectual disability (thus not included in TDT analysis). For females, because we only include mothers without ASD diagnoses or intellectual disability and affected sons in TDT, female pseudo-cases include all affected females. Female pseudo-controls were established from unaffected mothers in simplex families using alleles that do not transmit to affected sons. Each unaffected mother contributes a sample size of 0.5 to pseudo-controls. In both sexes, DNVs were removed from pseudo-cases.

For gene-based tests in Stage 2, case-control comparisons are not independent of TDT. So we used population references as controls, including gnomAD exomes^26^ (v2.1.1 non-neuro subset), gnomAD genomes^26^ (v3.1 non-neuro subset), and TopMed genomes^88^ (Freeze 8). Variants in the population references were filtered to keep those passed default QC filter in released data. For variants in gnomAD data set, we further removed variants located in low complexity region, because such regions are enriched with false positive calls^89^ but the default filter does not effectively remove variants in those regions. QC filters in the inhouse ASD cohort and in TopMed had already removed most of variants located in such regions. Variants from population references were re-annotated in the same way as rare variants identified in ASD cohort. In gene level case-control comparison of LoF burden, we used baseline pExt>=0.1 filter or gene-specific pExt threshold if available to define HC LoF variants. For LoF variants in selected genes, we also extracted curation results by gnomAD to remove curated non-LoF variants and manually reviewed IGV snapshots from gnomAD browser if available to remove likely variant calling artifacts (Supplementary Data 1). Number of HC LoF variants were obtained from the summation of allele count in site level VCF files. Gene level burden of HC LoF variants between cases and population controls are tested by comparing the HC LoF variant rates between cases and controls using Poisson test. To account for different in depth of coverage, sample sizes are multiplied by the fraction of callable coding regions of each gene (>=15X for autosomes or female X chromosome, >=10X for male X chromosome) in ASD cases and in population controls respectively.

To account for sample relatedness in case-control analysis, we created a relationship graph in which each node represents an individual and each edge represents a known first or second-degree relationship between two individuals. We also add edges to pairs of individuals without known familial relationship but have estimated kinship coefficient >=0.1. From the graph, we select one individual from each connected component to create unrelated case-control samples. For chromosome X, father and sons were treated as unrelated. For population controls, only gnomAD data included sex specific allele counts and were used in the sex-specific analysis.

Meta-analysis was performed for prioritized autosomal genes among top 30% LOEUF. We integrated evidence from the enrichment of all DNVs, transmission disequilibrium, and increased burden in case compared with population controls by combining p-values using Fisher’s method^40^. Experiment-wide error rate was set at 9e-6 (=0.05 divided by 5340 autosomal genes at LOEUF 30%). In mega-analysis, we combined all unrelated ASD cases together and compared CAFs of HC LoF variants with three population references.

##### Power calculation

To calculate statistical power of the current study and to estimate sample size for future gene discovery efforts, we adopted the statistical framework by Zuk et al. 2014^41^ comparing CAF of LoF variants in *N* unrelated cases *f*_case,_ with CAF *f* in natural population. The effect of LoFs in the same gene are assumed to be the same and increase ASD risk by *γ* fold. The population CAF *f* is assumed to be known with high precision from large cohorts. Since we only focus on LoF-intolerant genes in the population, *f* is assumed to be at selection-mutation equilibrium 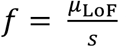 where *μ*_LOF_ is LoF mutation rate and *s* is selection coefficient. The test statistic asymptotically follows a non-central chi-squared distribution with 1-df and non-centrality parameter (NCP):

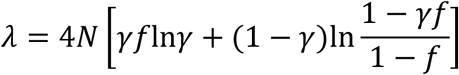

Given the significance threshold *α*, power can be calculated analytically by

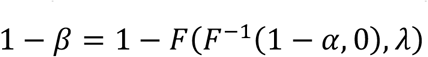

where *F*(*x, λ*) is the cumulative distribution of 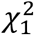 with NCP *λ*.

To calculate sample size to achieve desired power 1 − *β* at significance level *α*, we first solve NCP *λ*_*α,β*_ from the above equation. Then sample size can be approximated by:

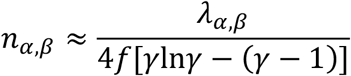

For current study in ASD, sample size is *N*= 31,976 unrelated cases, experimental wide error rate is *α*=9e-6. Given continuing expansion of population reference, treating *f* as known without error is a reasonable assumption for future studies. To calculate power for new genes identified in this study, we used point estimates of *γ* and *f* from mega-analysis using gnomAD exomes as population controls, and used *μ*_LOF_ computed from the 7mer context dependent mutation rate model^52^ to convert *f* to 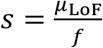. The required sample sizes were calculated to achieve 90% of power.

Power and sample size are both calculated as a function of relative risk for ASD (*γ*) and selection coefficient (*s*) across different haploid LoF mutation rates (*μ*_LOF_). We only considered *s* between 0.01 and 0.5, because most prioritized genes have point estimates of *s*>0.01(Error! Reference source not found.) and genes with *s*>0.5 are expected to harbor to *de novo* than inherited LoF variants and can to be identified from the enrichment of DNVs. Relative risk to ASD (*γ*) was constrained between 1 and 20 since we are mainly interested in discovering genes with moderate to small effects. The reduction in fitness *s* is correlated with the increases in ASD risk *γ* by *s* = *γπs*_*D*_ under the assumption of no pleiotropic effect, where *π* is ASD prevalence and *s*_*D*_ is decreased reproductive fitness of ASD cases. Based on epidemiological studies, current estimated prevalence of ASD is 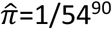, estimated *s*_*D*_ is for 0.75 male and for 0.52 female^91^ so sex averaged *ŝ*_*D*_=0.71 (assuming male-to-female ratio of 4.2). In reality, most known ASD genes also show pleiotropic effects with other NDDs or associated with prenatal death and therefore 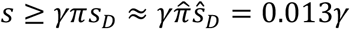. So we only considered combinations of (*s, γ*) that satisfy the condition: *s* ≥ 0.013*γ*.

##### Gene sets

To evaluate the contribution of known ASD risk genes to the burdens of DNVs and inherited LoF variants identified in this study, we collected 618 known dominant ASD/NDD genes from the following sources:

1. Known developmental disorder genes from DDG2P^92^ (2020-02) that are dominant or X-linked and have organ specificity list includes brain or cause multi-system syndrome.
2. High confidence ASD genes collected by SFARI^93^ (2019-08) with score of 1 or 2 excluding known recessive genes.
3. Newly emerging dominant ASD genes reported in recent literatures and included in SPARK genes list^94^ (2020-07).

To evaluate the gene sets enriched by damaging DNVs or inherited HC LoFs, we used all constrained genes by ExAC pLI>=0.5 or in top 20% of LOEUF as the background. Gene sets of the following five categories were collected for gene sets enrichment analysis.

##### Transcriptome and proteome

- For genes with brain-specific expression, we used processed RNA-seq data from Fagerberg *et al*. 2014^95^ and selected genes with average reads per kilobase of transcript per million mapped reads (RPKM)>1 in brain and over four times of median RPKM of 27 tissues.
- Genes in co-expression modules M2 and M3 derived from weighted gene correlation network analysis (WGCNA) analysis of BrainSpan developmental RNAseq data were previously reported to enrich for known ASD genes^33^ and collected from Table S1 from that reference.
- To find genes expressed in excitatory or inhibitory neurons, we selected genes from Mo *et al*. 2015^96^ that have average transcripts per million (TPM) greater than 100 in excitatory and inhibitory neurons respectively.
- Synaptic genes including those encode presynaptic proteins, presynaptic active zone, synaptic vesicles, and postsynaptic density were collected from SynaptomeDB^97^.

##### Neuronal regulome

- Putative CELF4 target genes are defined as genes whose iCLIP occupancy>0.2 in Wagnon *et al*. 2012^98^.
- CHD8 target genes are defined as genes whose promoter or enhancer region overlap with CHD8 binding peaks in human neural stem cells or mid-fetal brain in Cotney *et al*. 2015^36^.
- FMRP target genes in mouse were first collected from Table S2C of Darnell *et al*. 2011^35^ with FDR<0.1. They were then mapped to orthologous human genes using homology mapping provided by MGI^99^ (2018-07).
- Genes targeted by RBFOX2 were selected from Weyn-Vanhentenryck *et al*. 2014^34^ to have Rbfox2 tag counts greater 8. Due to high correlations between RBFOX1 and RBFOX3, targeted genes by the two RNA binding proteins were merged in one gene set and selected to have total tag counts of Rbfox1 and Rbfox3 greater than 24. Selected mouse genes symbols were then mapped to orthologous human genes using homology mapping provided by MGI.

##### Autism gene predictions

- ForecASD is an ensemble classifier that integrates brain gene expression, heterogeneous network data, and previous gene-level predictors of autism association to yield a single prediction score^37^. We created two sets of genes with forecASD prediction score greater than 0.4 or 0.5.
- A-risk is a classifier that uses a used gradient boosting tree to predict autism candidate genes using cell-type specific expression signatures in fetal brain^38^. We created three sets of genes with prediction score greater 0.4, 0.5 or 0.6.

##### Genetic evidence

- For genes enriched by DNVs in ASD, we selected genes showing nominal statistical evidence (P<0.01 or P<0.05 by DeNovoWEST) in discovery cohort of 16,877 trios.
- For genes implicated by in other NDD, we selected genes nominally enriched by DNVs in 31,058 NDDs^11^ (P<0.01 or P<0.01 by DeNovoWEST using our implementation).
- For genes in implicated in schizophrenia, we selected genes nominally significant (P<0.05) by gene-based test in latest schizophrenia case-control study of 24,248 cases and 97,322 controls^47^.

###### Archetypal analysis

STRING v11^100^ clusters and Human Phenotype Ontology (HPO)^101^ terms were formatted as gene-by-term binary matrices. The working gene list was taken as the union of forecASD top decile genes and the 62 autism-associated gene from this study (total 1,776 genes). A total of 583 genes from this set had annotations in both STRING and HPO, and using these genes, a canonical correlation analysis (CCA) was carried out using the RGCCA package for R (https://cran.r-project.org/web/packages/RGCCA/index.html) using five components and sparsity parameter c1 set to 0.8 for both the HPO and STRING matrices. Component scores for all 1,776 genes were calculated using the STRING cluster annotations and the corresponding coefficients from the CCA. This 1,776 gene by 5 CC component matrix was used as input for archetypal analysis ^102^, and the optimal *k* (number of archetypes) was selected using the elbow plot heuristic^103^, with the residual sums of squares (RSS) plotted as a function of *k*. We displayed the archetypal embedding using the simplexplot() function of the archetypes R package. Genes were identified as “archetypal” if their top archetype coefficient was > 2x the next highest archetypal coefficient. Those genes that did not fulfill this criterion were classified as “mixed”, while those that did were assigned to their maximally-scoring archetype. Each of the six identified archetypes were given a human-readable summary description based on review of the top associated STRING clusters (Figure 7). Further cluster/term association results are available in Supplementary Table S10. Representative genes for each archetype were chosen from among the list of 62 risk genes identified in this study, using the top 6 genes for each archetype (note that these genes do not necessarily fulfill the “archetypal” criterion described above, but are simply the top six of the 62 for each archetype).

## Data Availability

All data in this manuscript is currently available to qualified researchers at base.sfari.org

https://base.sfari.org/

## Author Contributions

I.Astrovskaya, J.B.H., J.J.M., N.V., P.F., C.Shu, T.W., W.K.C., X.Z., and Y.S. designed and conceived this study. A.Adams, A.Andrus, A.Berman, A.Brown, A.C., A.C.G., A.D.S., A.E., A.Fanta, A.Fatemi, A.Fish, A.Goler, A.Gonzalez, A.Gutierrez, Jr., A.Hardan, A.Hess, A.Hirshman, A.Holbrook, A.J.A., A.J.Griswold, A.Jarratt, A.Jelinek, A.Jorgenson, A.Juarez, A.Kim, A.Kitaygorodsky, A.L., A.L.R., A.L.W., A.M.D., A.Mankar, A.Mason, A.Miceli, A.Milliken, A.M.-L., A.N.S., A.Nguyen, A.Nicholson, A.Nishida, A.P., A.P.M., A.R.G., A.Raven, A.Rhea, A.Simon, A.Swanson, A.Sziklay, A.Tallbull, A.Tesng, A.W., A.Z., B.A.H., B.B., B.E., B.E.R., B.Hauf, B.J.O., B.L., B.M.V., B.S., B.V., C.A.E., C.A.W.S., C.Albright, C.Anglo, C.B., C.C.B., C.C.-S., C.Cohen, C.Colombi, C.D., C.E., C.E.R., C.Fassler, C.Gray, C.Gunter, C.H.W., C.K., C.Leonczyk, C.L.M., C.Lord, C.M.T., C.M., C.O.-L., C.Ortiz, C.P., C.R.R., C.Roche, C.Shrier, C.Smith, C.V., C.W.-L., C.Zaro, C.Zha, D.B., Dan.Cho, D.Correa, D.E.S., D.G., D.G.A., D.H., D.I., D.L.C., D.Li, D.Limon, D.Limpoco, D.P., D.Rambeck, D.Rojas, D.Srishyla, D.Stamps, E.A.F., E.Bahl, E.B.-K., E.Blank, E.Bower, E.Brooks, E.C., E.Dillon, E.Doyle, E.Given, E.Grimes, E.J., E.J.F., E.K., E.L.W., E.Lamarche, E.Lampert, E.M.B., E.O’Connor, E.Ocampo, E.Orrick, E.P., E.R., E.S., E.T.M., E.V.P., F.F., F.K.M., G.A., G.B., G.D., G.H., G.M., G.S., G.S.D., G.T., H.C., H.E.K., H.G., H.H., H.K., H.L.S., H.Lechniak, H.Li, H.M., H.R., H.Zaydens, I.Arriaga, I.F.T., J.A., J.A.G., J.Beeson, J.Brown, J.Comitre, J.Cordova, J.D., J.F.C., J.F.H., J.Gong, J.Gunderson, J.H., J.J.M., J.Judge, J.Jurayj, J.Manoharan, J.Montezuma, J.N., J.O., J.Pandey, J.Piven, J.Polanco, J.Polite, J.R., J.S., J.S.S., J.T.M., J.Tjernagel, J.Toroney, J.V.-V., J.Wang, J.Wright, K.A., K.A.S., K.Baalman, K.Beard, K.Callahan, K.Coleman, K.D.F., K.Dent, K.Diehl, K.G., K.G.P., K.H., K.L., K.L.P., K.Murillo, K.Murray, K.N., K.O., K.Pama, K.R., K.Singer, K.Smith, K.Stephenson, K.T., L.A., L.A.C., L.Beeson, L.Carpenter, L.Casten, L.Coppola, L.Cordiero, L.D., L.D.P., L.F.C., L.G.S., L.H.S., L.K.W., L.L., L.M.H., L.M.P., L.Malloch, L.Mann, L.P.G., L.S., L.V.S., L.W., L.Y., L.Y.-H., M.A., M.Baer, M.Beckwith, M.Casseus, M.Coughlin, M.Currin, M.Cutri, M.DuBois, M.Dunlevy, M.F., M.F.G., M.G., M.Haley, M.Heyman, M.Hojlo, M.J., M.J.M., M.Kowanda, M.Koza, M.L., M.M., M.N., M.N.H., M.O., M.P., M.R., M.Sabiha, M.Sahin, M.Sarris, M.Shir, M.Siegel, M.Steele, M.Sweeney, M.T., M.V.-M., M.Verdi, M.Y.D., N.A., N.Bardett, N.Berger, N.C., N.D., N.G., N.H., N.Lillie, N.Long, N.M.R.-P., N.Madi, N.Mccoy, N.N., N.Rodriguez, N.Russell, N.S., N.Takahashi, N.Targalia, N.V., O.N., O.Y.O., P.F., P.H., P.M., P.S.C., R.A.B., R.A.G., R.C.S., R.D.A., R.D.C., R.J., R.J.L., R.K.E., R.L., R.P.G.-K., R.Remington, R.S., R.T.S., S.A., S.Birdwell, S.Boland, S.Booker, S.Carpenter, S.Chintalapalli, S.Conyers, S.D., S.D.B., S.E., S.F., S.G., S.Hepburn, S.Horner, S.Hunter, S.J.B., S.J.L., S.Jacob, S.Jean, S.Kim, S.Kramer, S.L.F., S.Licona, S.Littlefield, S.M.K., S.Mastel, S.Mathai, S.Melnyk, S.Michaels, S.Mohiuddin, S.Palmer, S.Plate, S.Q., S.R., S.Sandhu, S.Santangelo, S.Skinner, S.T., S.Xu, S.Xiao, Sa.White, St.White, T.C., T.G., T.H., T.I., T.K., T.P., T.R., T.S., T.Thomas, T.Tran, V.Galbraith, V.Gazestani, V.J.M., V.R., V.S., W.C.W., W.Cal, W.K.C., W.S.Y., Y.C. and Z.E.W. recruited participants and collected clinical data and biospecimens. A.Amatya, A.Bashar, A.E.L., A.Mankar, A.Nguyen, B.J., C.Rigby, Dav.Cho, D.V.M., E.O’Connor, J.A., M.D.M., M.E.B., N.Lawson, N.Lo, N.V., R.M., R.Rana, S.G., S.Jean, S.Shah and W.Chin built and supported the SPARKforAutism.org website, software, databases and systems, and managed SPARK data. A.D.K., A.J.Gruber, A.Nishida, B.Han, B.J.O., C.Fleisch, C.Shu, D.V.M., E.Brooks, G.J.F., I.Astrovskaya, J.B.H., J.J.M., J.U.O., J.Wright, L.Brueggeman, L.G.S., M.A.P., N.Lo, N.V., O.M., P.F., S.D.B., S.Murali, S.X.X., T.N.T., T.S.C., T.W., W.H., X.Z. and Y.S. performed analyses, processed biospecimens and sequenced DNA samples. A.Fatemi, A.Kitaygorodsky, A.Soucy, C.Shu, D.H.G., E.B.-K., E.E.E., E.R., H.Q., H.Zhang, H.Zhao, I.Astrovskaya, J.B.H., J.J.M., J.U.O., J.Wright, L.Brueggeman, L.G.S., M.Y.D., N.V., O.M., P.F., R.N.D., S.D.B., S.Murali, S.X.X., T.K., T.N.T., T.P., T.S., T.Thomas, T.W., T.W.Y., W.K.C., X.Z. and Y.S. helped with data interpretation. A.E.L., E.E.E., J.J.M., N.V., P.F., W.K.C. and X.Z. supervised the work. B.J.O., I.Astrovskaya, J.B.H., J.J.M., J.U.O., C.Shu, J.Wright, N.V., P.F., T.N.T., T.W., W.K.C., X.Z. and Y.S. wrote this paper.

## Competing interests

D.H.G. has received research funding from Takeda Pharmaceuticals, and consulting fees or equity participation for scientific advisory board work from Ovid Therapeutics, Axial Bio-therapeutics, Acurastem, and Falcon Computing. E.E.E. is on the Scientific Advisory Board (SAB) of DNAnexus, Inc. M.Sahin has received research funding from Novartis, Roche, Biogen, Astellas, Aeovian, Bridgebio, Aucta and Quadrant Biosciences and has served on Scientific Advisory Boards for Roche, Celgene, Regenxbio and Takeda. W.K.C. serves on the Regeneron Genetics Center Scientific Advisory Board and is the Director of Clinical Research for SFARI. A.D.K. is an employee of PreventionGenetics and a member of PrevGen Employees LLC, which owns units in PreventionGenetics. Z.E.W. serves as a consultant for Roche and receive research support from Adaptive Technology Consulting. All other authors declare no competing interests.

### The SPARK Consortium

Adrienne Adams^13^, Alpha Amatya^2^, Alicia Andrus^14^, Asif Bashar^2^, Anna Berman^15^, Alison Brown^16^, Alexies Camba^17^, Amanda C. Gulsrud^17^, Anthony D. Krentz^18^, Amanda D. Shocklee^19^, Amy Esler^20^, Alex E. Lash^2^, Anne Fanta^21^, Ali Fatemi^22^, Angela Fish^23^, Alexandra Goler^2^, Antonio Gonzalez^24^, Anibal Gutierrez, Jr.^24^, Antonio Hardan^25^, Amy Hess^26^, Anna Hirshman^13^, Alison Holbrook^2^, Andrea J. Ace^2^, Anthony J. Griswold^27^, Angela J. Gruber^18^, Andrea Jarratt^28^, Anna Jelinek^29^, Alissa Jorgenson^28^, A. Pablo Juarez^30^, Annes Kim^23^, Alex Kitaygorodsky^31^, Addie Luo^32^, Angela L. Rachubinski^33^, Allison L. Wainer^13^, Amy M. Daniels^2^, Anup Mankar^2^, Andrew Mason^34^, Alexandra Miceli^15^, Anna Milliken^35^, Amy Morales-Lara^36^, Alexandra N. Stephens^2^, Ai Nhu Nguyen^2^, Amy Nicholson^30^, Anna Marie Paolicelli^37^, Alexander P. McKenzie^16^, Abha R. Gupta^21^, Ashley Raven^23^, Anna Rhea^38^, Andrea Simon^39^, Aubrie Soucy^40^, Amy Swanson^15^, Anthony Sziklay^34^, Amber Tallbull^33^, Angela Tesng^28^, Audrey Ward^38^, Allyson Zick^23^, Brittani A. Hilscher^41^, Brandi Bell^38^, Barbara Enright^42^, Beverly E. Robertson^2^, Brenda Hauf^43^, Bill Jensen^2^, Brandon Lobisi^24^, Brianna M. Vernoia^2^, Brady Schwind^2^, Bonnie VanMetre^16^, Craig A. Erickson^29^, Catherine A.W. Sullivan^21^, Charles Albright^26^, Claudine Anglo^41^, Cate Buescher^4^, Catherine C. Bradley^38^, Claudia Campo-Soria^28^, Cheryl Cohen^2^, Costanza Colombi^23^, Chris Diggins^2^, Catherine Edmonson^16^, Catherine E. Rice^45^, Carrie Fassler^29^, Catherine Gray^43^, Chris Gunter^46^, Corrie H. Walston^43^, Cheryl Klaiman^46^, Caroline Leonczyk^13^, Christa Lese Martin^47^, Catherine Lord^17^, Cora M. Taylor^47^, Caitlin McCarthy^38^, Cesar Ochoa-Lubinoff^48^, Crissy Ortiz^38^, Cynthia Pierre^13^, Cordelia R. Rosenberg^33^, Chris Rigby^2^, Casey Roche^38^, Clara Shrier^38^, Chris Smith^34^, Candace Van Wade^38^, Casey White-Lehman^2^, Christopher Zaro^35^, Cindy Zha^24^, Dawn Bentley^14^, Dahriana Correa^24^, Dustin E. Sarver^49^, David Giancarla^24^, David G. Amaral^41^, Dain Howes^28^, Dalia Istephanous^28^, Daniel Lee Coury^26^, Deana Li^41^, Danica Limon^39^, Desi Limpoco^32^, Diamond Phillips^13^, Desiree Rambeck^28^, Daniela Rojas^25^, Diksha Srishyla^28^, Danielle Stamps^49^, Dennis Vasquez Montes^2^, Daniel Cho^50^, Dave Cho^2^, Emily A. Fox^50^, Ethan Bahl^4^, Elizabeth Berry-Kravis^48^, Elizabeth Blank^29^, Erin Bower^34^, Elizabeth Brooks^2^, Eric Courchesne^34^, Emily Dillon^16^, Erin Doyle^38^, Erin Given^26^, Ellen Grimes^15^, Erica Jones^2^, Eric J. Fombonne^32^, Elizabeth Kryszak^26^, Ericka L. Wodka^16^, Elena Lamarche^43^, Erica Lampert^29^, Eric M. Butter^26^, Eirene O’Connor^2^, Edith Ocampo^13^, Elizabeth Orrick^25^, Esmeralda Perez^2^, Elizabeth Ruzzo^5^, Emily Singer^2^, Emily T. Matthews^35^, Ernest V. Pedapati^29^, Faris Fazal^32^, Fiona K. Miller^23^, Gabriella Aberbach^35^, Gabriele Baraghoshi^14^, Gabrielle Duhon^39^, Gregory Hooks^28^, Gregory J. Fischer^18^, Gabriela Marzano^39^, Gregory Schoonover^14^, Gabriel S. Dichter^43^, Gabrielle Tiede^26^, Hannah Cottrell^19^, Hannah E. Kaplan^34^, Haidar Ghina^50^, Hanna Hutter^16^, Hope Koene^21^, Hoa Lam Schneider^24^, Holly Lechniak^13^, Hai Li^48^, Hadley Morotti^32^, Hongjian Qi^31^, Harper Richardson^38^, Hana Zaydens^2^, Haicang Zhang^31^, Haoquan Zhao^31^, Ivette Arriaga^17^, Ivy F. Tso^51^, John Acampado^2^, Jennifer A. Gerdts^50^, Josh Beeson^48^, Jennylyn Brown^2^, Joaquin Comitre^24^, Jeanette Cordova^33^, Jennifer Delaporte^19^, Joseph F. Cubells^45^, Jill F. Harris^42^, Jared Gong^25^, Jaclyn Gunderson^28^, Jessica Hernandez^2^, Jessyca Judge^23^, Jane Jurayj^21^, Julie Manoharan^2^, Jessie Montezuma^38^, Jason Neely^16^, Jessica Orobio^39^, Juhi Pandey^52^, Joseph Piven^43^, Jose Polanco^29^, Jibrielle Polite^2^, Jacob Rosewater^24^, Jessica Scherr^26^, James S. Sutcliffe^53^, James T. McCracken^17^, Jennifer Tjernagel^2^, Jaimie Toroney^2^, Jeremy Veenstra-Vanderweele^54^, Jiayao Wang^31^, Katie Ahlers^50^, Kathryn A. Schweers^13^, Kelli Baalman^39^, Katie Beard^28^, Kristen Callahan^49^, Kendra Coleman^34^, Kate D. Fitzgerald^23^, Kate Dent^47^, Katharine Diehl^2^, Kelsey Gonring^48^, Katherine G. Pawlowski^35^, Kathy Hirst^19^, Kiely Law^2^, Karen L. Pierce^34^, Karla Murillo^17^, Kailey Murray^43^, Kerri Nowell^19^, Kaela O’Brien^29^, Katrina Pama^16^, Kelli Real^43^, Kaitlyn Singer^47^, Kaitlin Smith^41^, Kevin Stephenson^26^, Katherine Tsai^17^, Leonard Abbeduto^41^, Lindsey A. Cartner^2^, Landon Beeson^32^, Laura Carpenter^38^, Lucas Casten^4^, Leigh Coppola^32^, Lisa Cordiero^33^, Lindsey DeMarco^52^, Lillian D. Pacheco ^32^, Lorena Ferreira Corzo^48^, Lisa H. Shulman^36^, Lauren Kasperson Walsh^47^, Laurie Lesher^14^, Lynette M. Herbert^24^, Lisa M. Prock^35^, Lacy Malloch^49^, Lori Mann^2^, Luke P. Grosvenor^2^, Laura Simon^28^, Latha V. Soorya^13^, Lucy Wasserburg^28^, Lisa Yeh^13^, Lark Y. Huang-Storms ^32^, Michael Alessandri^24^, Marc A. Popp^18^, Melissa Baer^48^, Malia Beckwith^42^, Myriam Casseus^42^, Michelle Coughlin^35^, Mary Currin^43^, Michele Cutri^24^, Malcolm D. Mallardi^2^, Megan DuBois^28^, Megan Dunlevy^46^, Martin E. Butler^2^, Margot Frayne^25^, McLeod F. Gwynette^55^, Mohammad Ghaziuddin^23^, Monica Haley^17^, Michelle Heyman^37^, Margaret Hojlo^35^, Michelle Jordy^43^, Michael J. Morrier^45^, Misia Kowanda^2^, Melinda Koza^16^, Marilyn Lopez^42^, Megan McTaggart^16^, Megan Norris^26^, Melissa N. Hale^24^, Molly O’Neil^36^, Madison Printen^13^, Madelyn Rayos^24^, Mahfuza Sabiha^2^, Mustafa Sahin^56^, Marina Sarris^2^, Mojeeb Shir^34^, Matthew Siegel^57^, Morgan Steele^25^, Megan Sweeney^19^, Maira Tafolla^17^, Maria Valicenti-McDermott^36^, Mary Verdi^57^, Megan Y. Dennis^58^, Nicolas Alvarez^16^, Nicole Bardett^15^, Natalie Berger^13^, Norma Calderon^13^, Nickelle Decius^24^, Natalia Gonzalez^42^, Nina Harris^15^, Noah Lawson^2^, Natasha Lillie^28^, Nathan Lo^2^, Nancy Long^26^, Nicole M. Russo-Ponsaran^13^, Natalie Madi^29^, Nicole Mccoy^29^, Natalie Nagpal^2^, Nicki Rodriguez^41^, Nicholas Russell^26^, Neelay Shah^2^, Nicole Takahashi^19^, Nicole Targalia^33^, Olivia Newman^28^, Opal Y. Ousley^45^, Peter Heydemann^48^, Patricia Manning^29^, Paul S. Carbone^14^, Raphael A. Bernier^50^, Rachel A. Gordon^13^, Rebecca C. Shaffer^29^, Robert D. Annett^49^, Renee D. Clark^43^, Roger Jou^21^, Rebecca J. Landa^16^, Rachel K. Earl^50^, Robin Libove^25^, Richard Marini^2^, Ryan N. Doan^40^, Robin P. Goin-Kochel^39^, Rishiraj Rana^2^, Richard Remington^2^, Roman Shikov^16^, Robert T. Schultz^52^, Shelley Aberle^32^, Shelby Birdwell^19^, Sarah Boland^21^, Stephanie Booker^29^, S. Carpenter^15^, Sharmista Chintalapalli^23^, Sarah Conyers^38^, Sophia D’Ambrosi^38^, Sara Eldred^26^, Sunday Francis^28^, Swami Ganesan^2^, Susan Hepburn^33^, Susannah Horner^52^, Samantha Hunter^19^, Stephanie J. Brewster^59^, Soo J. Lee^13^, Suma Jacob^28^, Stanley Jean^2^, So Hyun Kim^60^, Sydney Kramer^4^, Sandra L. Friedman^33^, Sarely Licona^13^, Sandy Littlefield^25^, Stephen M. Kanne^19^,^61^, Sarah Mastel^32^, Sheena Mathai^46^, Sophia Melnyk^29^, Sarah Michaels^16^, Sarah Mohiuddin^23^, Samiza Palmer^52^, Samantha Plate^52^, Shanping Qiu^37^, Shelley Randall^29^, Sophia Sandhu^17^, Susan Santangelo^57^, Swapnil Shah^2^, Steve Skinner^62^, Samantha Thompson^41^, Sabrina Xiao^2^, Sidi Xu^34^, Sabrina White^49^, Stormi White^46^, Tia Chen^34^, Tunisia Greene^2^, Theodore Ho^50^, Teresa Ibanez^26^, Tanner Koomar^4^, Tiziano Pramparo^34^, Tara Rutter^50^, Tamim Shaikh^33^, Taylor Thomas^4^, Thao Tran^48^, Timothy W. Yu^40^, Virginia Galbraith^38^, Vahid Gazestani^63^, Vincent J. Myers^2^, Vaikunt Ranganathan^52^, Vini Singh^16^, William Curtis Weaver^47^, Wenteng CaI^28^, Wubin Chin^2^, Wha S. Yang^17^, YB Choi^60^, Zachary E. Warren^30^

^13^Department of Psychiatry and Behavioral Sciences, Rush University Medical Center, Chicago, Illinois 60612; ^14^Department of Pediatrics, University of Utah, Salt Lake City, Utah 84108; ^15^Vanderbilt Kennedy Center, Vanderbilt University Medical Center, Nashville, Tennessee 37232; ^16^Center for Autism and Related Disorders, Kennedy Krieger Institute, Baltimore, Maryland 21211; ^17^Department of Psychiatry and Biobehavioral Sciences, University of California, Los Angeles, Los Angeles, California 90095; ^18^PreventionGenetics, Marshfield, Wisconsin 54449; ^19^Thompson Center for Autism and Neurodevelopmental Disorders, University of Missouri, Columbia, Missouri 65211; ^20^Department of Pediatrics, University of Minnesota, Minneapolis, Minnesota 55414; ^21^Child Study Center, Yale School of Medicine, New Haven, Connecticut 06519; ^22^Department of Neurogenetics, Kennedy Krieger Institute, Baltimore, Maryland 21205; ^23^Department of Psychiatry, University of Michigan, Ann Arbor, Michigan 48109; ^24^Department of Psychology, University of Miami’s Center for Autism and Related Disabilities (UM-CARD), Coral Gables, Florida 33146; ^25^Department of Psychiatry and Behavioral Sciences, Stanford University, Stanford, California 94305; ^26^Division of Pediatric Psychology and Neuropsychology, Nationwide Children’s Hospital (Child Development Center), Columbus, Ohio 43205; ^27^John P. Hussman Institute for Human Genomics, University of Miami Miller School of Medicine, Miami, Florida 33136; ^28^Department of Psychiatry, University of Minnesota, Minneapolis, Minnesota 55455; ^29^Department of Psychiatry and Behavioral Neuroscience, Cincinnati Children’s Hospital Medical Center - Research Foundation, Cincinnati, Ohio 45229; ^30^Department of Pediatrics, Vanderbilt University Medical Center, Nashville, Tennessee 37232; ^31^Department of Systems Biology, Columbia University Medical Center, New York, NY 10032; ^32^Department of Psychiatry, Oregon Health & Science University, Portland, Oregon 97239; ^33^Department of Pediatrics, JFK Partners/University of Colorado School of Medicine, Aurora, Colorado 80045; ^34^Department of Neurosciences, University of California, San Diego and SARRC Phoenix, La Jolla, California 92037; ^35^Department of Pediatrics, Boston Children’s Hospital, Boston, Massachusetts 02115; ^36^Department of Pediatrics, Montefiore Medical Center and The Albert Einstein College of Medicine, Bronx, New York 10461; ^37^Department of Psychiatry, Weill Cornell Medicine, White Plains, New York 10605; ^38^Department of Pediatrics, Medical University of South Carolina, Charleston, South Carolina 29425; ^39^Department of Pediatrics, Texas Children’s Hospital (Baylor College of Medicine), Houston, Texas 77030; ^40^Department of Medicine, Boston Children’s Hospital, Boston, Massachusetts 02115; ^41^MIND Institute and Department of Psychiatry and Behavioral Sciences, University of California, Davis, Sacramento, California 95817; ^42^Children’s Specialized Hospital, Toms River, New Jersey 08755; ^43^Department of Psychiatry, University of North Carolina (UNC, TEACCH, CIDD), Chapel Hill, North Carolina 27599; ^44^Department of Molecular and Medical Genetics, Oregon Health & Science University, Portland, Oregon 97239; ^45^Department of Psychiatry and Behavioral Sciences, Emory University and Marcus Autism Center, Atlanta, Georgia 30033; ^46^Department of Pediatrics, Emory University and Marcus Autism Center, Atlanta, Georgia 30329; ^47^Geisinger Autism & Developmental Medicine Institute, Lewisburg, Pennsylvania 17837; ^48^Department of Pediatrics, Rush University Medical Center, Chicago, Illinois 60612; ^49^Department of Pediatrics, University of Mississippi Medical Center, Jackson, Mississippi 39110; ^50^Department of Psychiatry and Behavioral Sciences, University of Washington/Seattle Children’s Autism Center, Seattle, Washington 98195; ^51^Department of Psychology, University of Michigan, Ann Arbor, Michigan 48109; ^52^Center for Autism Research, Children’s Hospital of Philadelphia, Philadelphia, Pennsylvania 19146; ^53^Department of Molecular Physiology and Biophysics, Vanderbilt University, Nashville, Tennessee 37232; ^54^Department of Psychiatry, Columbia University Medical Center, New York, NY 10032; ^55^Department of Psychiatry and Behavioral Sciences, Medical University of South Carolina, Charleston, South Carolina 29425; ^56^Department of Neurology, Boston Children’s Hospital, Boston, Massachusetts 02115; ^57^Maine Medical Center Research Institute, Scarborough, Maine 04074; ^58^Genome Center, MIND Institute, Department of Biochemistry and Molecular Medicine, University of California, Davis, Sacramento, California 95616; ^59^Translational Neuroscience Center, Boston Children’s Hospital, Boston, Massachusetts 02115; ^60^Center for Autism and the Developing Brain (CADB), Weill Cornell Medicine, White Plains, New York 10605; ^61^Department of Health Psychology, University of Missouri, Columbia, Missouri 65211; ^62^Greenwood Genetic Center, Greenwood, South Carolina 29646; ^63^Department of Pediatrics, University of California, San Diego and SARRC Phoenix, La Jolla, California 92037

