## Supplementary Material for "Integrating *de novo* and inherited variants in over 42,607 autism cases identifies mutations in new moderate risk genes"

### Supplemental Contents

**Supplementary Note** 4

*Variant filtering on rare inherited LoFs* 4

Supplementary Figure S30: Workflow for variant filtering on rare inherited LoFs 5

**Supplementary Table Legends** 6

**Supplementary Figure S1: Overall burden of *de novo* variants in four ASD cohorts included in the discovery sample** 9

**Supplementary Figure S2: Transmission disequilibrium of LoF variants of chromosome X genes** 10

**Supplementary Figure S3: Transmission disequilibrium of deleterious missense (D-mis) variants** 12

**Supplementary Figure S4: Number of genes in each gene set and pairwise overlaps between gene sets before (A) and after (B) excluding known ASD/NDD genes** 15

**Supplementary Figure S5: Association of *de novo* damaging variants and rare, inherited LoFs with cognitive impairment in ASD cases** 17

**Supplementary Figure S6: Inherited LoF variants in genes prioritized by A-risk are not associated with phenotype severity in cases** 19

**Supplementary Figure S7: Comparing carrier rates of LoFs between pseudo-controls and three panels of population-based references for genes selected for replication** 21

**Supplementary Figure S8: Empirical relationship between haploid LoF mutation rate, cumulative allele frequency (CAF) of HC LoFs, and fraction of de novo LoFs in ASD cases** 23

**Supplementary Figure S9: In genes selected for replication, most LoFs are ultra-rare** 25

**Supplementary Figure S10: Comparison on carrier rates of ultra-rare LoFs between European and non-European samples in gnomAD exomes and gnomAD genomes** 27

**Supplementary Figure S11: Comparing the high confidence LoF rate in 31,976 unrelated ASD cases with gnomAD exomes and TopMed** 29

**Supplementary Figure S12: Expression signatures of new ASD genes** 31

**Supplementary Figure S13: Transmission disequilibrium of exonic or single gene deletions of *ITSN1* and *NAV3*** 33

**Supplementary Figure S14: Calculated cognitive impairment and sex ratio in individuals with ASD in SPARK** 36

**Supplementary Figure S15: Distribution of different types of LoF variants in known ASD genes enriched by de novo variants (DNVs) and comparison with population controls** 37

**Supplementary Figure S16: Empirical relationship between estimated relative risk to ASD and estimated selection coefficient** 39

**Supplementary Figure S17: Cumulative distribution of haploid mutation rates of LoF and D-mis variants of all protein coding genes on autosomes** 41

**Supplementary Figure S18: Burden of de novo and inherited LoFs in genes with high and low LoF mutation rates** 43

**Supplementary Figure S19: Power of case-control association by rare LoFs variants with sample size equal to current study** 46

**Supplementary Figure S20: Sample sizes required for achieving 90% of power** 48

**Supplementary Figure S21: SPARK sample QC: relatedness check and sex validation** 49

**Supplementary Figure S22: SPARK principal component analysis (PCA) and ancestry inference** 51

**Supplementary Figure S23: SPARK self-reported cognitive impairment shows stronger correlation with Vineland score than full-scale IQ** 53

**Supplementary Figure S24: Comparing phenotypes of samples from simplex and multiplex families in SPARK cohort** 54

**Supplementary Figure S25: Summary of final DNV call sets for SPARK and SSC discovery samples** 56

**Supplementary Figure S26: Evidence of post-zygotic mosaicisms in the final DNV call set** 57

**Supplementary Figure S27: Rare variant workflow and QC strategy** 58

**Supplementary Figure S28: Comparison of inhouse DenovoWEST results on NDD trios with published results** 59

**Supplementary Figure S29: Illustration of pseudo cases and contributing sample sizes in different types of pedigrees** 61

**Reference** 62

### Supplementary Note

#### *Variant filtering on rare inherited LoFs*

Standing LoFs are notoriously fraught with variant calling artefacts, low confidence LoFs that escape nonsense mediated decay or do not affect splicing[1-3], or LoFs that only affect transcripts that have low expression in disease relevant tissues[4]. In constrained genes, we found about 6% QC passed variant calls initially annotated as LoFs are part of non-LoF MNV or frame-restoring indels, in contrast to <1% in dnLoFs (**Supplementary Figure S26A**). To prioritize high confidence standing LoFs, we applied of LOFTEE/pExt and allele frequency filters. Using pExt>=0.1 in developing brain removes more than 1/3 of LoFs without changing the over-transmission rate to affected offspring. Further applying ultra-rare allele frequency filter (allele frequency<1.5e-4 or singleton in cohort and <5e-5 in populations) removes additional 11% standing LoFs with minimal changes to over-transmission rate. Together, close to half of standing LoF variants are removed that does not contribute to ASD in offspring. Although further increasing pExt threshold to 0.9 will reduce over-transmission rate, it is likely that optimal pExt threshold is gene-specific and we may underestimate fraction of standing LoFs that does not contribute to ASD. In comparison, the same set of filters only removes 3% dnLoFs and 5% dnDmis in ASD, 3% dnLoFs and 2% dnDmis in other NDD with minimal changes to rate difference between affected and control trios (**Supplementary Figure S26B**, **Figure 2A**). LoFs in pseudo cases is a mixture of de novo and inherited LoFs, and as expected 25% of them are removed by the same filters which do not change rate difference between cases and pseudo controls (**Figure 2B**). We used ultra-rare high confidence (pExt>=0.1) standing LoFs in transmission analysis.

*Supplementary Figure S30: Workflow for variant filtering on rare inherited LoFs*

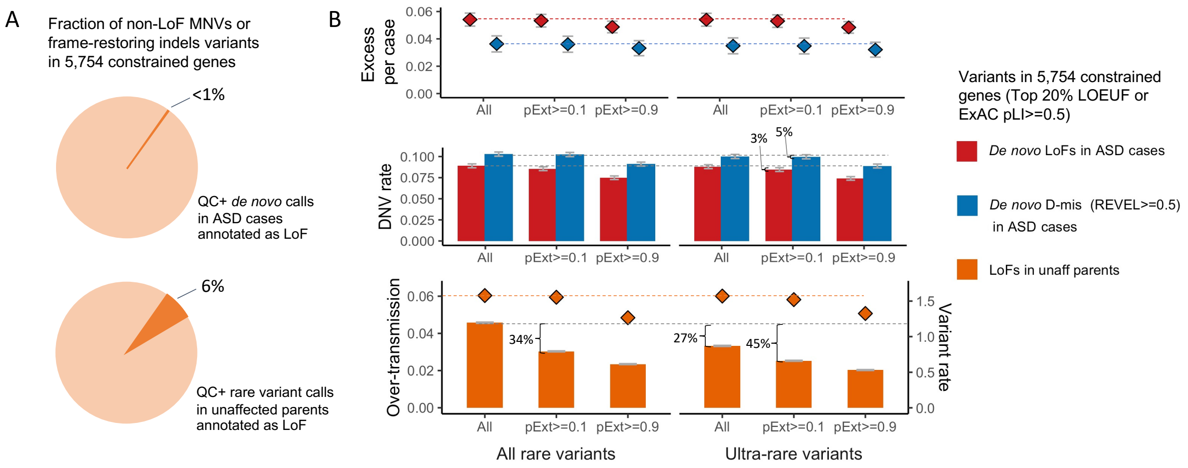


Among QC passed rare variant calls that were initially annotated as LoFs in 5,754 constrained genes (ExAC pLI>=0.5 or top 20% LOEUF), 6% of standing LoFs in 20,491 unaffected parents are part of non-LoF multi-nucleotide variations (MNVs) or frame-restoring indels. In contrast, we observed less than 1% such variants in *de novo* calls.

#### Supplementary Table Legends

Table S1. Cohorts and number of trios included in de novo analysis.

Table S2. Full list of 618 known dominant or X-linked ASD or NDD genes.

Table S3. Genes with p-value < 0.001 in DeNovoWEST analysis of de novo variants in the Stage 1 data. The table summarizes numbers of different types of de novo variants in each gene and results from DenovoWEST test in 16,877 ASD trios.

Table S4. Number of unaffected parents and offspring in trios and duos used in transmission disequilibrium analysis.

Table S5. Gene sets memberships. Gene set enrichment analyses for de novo and rare, inherited LoFs used 5,754 constrained genes (gnomAD LOEUF top 20% or ExACpLI >=0.5) as background. Their memberships in 25 gene sets of 5 categories are listed in this table.

Table S6. Transmission disequilibrium test (TDT) of rare, inherited LoFs in the discovery cohort. For each autosomal gene, rare, inherited LoFs that passed different allele frequency and pExt filters were identified 20,491 unaffected parents and evaluated over-transmission to ASD offspring in 9,504 trios and 2,966 duos. And for each non-PAR chrX gene, LoFs were identified in 11,354 unaffected mothers and evaluated over-transmission to affected sons in 9,883 duos. A total of 260 genes were prioritized for replication (15 overlap with top de novo enriched genes).

Table S7. Effective number of cases and controls in case-control comparison.

Table S8. Estimated average relative risk of rare LoFs of selected autosomal genes.

Table S9. Phenotypic information for individuals with HC LoFs in novel, exome-wide significant ASD risk genes and individuals with HC LOFs in 5 well-established ASD risk genes. 1 means yes, condition is present. 0 means no, condition is not present. NA means the information is not known.

Table S10. Enrichments of STRING clusters for each archetype. Each STRING cluster (a binary label across 1,776 genes in our embedding space) was predicted as a function of the six archetype scores we derived. The significance of each model parameter was assessed and the corresponding p-value is reported if the coefficient was positive (enrichment) or set to 1 if the coefficient was negative.

Table S11. Software tools and their parameter settings used in data processing.

Table S12. Gene-specific pExt thresholds for 96 de novo LoF (dnLoF) enriched genes. We selected 96 known or DenovoWEST exome-wide significant genes that are in the top 30% of gnomAD LOEUF scores and have more than 4 dnLoFs in 23,053 ASD trios. For each gene, a gene-specific pExt threshold was selected from {0.1, 0.5, 0.9} such that all dnLoFs with pExt>=0.1 were be retained. For selected genes that were manually reviewed, we further removed curated non-LoF variants in gnomAD. (A) Compared with the baseline filter of pExt>=0.1, applying gene-specific pExt thresholds and removing curated non-LoFs further filtered out 19% of rare, inherited LoFs in selected genes that have minimal contribution to transmission disequilibrium to affected offspring in 15,603 trios and 4,925 duos. (B) Those filters also further removed an additional 3~4% LoFs in cases after the baseline filter pExt>=0.1, while retaining similar LoF rate differences compared to pseudo-controls. (C) Gene specific pExt thresholds for 96 selected genes.

Tab S13. Burden of de novo variants (DNVs). Burden of DNVs were evaluated by comparing observed with expected rates calculated from baseline mutation rates. The table summarizes burdens of different types of DNVs in constrained (pLI>=0.5) and non-constrained genes (pLI<0.5) and the corresponding positive predictive values that are used as weights in DenovoWEST.

#### Supplementary Figure S1: Overall burden of *de novo* variants in four ASD cohorts included in the discovery sample

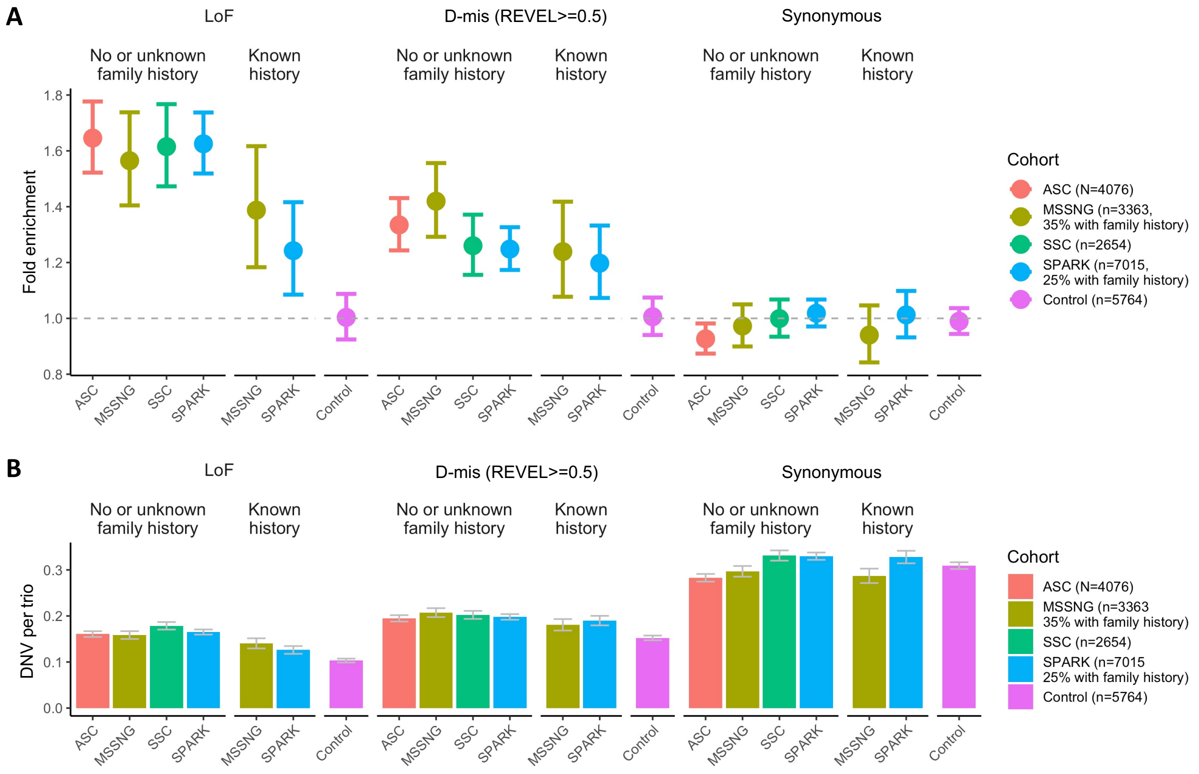


(A) Observed rates of *de novo* LoF, Dmis (REVEL>=0.5) and silent variants (B) are compared with expected rates. We used a 7mer sequence context dependent mutation rate model[5] to calculate expected rates for different classes of *de novo* variants after adjusting sequencing coverage, and found a close match with observed *de novo* rates in control trios. The rates of *de novo* LoF and Dmis variants in ASD cases are significantly higher than baseline expectation and are reduced in cases with known family history.

#### Supplementary Figure S2: Transmission disequilibrium of LoF variants of chromosome X genes

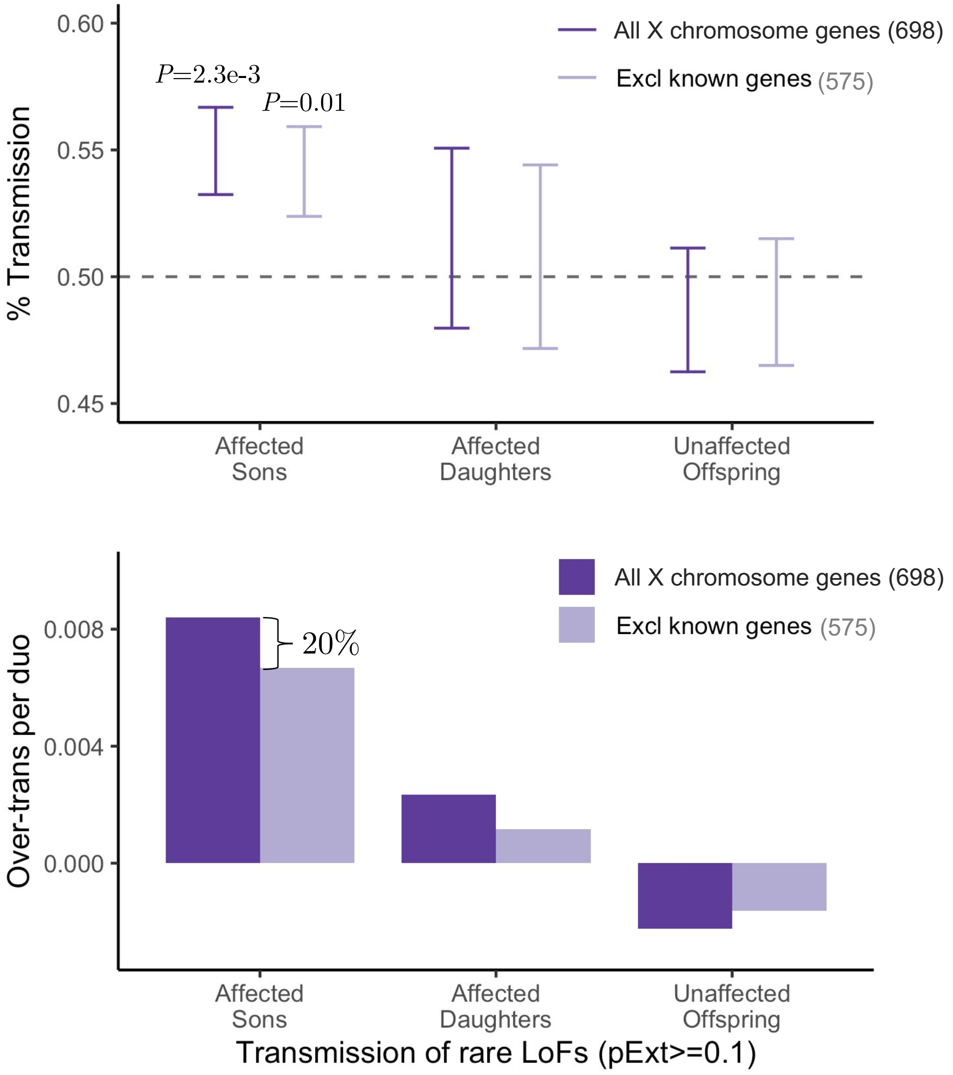


The burden of inherited LoFs on non-PAR part of chrX was evaluated by analyzing the transmission disequilibrium of rare (cohort AF<0.001 and population AF<2e-4) rare, inherited LoFs (pExt>=0.1) identified unaffected mothers. Across all chrX genes, only the proportion of transmission to affected son are significantly above 50% and remain so after excluding known ASD/NDD genes. We observed no significant over-transmission of chrX LoFs to affected daughter or unaffected offspring.

### Supplementary Figure S3: Transmission disequilibrium of deleterious missense (D-mis) variants

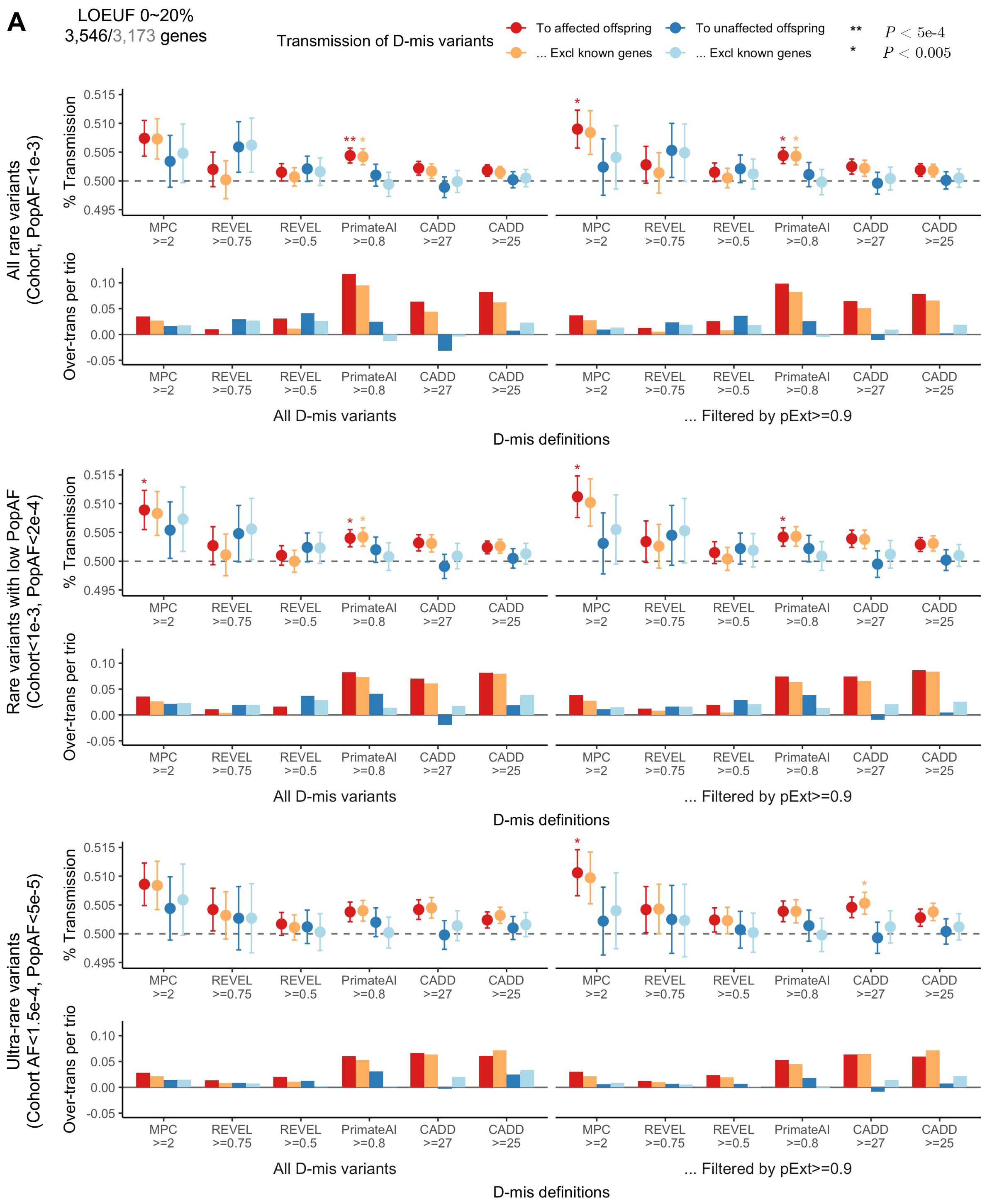

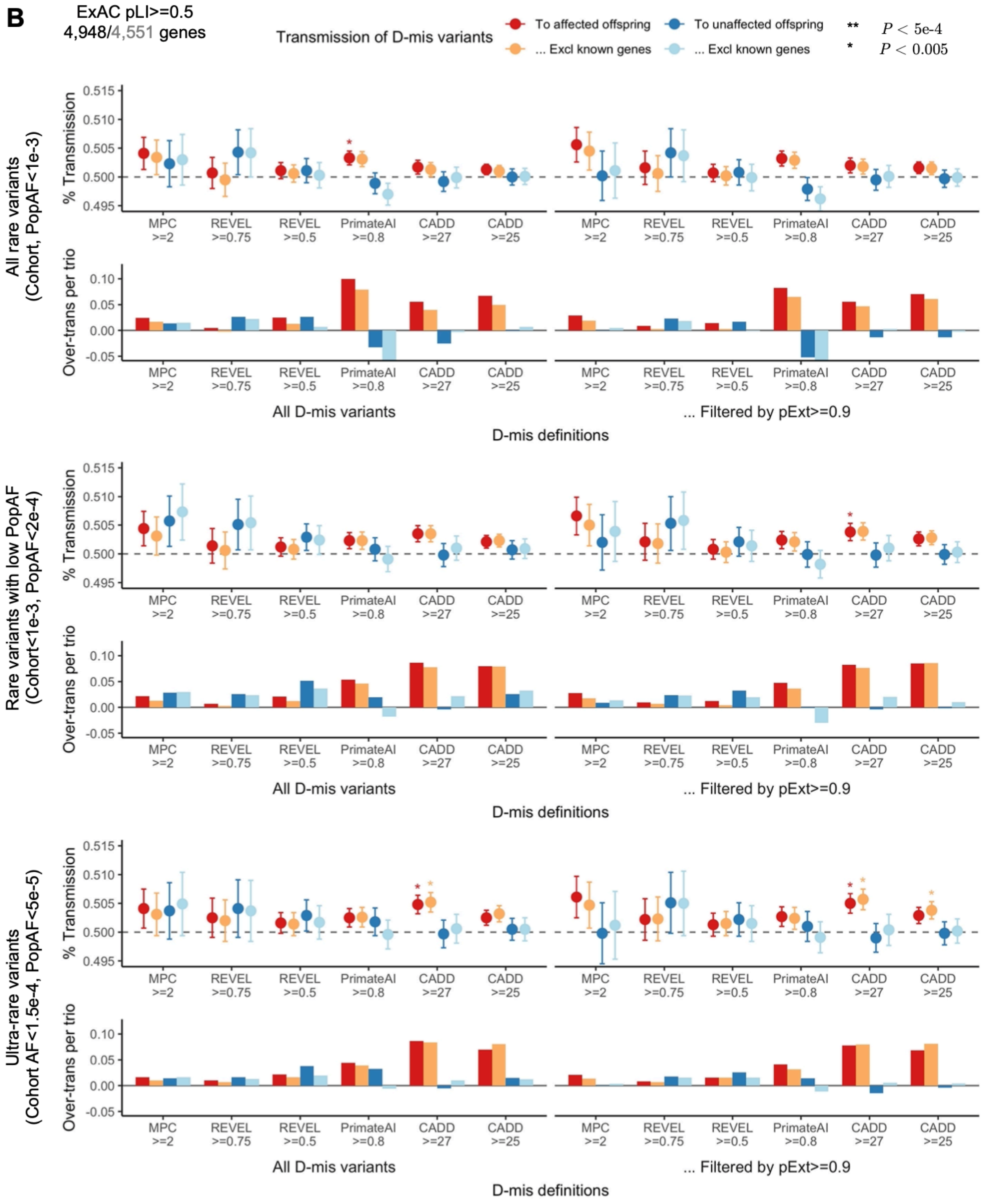


The transmission disequilibrium signals of rare, inherited D-mis variants are generally weaker than LoFs and sensitive to the definition of D-mis and the choice of gene set. For example, in genes of top 20% gnomAD LOEUF (A), Significant over-transmission to affected offspring was observed for rare, inherited D-mis variants defined by MPC>=2, especially those that are further filtered by pExt>=0.9 (P<0.005). PrimateAI>=0.8 prioritized more than two times D-mis variants than MPC>=2 and show significant over-transmission to affected but with lower magnitude than MPC>=2. As a comparison in constrained genes with ExAC pLI>=0.5 (B), over-transmission of D-mis variants defined by MPC>=2 become non-significant. The ultra-rare inherited D-mis defined by CADD>=27 shows strong evidence of over-transmission. Most significant transmission disequilibrium signals remain significant after removing known ASD/NDD genes.

### Supplementary Figure S4: Number of genes in each gene set and pairwise overlaps between gene sets before (A) and after (B) excluding known ASD/NDD genes

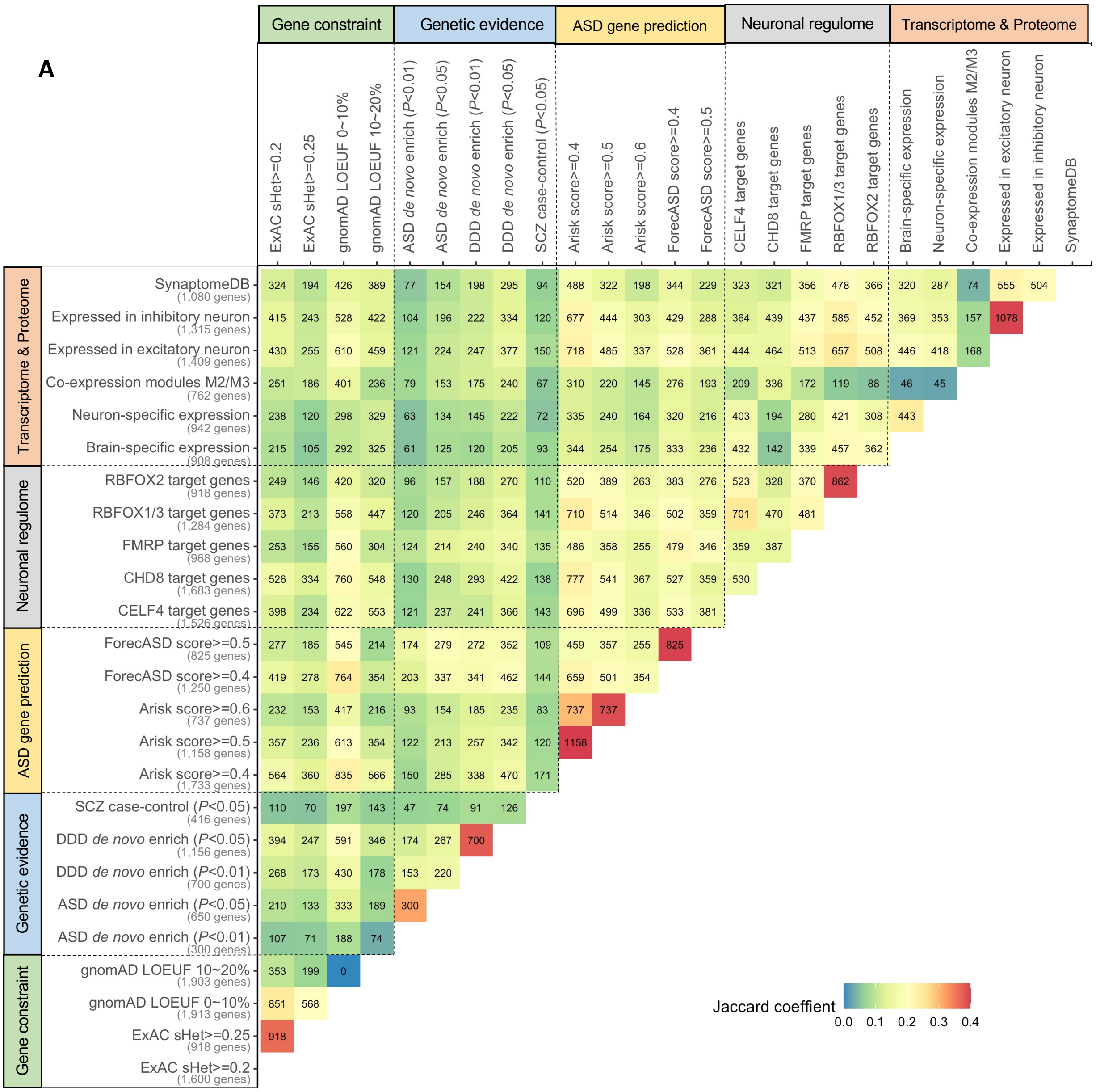

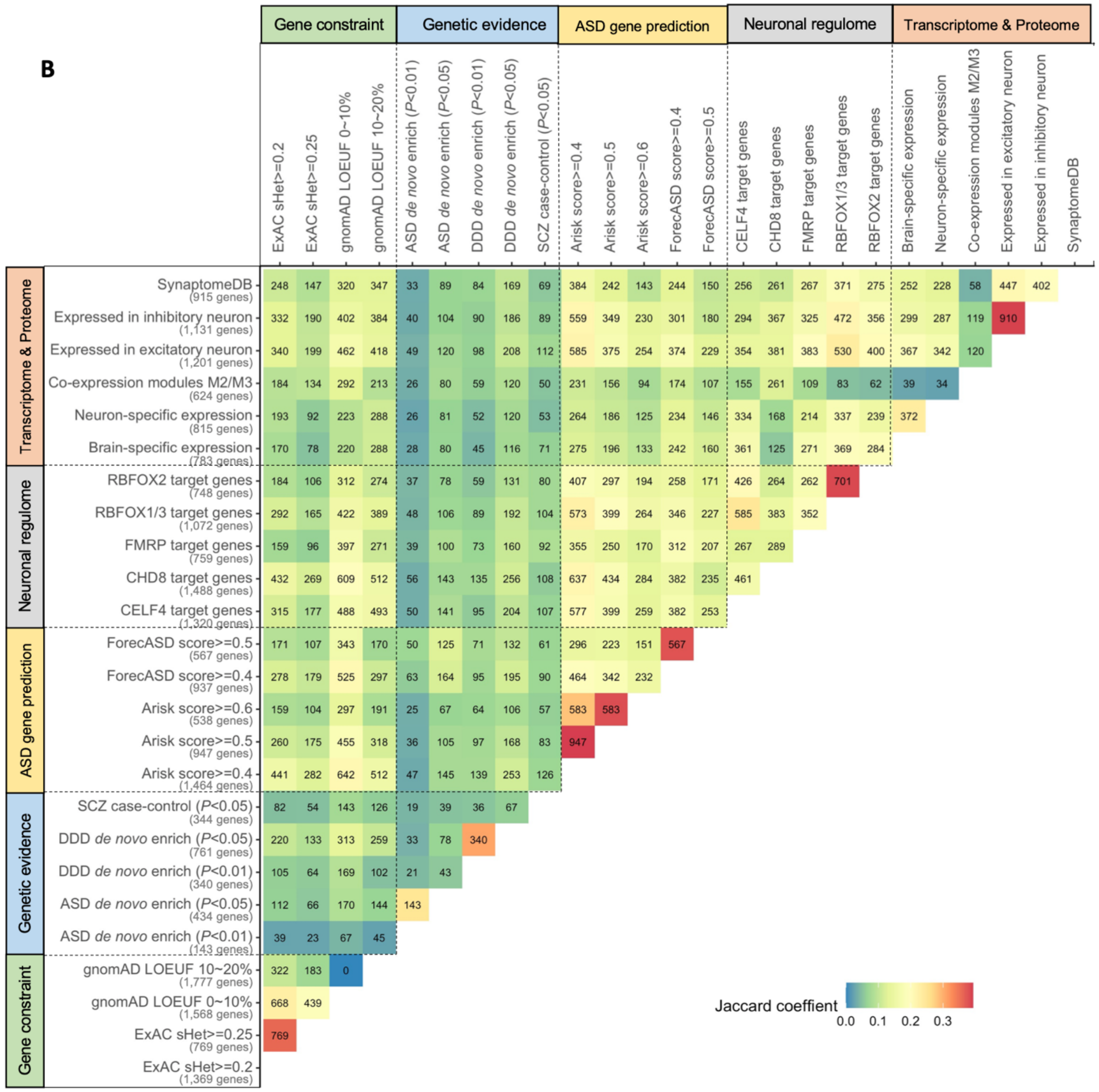


Similarities between gene sets are measured and visualized by Jaccard coefficients.

### Supplementary Figure S5: Association of *de novo* damaging variants and rare, inherited LoFs with cognitive impairment in ASD cases

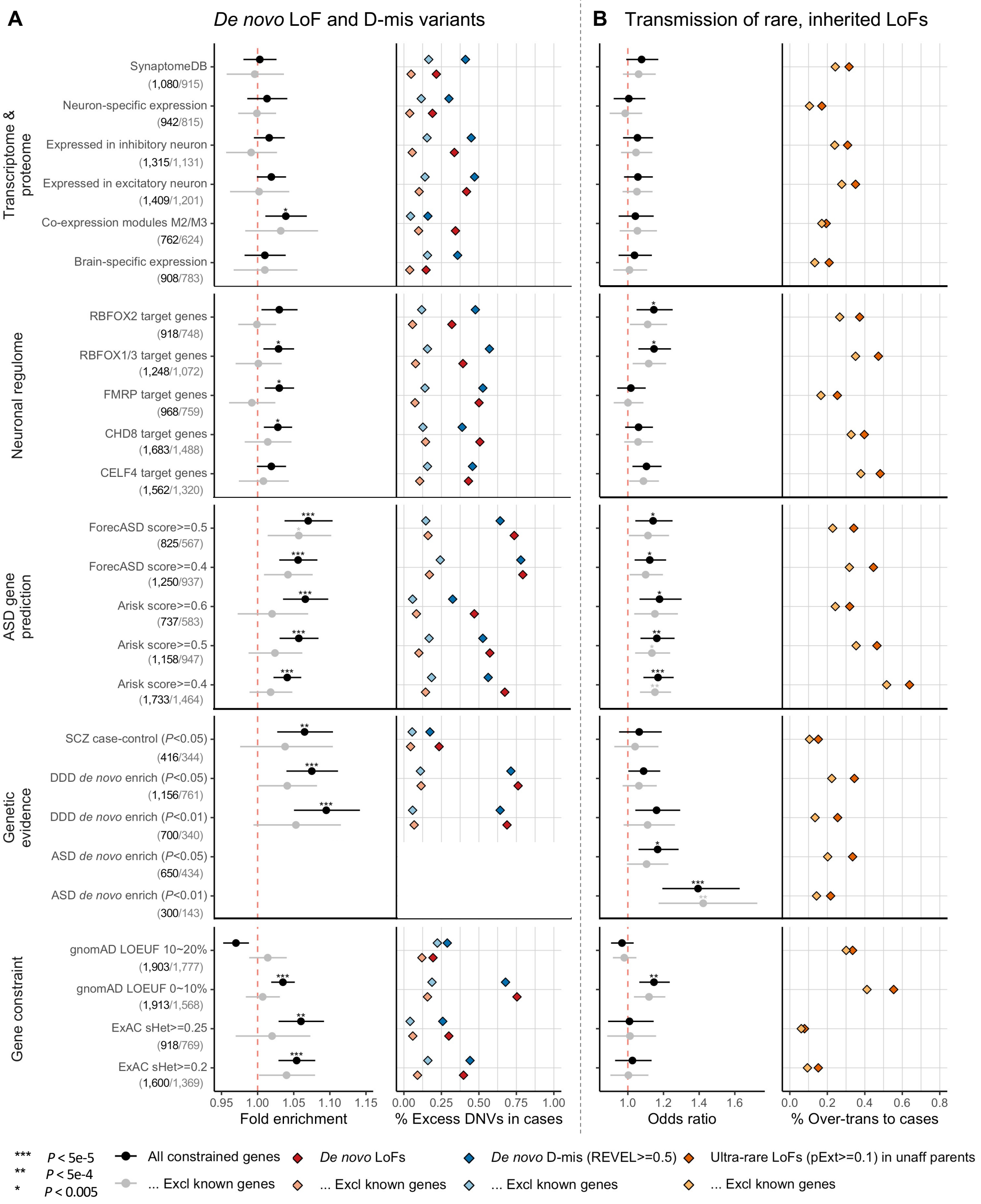

(A) De novo LoF and D-mis variants in genes within the top 10% of LOEUF scores in gnomAD show higher rate and rate ratios between unaffected trios ASD cases with cognitive impairment (Cog Imp) than ASD cases without (No Cog Imp). The increased burden of de novo damaging variants in cases associated with cognitive impairment can be explained by known ASD/NDD genes. De novo damaging variants in other constrained genes are not associated with phenotypic severity. (B) Similarly, ultra-rare inherited LoFs with pExt>=0.1 in genes with the top 10% gnomAD LOEUF scores also show a higher proportion of transmission and a higher over-transmission rate to ASD offspring with cognitive impairment than those without. Rare LoFs in other constrained genes are not significantly associated with phenotypic severity. The increased burden of inherited LoFs in cases with cognitive impairment remains significant after removing known ASD/NDD genes

#### Supplementary Figure S6: Inherited LoF variants in genes prioritized by A-risk are not associated with phenotype severity in cases

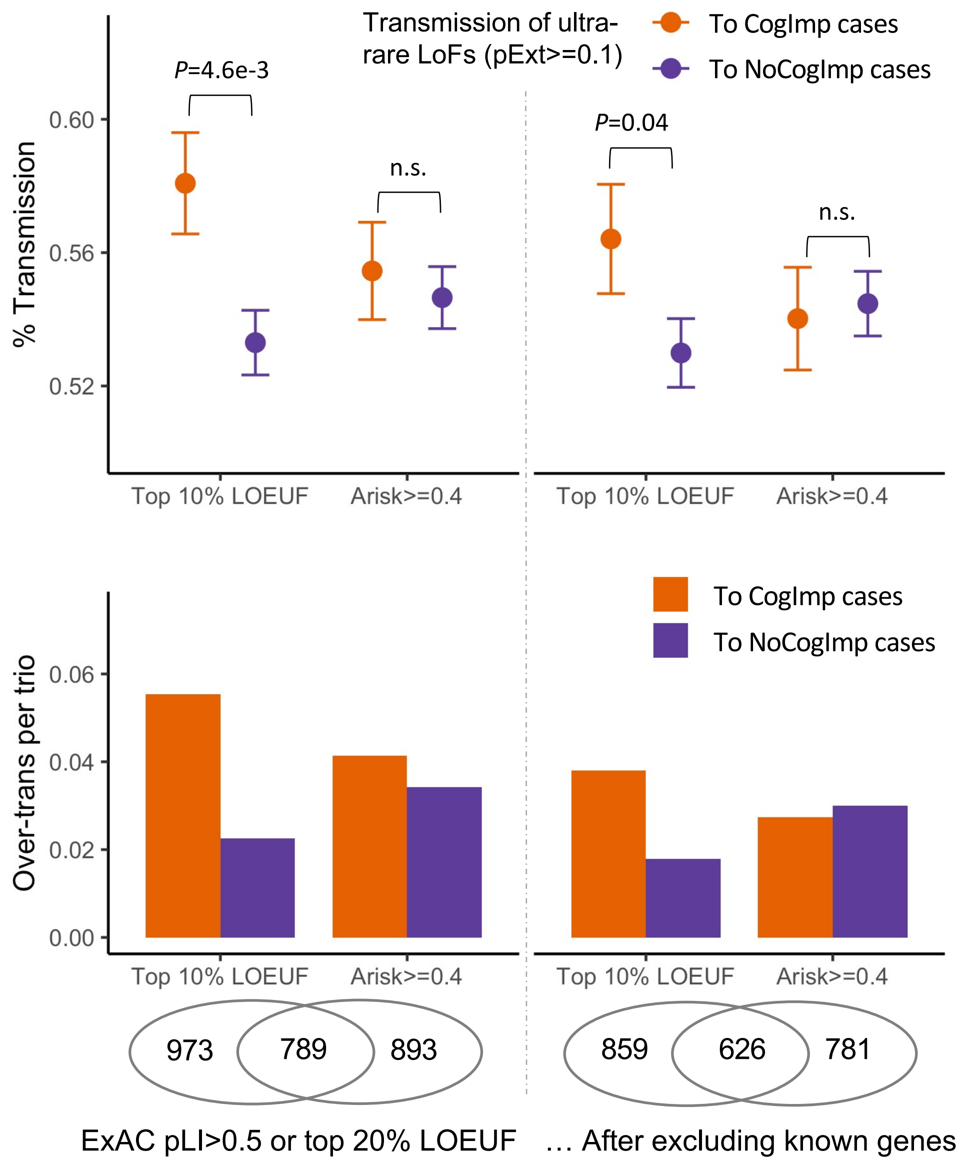


Transmission disequilibrium signals are significantly enriched in genes at top 10% gnomAD LOEUF metrics or having A-risk score>=0.4 (Supplementary Figure S5).

Number of autosomal genes in each set are shown as Venn diagram below the plot. Despite of over 700 overlapping genes (over 600 after removing known genes) in the sets, ultra-rare LoFs with pExt>=0.1 in genes at top 10% gnomAD LOEUF shows significantly higher proportion of transmission to with cognitive impairment ASD cases, whereas those in genes with A-risk>=0.4 show similar proportion of transmission to cases with or without cognitive impairment.

### Supplementary Figure S7: Comparing carrier rates of LoFs between pseudo-controls and three panels of population-based references for genes selected for replication

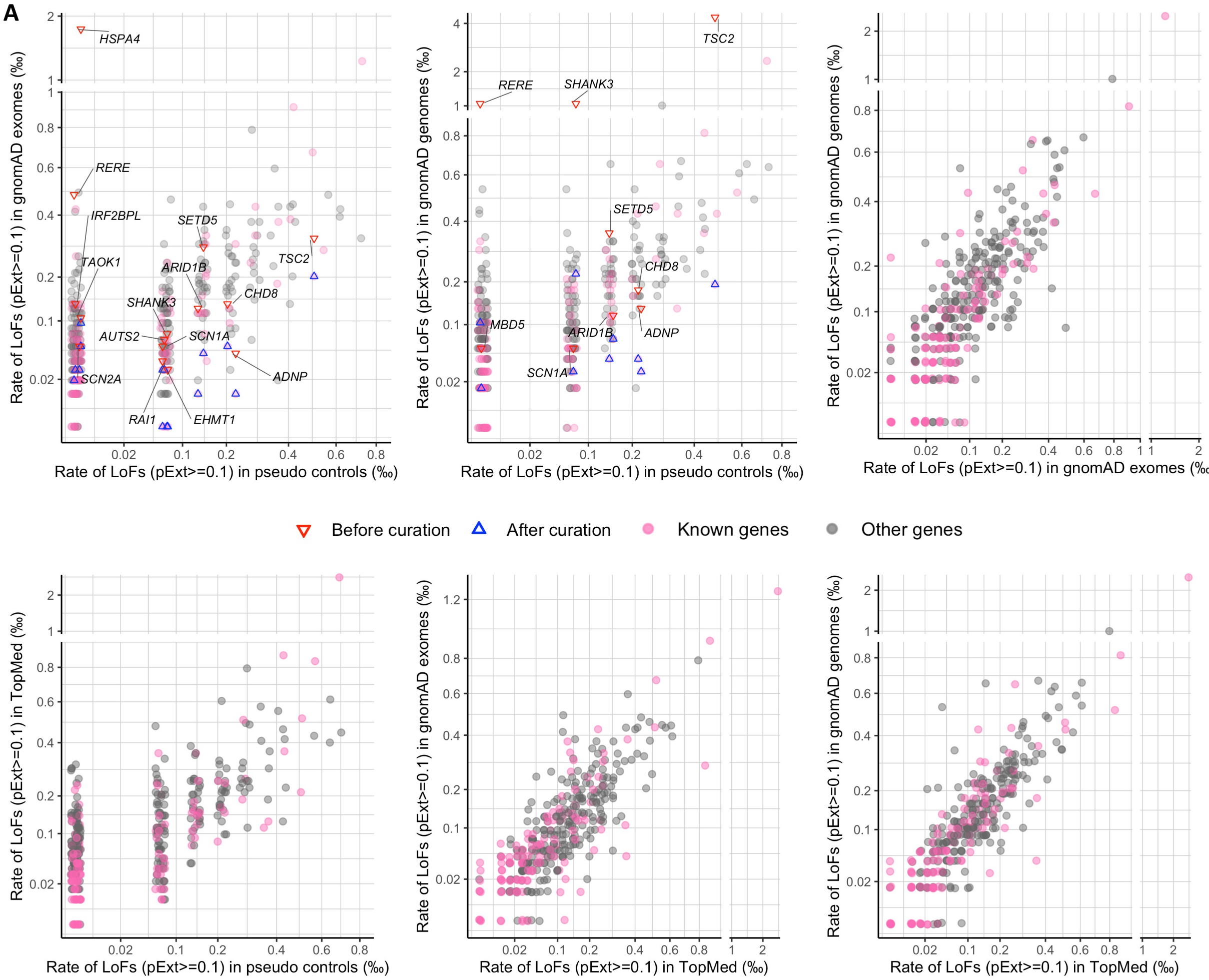

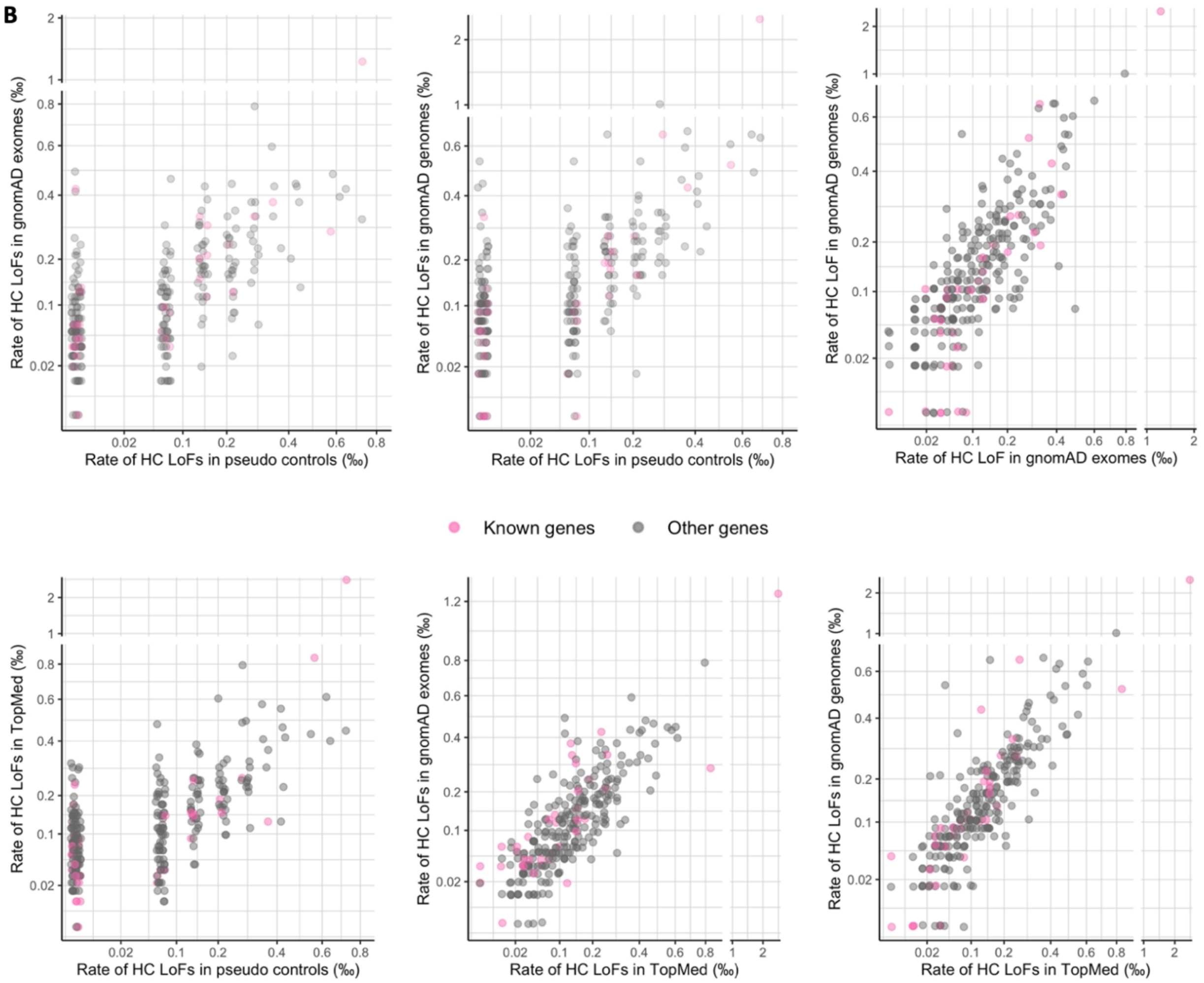


Prioritized gene in top 30% gnomAD LOEUF were used in meta and mega analysis; shown in this plot are 367 autosomal genes. Carrier rates are estimated from 14,128 unrelated pseudo-controls, 104,068 gnomAD exome samples (non-neuro subset), 67,442 gnomAD genome samples (non-neuro subset), and 132,345 TopMed samples. (A) LoFs were filtered by pExt>=0.1. Selected genes whose carrier frequencies change by over 1/3 in gnomAD after manual curation are highlighted. (B) LoFs in de novo LoF enriched genes were further filtered by gene-specific pExt thresholds.

**Supplementary Figure S8: Empirical relationship between haploid LoF mutation rate, cumulative allele frequency (CAF) of HC LoFs, and fraction of de novo LoFs in ASD cases**

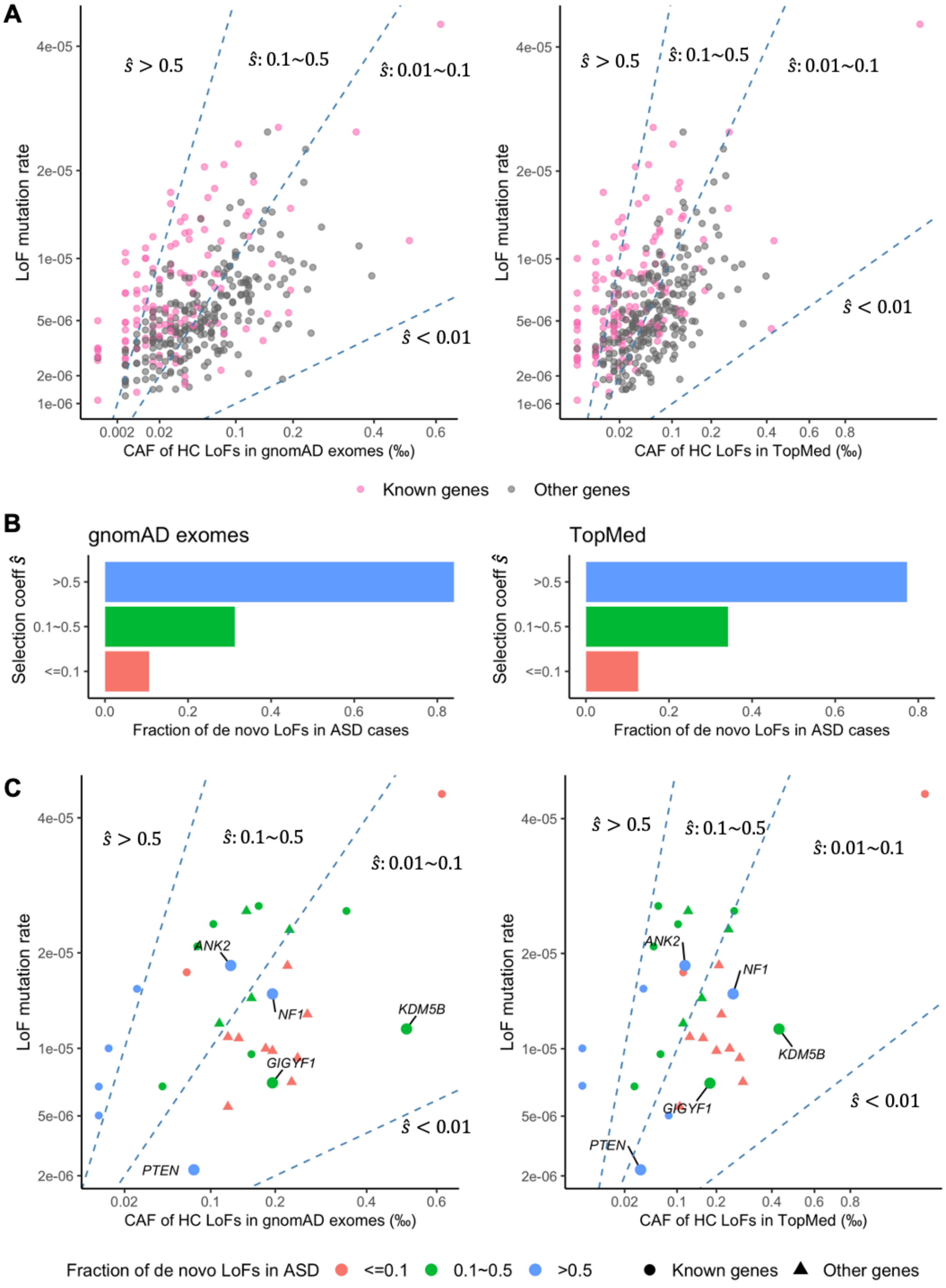

Selection coefficient can be estimated as the ratio of mutation rate over CAF ($\hat{s} = \mu/\hat{f}$). (A) Comparing haploid LoF mutation rate with CAFs of HC LoFs in populations on 367 autosomal genes selected for replication and among top 30% gnomAD LOEUF. CAFs in populations are estimated from gnomAD exomes (125,748 individuals), gnomAD genomes (76,156 individuals), and TopMed (132,345 individuals). Three dashed lines in each plot demarcate the quadrant into areas different estimated selection coefficient ($\hat{s}$: <0.01, 0.01~0.1, 0.1~0.5, and >0.5). Almost all genes have $\hat{s}$>0.01 and known ASD/NDD genes tend to have higher $\hat{s}$. (B) We grouped genes into three bins of $\hat{s}$ (0.01~0.1, 0.1~0.5, and >0.5) and tallied the number of de novo and inherited LoFs in 32,024 unrelated ASD cases. Genes in higher $\hat{s}$ bin have higher proportion of LoFs that are de novo in cases. (C) The same as (A) but only show 30 genes with more than 10 LoFs with inheritance information in unrelated ADS cases. Gene are color coded by the observed proportion of de novo LoFs. There is a general concordance between $\hat{s}$ and fraction of de novo LoFs in these genes. Five genes that also harbor de novo LoFs in control trios (Supplementary Tab 4) are highlighted. These genes have likely under-estimated LoF mutation rates and shows higher proportion of de novo LoFs than $\hat{s}$ estimated from presumed mutation rates.

### Supplementary Figure S9: In genes selected for replication, most LoFs are ultra-rare

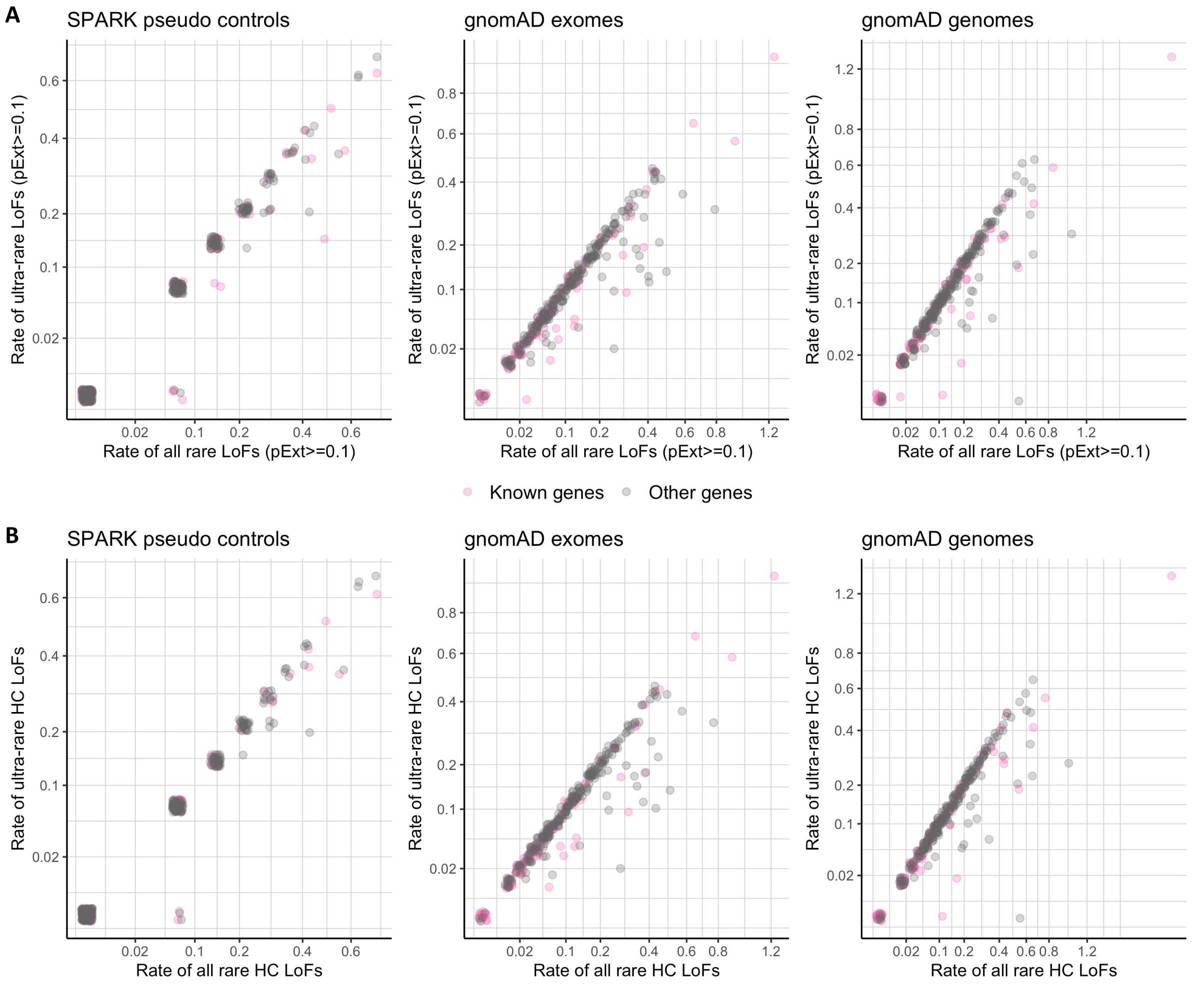


Prioritized gene in top 30% gnomAD LOEUF were used in meta and mega analysis, of which 367 are on autosomes and shown in the plot. We compared carrier rates of all LoFs versus ultra-rare LoFs observed in 14,128 SPARK pseudo controls, 104,068 gnomAD exomes (non-neuro subset), and 67,442 gnomAD genomes (non-neuro subset). Ultra-rare variants are defined by cohort allele frequency <1.5e-4 (or singleton in the cohort) and population allele frequency <5e-5. Most genes are shown along or close the diagonal line, suggesting that most LoFs in those genes are ultra-rare and originated from recent mutational events. (A) LoFs were filtered by pExt>=0.1. (B) LoFs in de novo LoF enriched genes were further filtered by gene-specific pExt thresholds.

#### Supplementary Figure S10: Comparison on carrier rates of ultra-rare LoFs between European and non-European samples in gnomAD exomes and gnomAD genomes

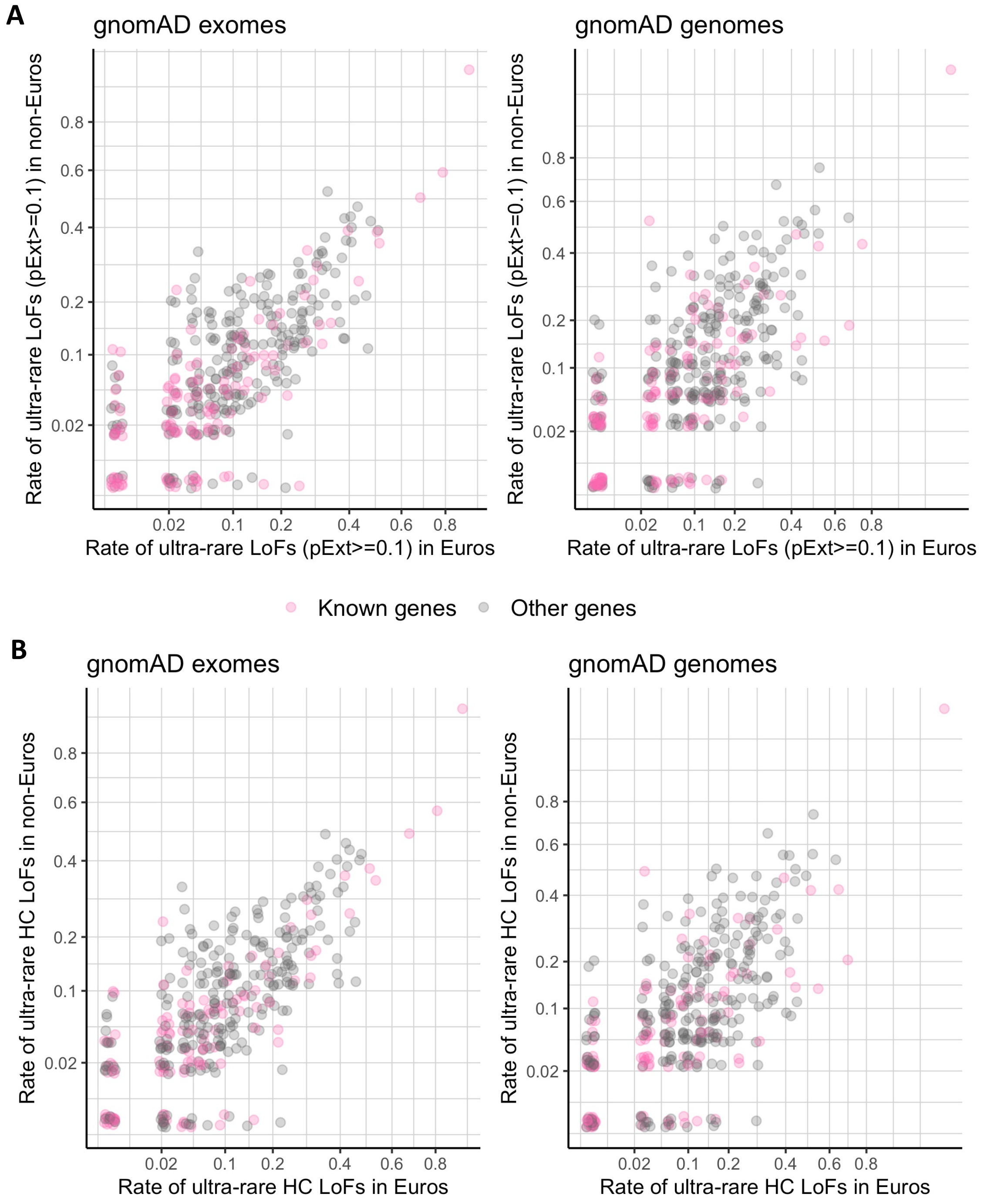

For the same 367 genes shown in Supplementary Figure S9, we compared carrier rates of ultra-rare LoFs between European and non-European samples in gnomAD exomes (44,779 Europeans, 59,289 non-Europeans) and gnomAD genomes (31,966 Europeans, 35,476 non-Europeans). Carrier rates of ultra-rare LoFs in European and non-European population samples are higher correlated, consistent with their recent mutational origin and insensitive to population demographic history. (A) LoFs were filtered by pExt>=0.1. (B) LoFs in de novo LoF enriched genes were further filtered by gene-specific pExt thresholds

### Supplementary Figure S11: Comparing the high confidence LoF rate in 31,976 unrelated ASD cases with gnomAD exomes and TopMed

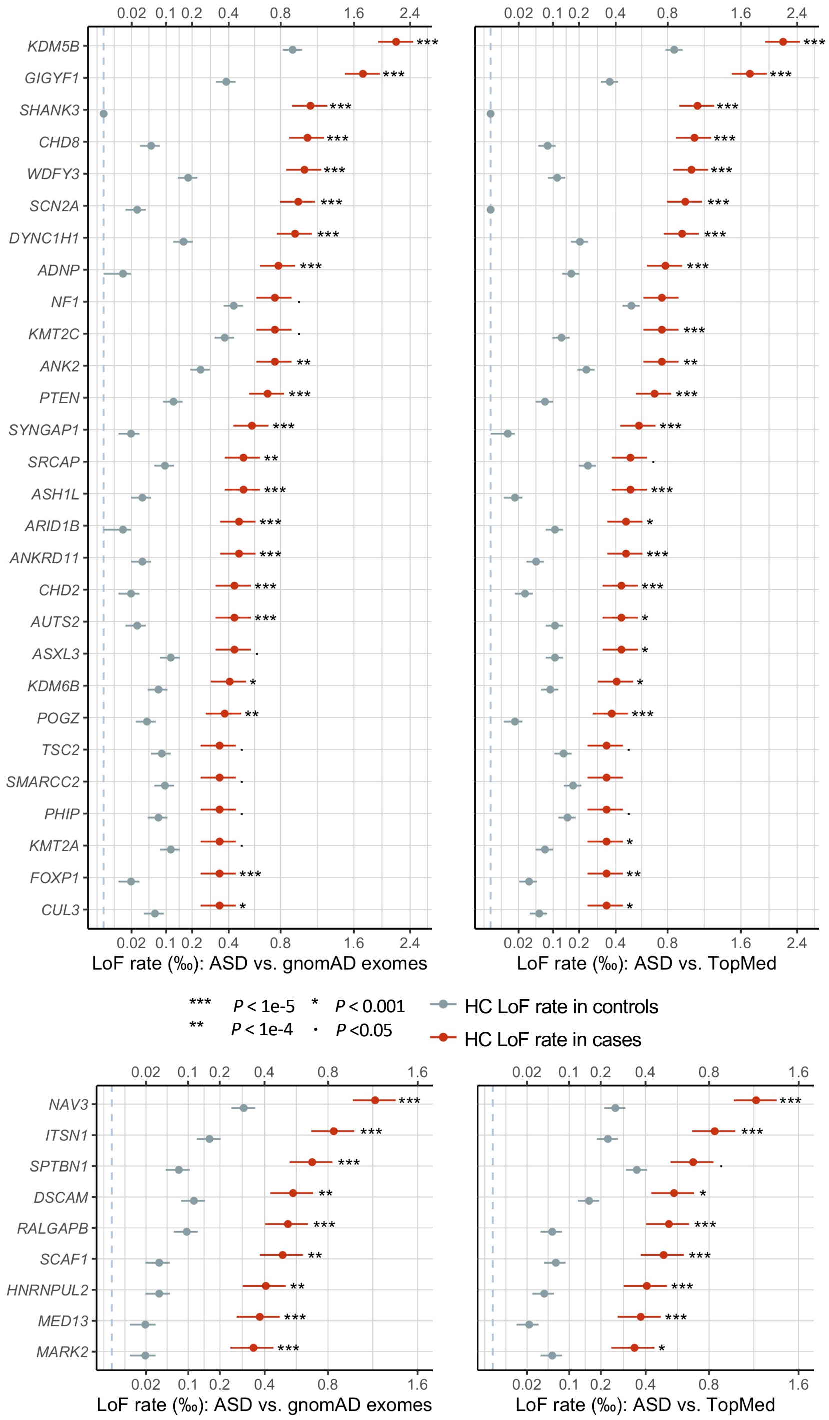

Horizontal bars indicate standard errors. (A) The upper panel shows 28 known ASD/NDD genes in which LOEUF scores are in the top 30% of gnomAD, have a p-value for enrichment among all DNVs (p <9e-6) in 23,039 ASD trios, and have more than 10 LoFs. (B) The lower panel shows 9 additional ASD risk genes that achieved a p-value of <9e-6 in Stage 2 of this analysis. The majority of genes in the lower panels harbor more inherited LoFs than de novo variants. All five novel genes (Table 1) are shown in the lower panel. Note that the x-axes of LoF rates are in the squared root scale.

### Supplementary Figure S12: Expression signatures of new ASD genes

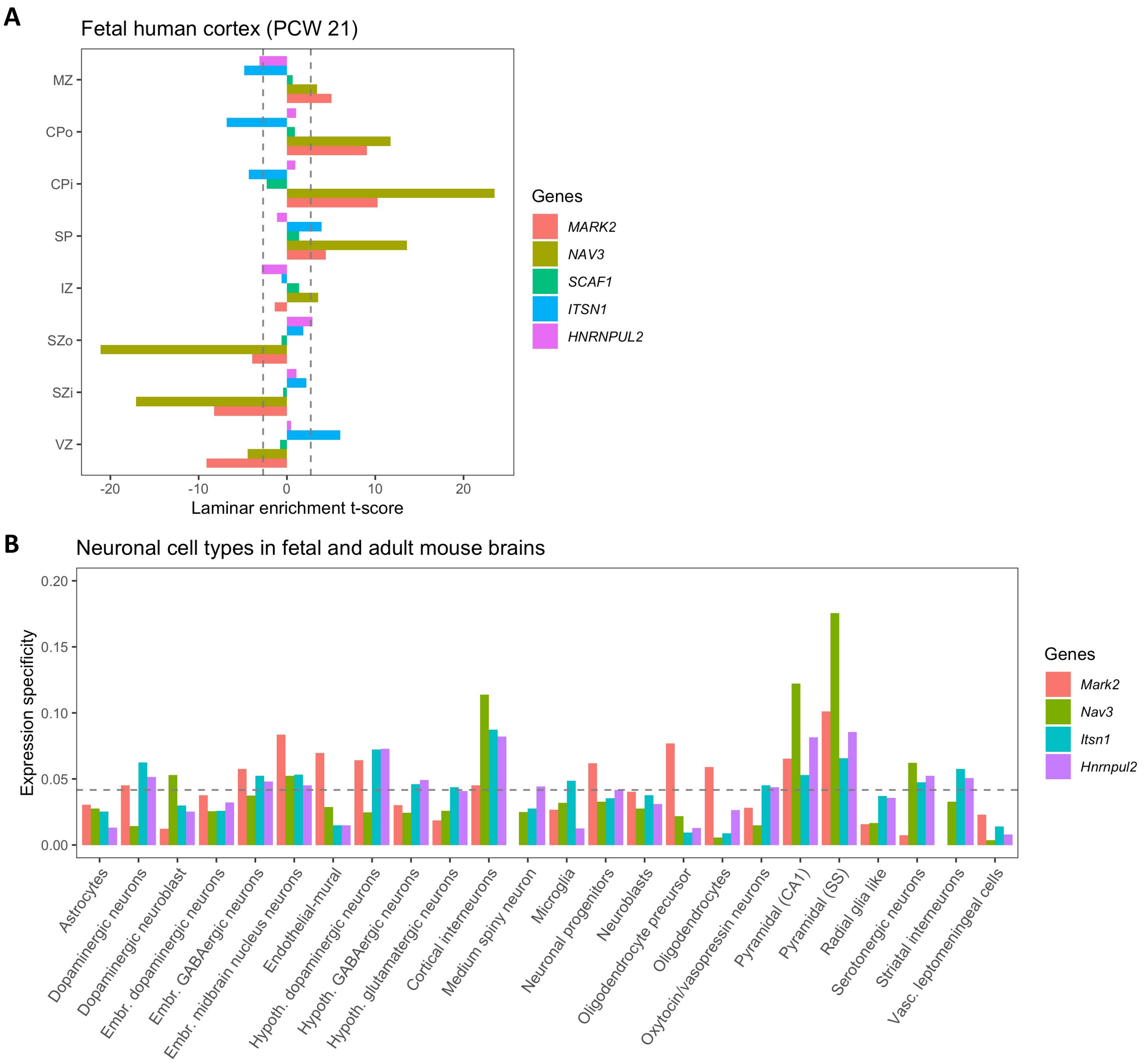


(A) Expression specificities in human fetal cortex laminar at post conceptual week (PCW) 15/16[6]. The specificity was measured by the t-statistics comparing the expression level in each layer against all other layers. Dashed lines at +/-2.7 corresponds to FDR threshold of 0.01 used in the previous study[6]. Abbreviations: marginal zone (MZ), outer/inner cortical plate (CPo/CPi), subplate (SP), intermediate zone (IZ), outer/inner subventricular zone (SZo/SZi), ventricular zone (VZ). (B) Expression specificities in neuronal cell types inferred from single cell RNA-seq data of fetal and adult mouse brains[7]. Cell type specificity was measured by mean expression level in one cell type over the summation of mean expression level across all cell types. Dashed line corresponds to uniform expression over 24 cell types. Human genes are mapped to their mouse orthologs. Single-cell expression data for SCAF1 is not available.

### Supplementary Figure S13: Transmission disequilibrium of exonic or single gene deletions of *ITSN1* and *NAV3*

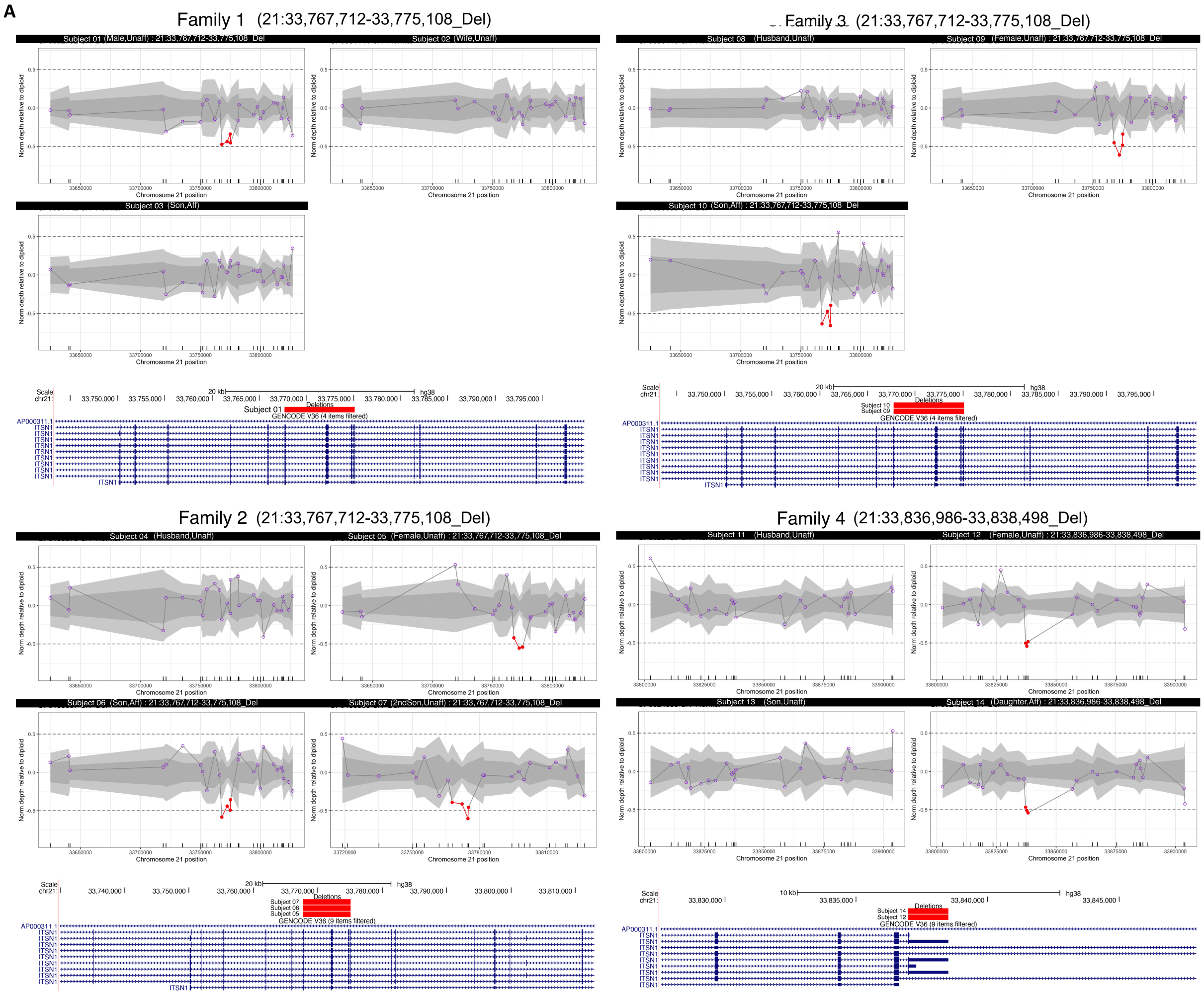

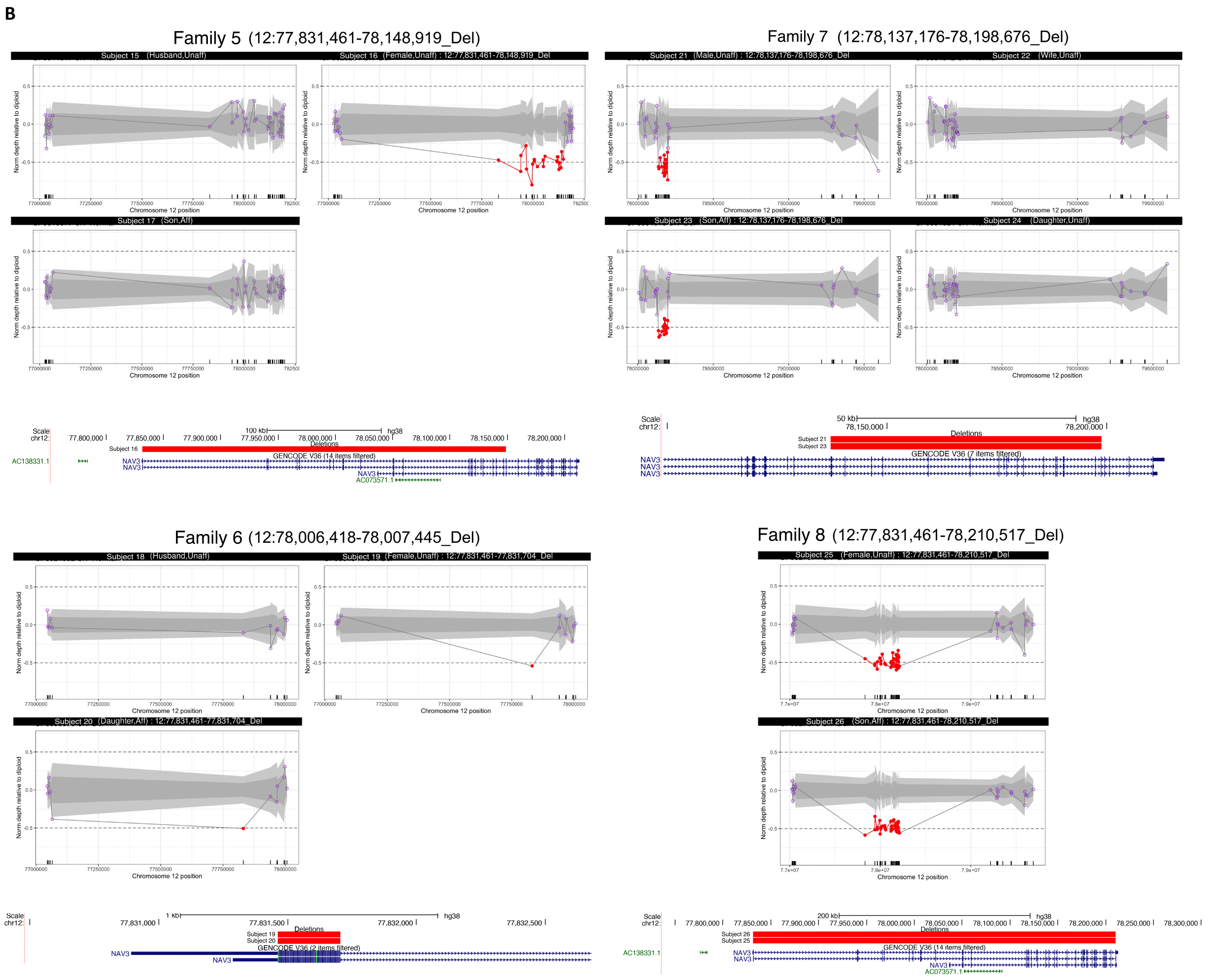


Transmission disequilibrium of exonic or single gene deletions of ITSN1 (A) and NAV3 (B). The read depth signal plots show normalized read depth (NDP) of exome targets used in CNV calling by CLAMMS93. NDPs of -0.5, 0 and 0.5 correspond to copy number (CN) of 1 (deletion), 2 (normal diploid) and 3 (duplication). NDPs of exon targets within deleted regions are colored red. Dark and gray areas correspond to 1 and 2 estimated standard deviations of NDP for each exon target. CN deletions were initially discovered from all unaffected parents, then subsequently genotyped on all family members. Signal plots of all family members are shown with parents appear in the top and offspring in the bottom. The deletion regions and affected exons of genes are shown below each plot.

### Supplementary Figure S14: Calculated cognitive impairment and sex ratio in individuals with ASD in SPARK

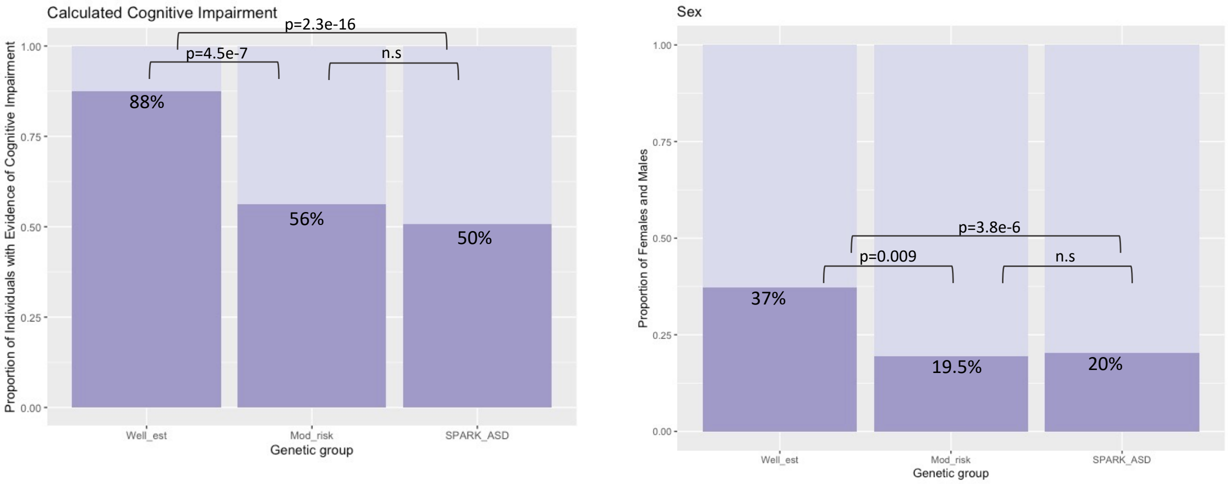


(A) The proportion of individuals with ASD with evidence of cognitive impairment is shown in dark purple and the proportion of individuals without evidence of cognitive impairment is shown in light purple. The proportion of in individuals with evidence of cognitive impairment (n=129) with HC LoF variants in well-established, highly-penetrant ASD risk genes (*CHD8, SCN2A, ADNP, FOXP1, SHANK3*) is significantly higher than 8,731 offspring with ASD in SPARK individuals (p=2.3e-16, chi-squared test), although the proportion of individuals (n=87) with LoF variants in novel, moderate ASD risk genes (*NAV3, ITSN1, SCAF1*, and *HNRNPUL2*) is not. (B) The proportion of individuals that are female is shown in dark purple and male is shown in light purple. The proportion of males to females in SPARK (n = 8,731) is 4:1 and is similar in 87 individuals with LoF variants in novel, moderate ASD risk genes. As previously reported, individuals with LoF variants in well-established ASD risk genes show an enrichment of females.

### Supplementary Figure S15: Distribution of different types of LoF variants in known ASD genes enriched by de novo variants (DNVs) and comparison with population controls

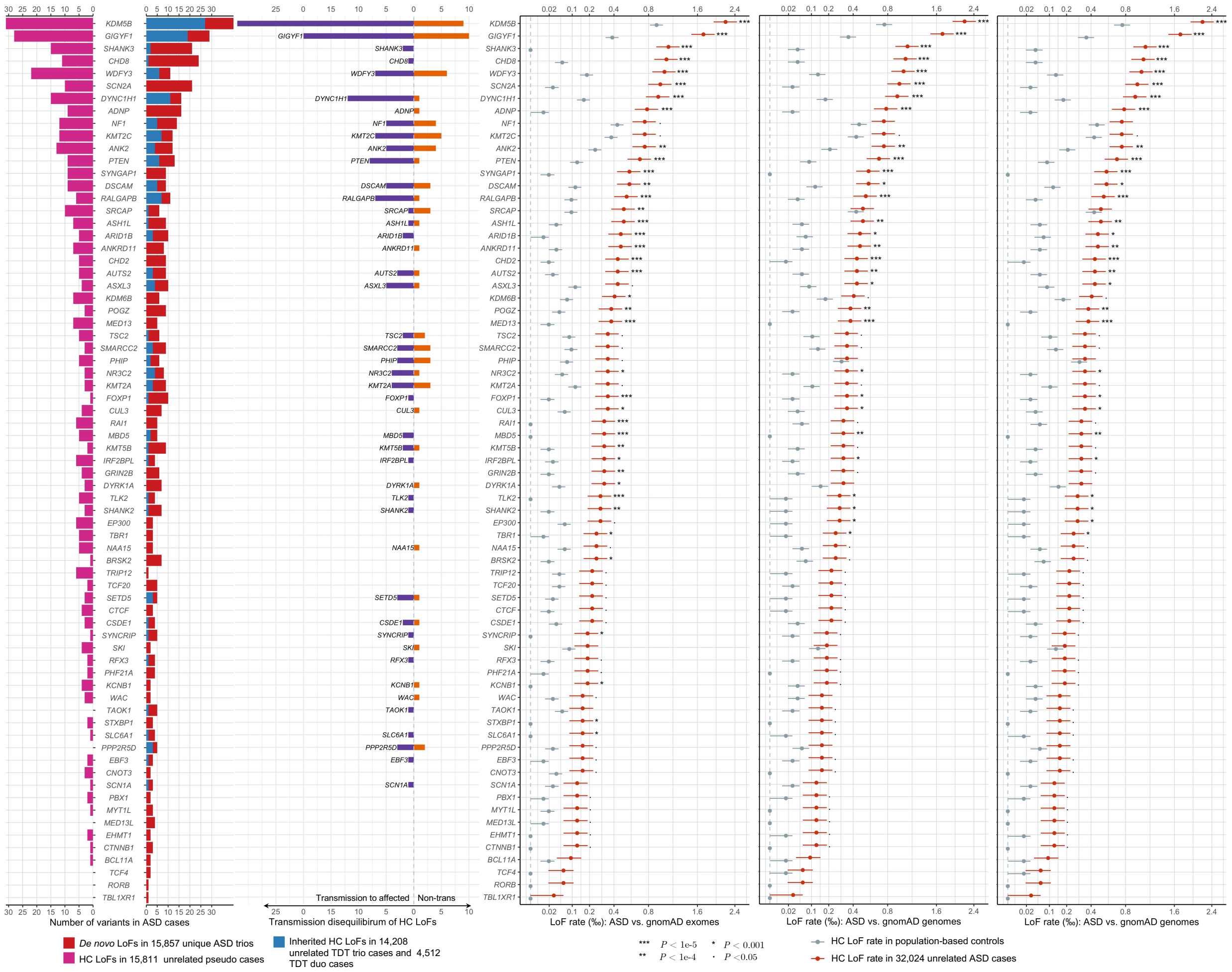


From left to right: pyramid plot summarizing the number of de novo and inherited HC LoFs in family-based samples, HC LoFs in unrelated pseudo cases; bar plot of transmission vs. non-transmission for standing HC LoFs identified in unaffected parents; and comparing HC LoF rate in mega cases with three population controls. The plot shows 71 known ASD/NDD genes that are in top 30% gnomAD LOEUF and have p-value for enrichment of all DNVs <1e-4 in 23,053 ASD trios.

#### Supplementary Figure S16: Empirical relationship between estimated relative risk to ASD and estimated selection coefficient

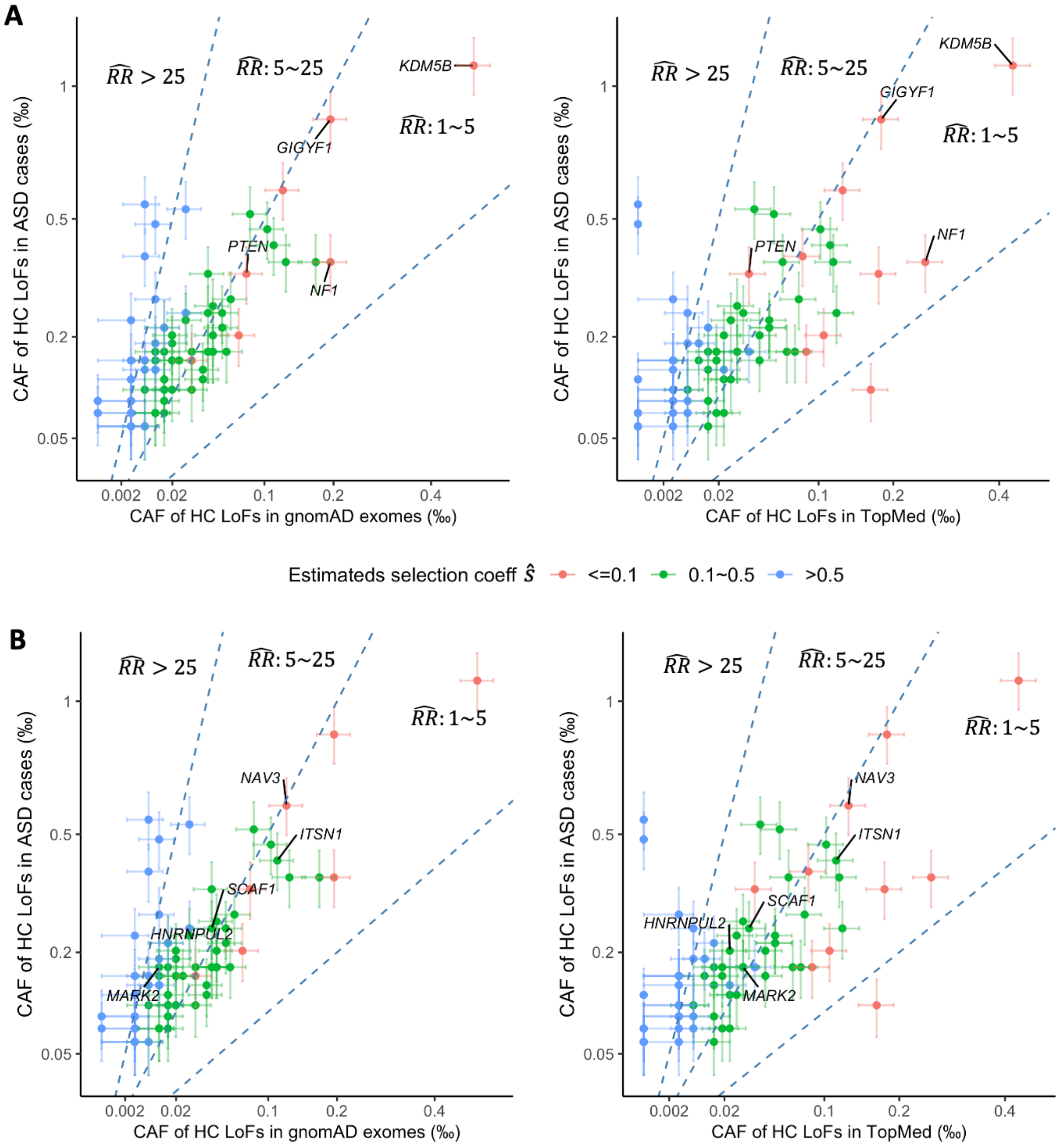

Empirical relationship between estimated relative risk ($\widehat{RR}$) to ASD and estimated selection coefficient ($\hat{s}$). We selected 66 known ASD/NDD genes from top 30% gnomAD LOEUF that have ASD DenovoWEST P-value<1e-4 and LoF mega-analysis P-value<0.05, and also included five novel ASD genes identified from the current study. Cumulative allele frequencies (CAFs) of HC LoFs were estimated from 32,024 unrelated ASD cases, and three panels of population-based controls from gnomAD and TopMed with sample size 76,000~132,000. Three dashed lines demarcate the quadrant into areas of different estimated relative risks ($\widehat{RR}$: <1, 1~5, 5~25, and >25). Genes with higher effect size to ASD (larger $\widehat{RR}$) are under stronger selective pressure (higher $\hat{s}$).

#### Supplementary Figure S17: Cumulative distribution of haploid mutation rates of LoF and D-mis variants of all protein coding genes on autosomes

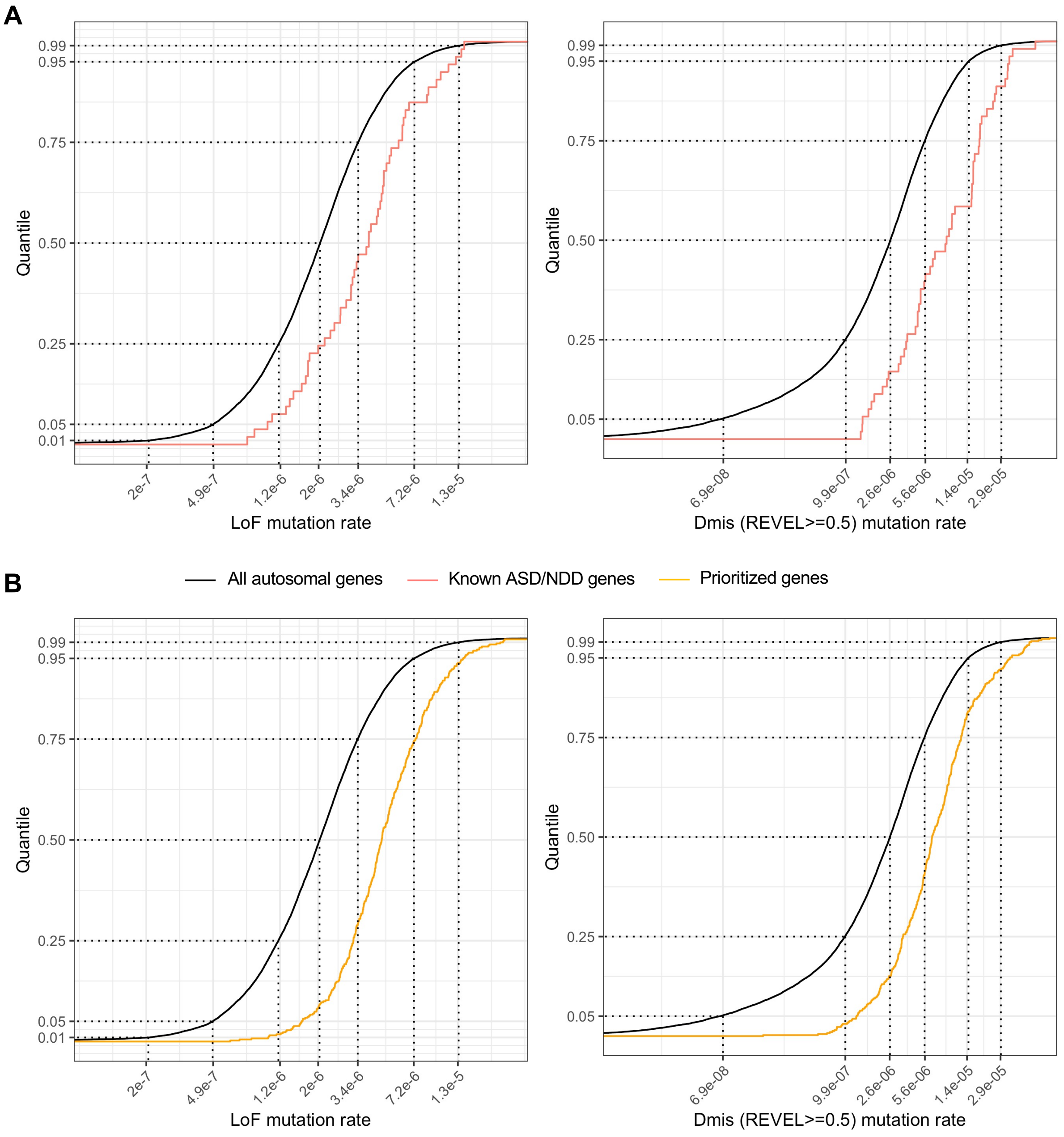

Cumulative distribution of haploid mutation rates (per generation) of LoF and D-mis (REVEL>=0.5) variants of all protein coding genes on autosomes. Baseline mutation rates were calculated using 7mer sequence context dependent mutation rates[5]. Known ASD/NDD genes tend to have higher mutation rates than average genes.

### Supplementary Figure S18: Burden of de novo and inherited LoFs in genes with high and low LoF mutation rates

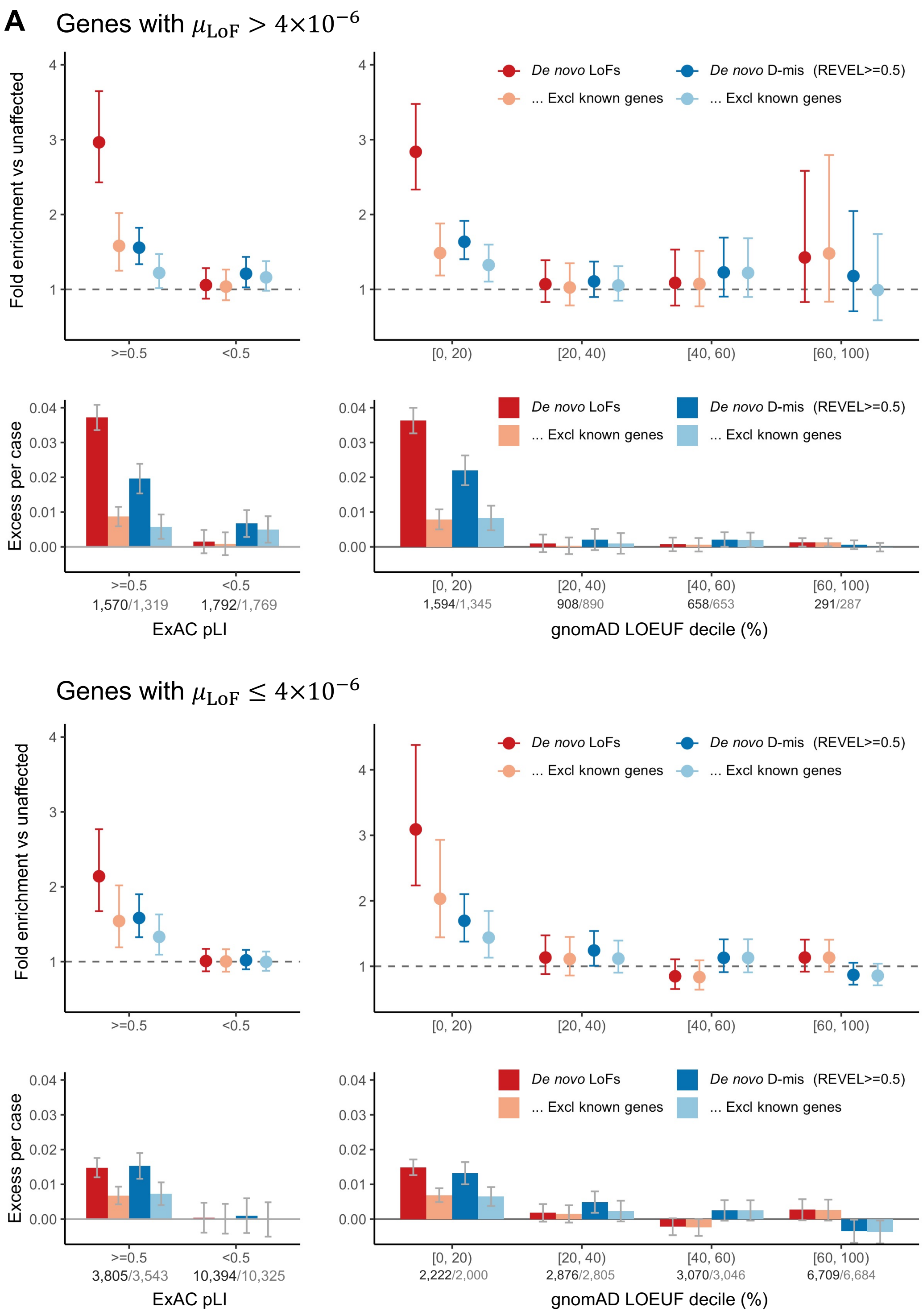

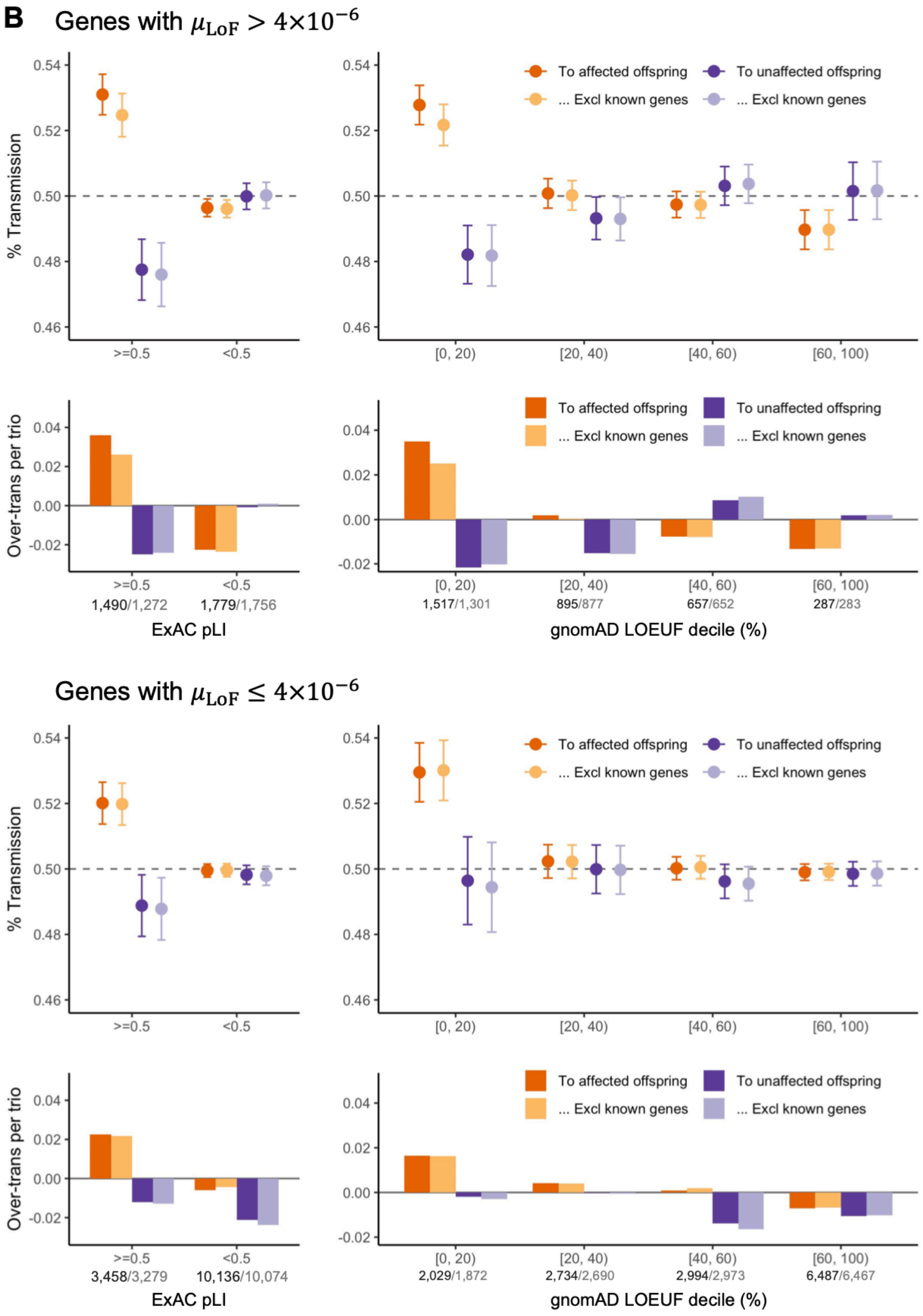
B
Genes with $\mu_{\mathrm{LoF}} > 4\times10^{-6}$
% Transmission
0.54
0.52
0.50
0.48
0.46
>=0.5
<0.5
[0, 20)
[20, 40)
[40, 60)
[60, 100)
To affected offspring
... Excl known genes
To unaffected offspring
... Excl known genes
Over-trans per trio
0.04
0.02
0.00
-0.02
1,490/1,272
1,779/1,756
1,517/1,301
895/877
657/652
287/283
ExAC pLI
gnomAD LOEUF decile (%)
Genes with $\mu_{\mathrm{LoF}} \leq 4\times10^{-6}$
3,458/3,279
10,136/10,074
2,029/1,872
2,734/2,690
2,994/2,973
6,487/6,467

(A) The burden of de novo variants among genes with high mutation rate (upper panel) and low mutation rate (lower panel) was evaluated by rate ratio and rate difference between 16,877 ASD and 5,764 unaffected trios. Exome-wide burdens of de novo LoF and Dmis (REVEL>=0.5) variants concentrate in constrained genes (ExAC pLI>=0.5) and in genes with highest level of LoF-intolerance in the population defined by top two deciles of gnomAD LOEUF scores. The number of genes before and after removing known genes in each constraint bin was shown below the axis label. (B) Burden of inherited LoFs with high mutation rate (upper panel) and low mutation rate (lower panel) was evaluated by looking at the proportion of rare LoFs in 20,491 unaffected parents that are transmitted to affected offspring in 9,504 trios and 2,966 duos and can be quantified as over-transmission of LoFs per ASD trio. As a comparison, we also show the transmission disequilibrium pattern to unaffected offspring in 5,110 trios and 129 duos. Using ultra-rare LoFs with pExt>=0.1, exome-wide signals of transmission disequilibrium of standing LoF variants also concentrate in constrained genes (ExAC pLI>=0.5) and in genes at top two decile of gnomAD LOEUF scores. Analysis was restricted to autosomal genes and repeated after removing known ASD/NDD genes (number of genes in each constrained bin before and after removing known genes is shown below the axis label).

#### Supplementary Figure S19: Power of case-control association by rare LoFs variants with sample size equal to current study

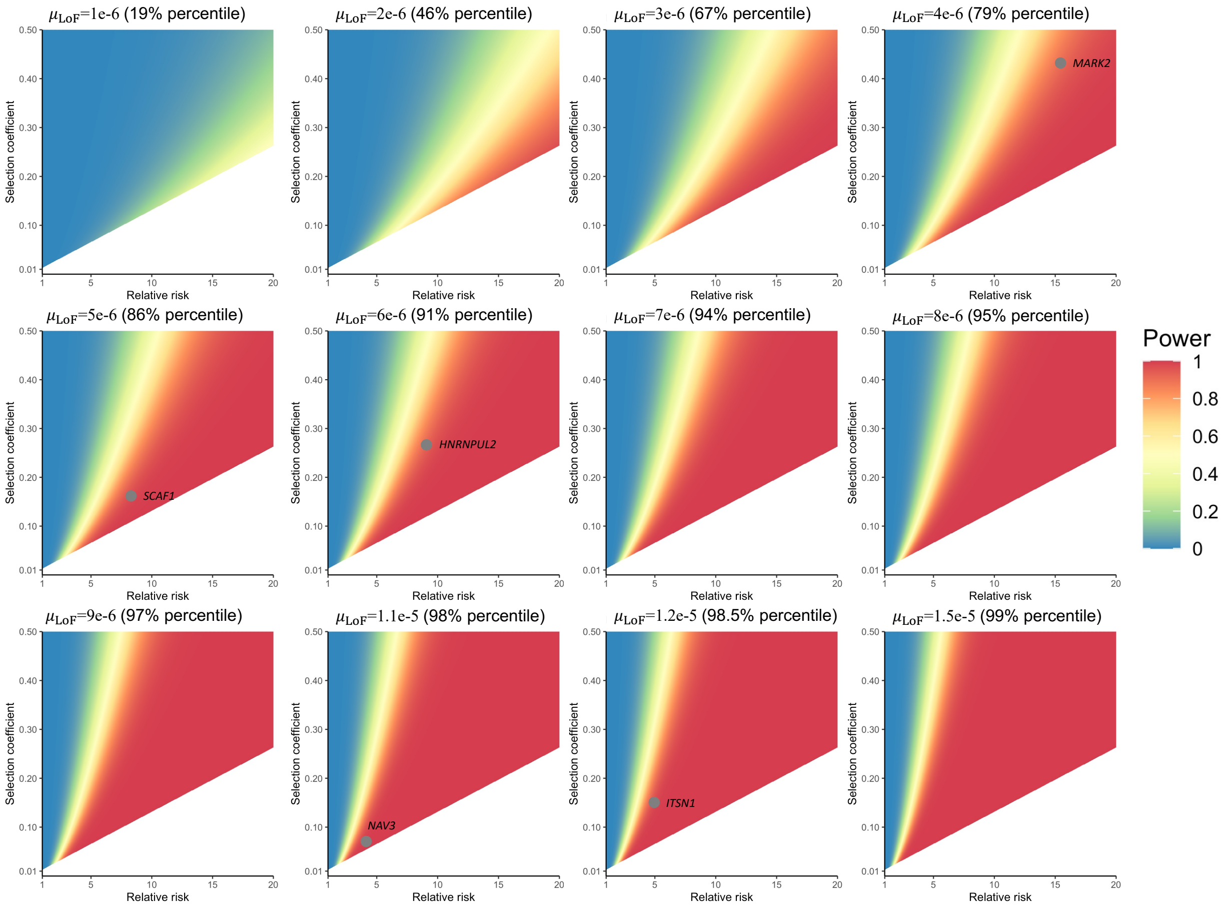


Power of case-control association by rare LoFs variants ("mega-analysis") with sample size equal to current study. The mega-analysis of current study compares the rate of LoF variants in 32,024 unrelated ASD cases with population controls with sample sizes about 76,000~132,000. For power calculation, we assumed that population controls are infinite so that cumulative allele frequency are known and presumed to be at equilibrium under selection-mutation balance for constrained genes ($f = \mu_{LoF}/s$). Experiment-wide error rate was set at 9e-6 (0.05 divided by the number of autosomal genes at gnomAD LOEUF 30%). Power is calculated as a function of relative risk for ASD ($RR$) and selection coefficient ($s$) across different haploid LoF mutation rates ($\mu_{LoF}$) using an analytic approximation by Zuk *et al.* 2014[8]. We only considered selection coefficient between 0.01 and 0.5 and relative risk to ASD between 1 and 20, because genes with huge effect sizes and larger selection coefficients are expected to be identified from the enrichment of *de novo* variants. The triangular region where $s < 0.013RR$ are left blank because the parameters in this region are not compatible with the current estimates of prevalence of ASD (1/54)[9] and sex-averaged reduction of reproductive fitness (0.71)[10]. Five new ASD genes identified in this study are placed onto the heatmap closest to its LoF mutation rate. Their positions within heatmaps are taken from point estimates of using gnomAD exomes (non-neuro subset) as population controls

#### Supplementary Figure S20: Sample sizes required for achieving 90% of power

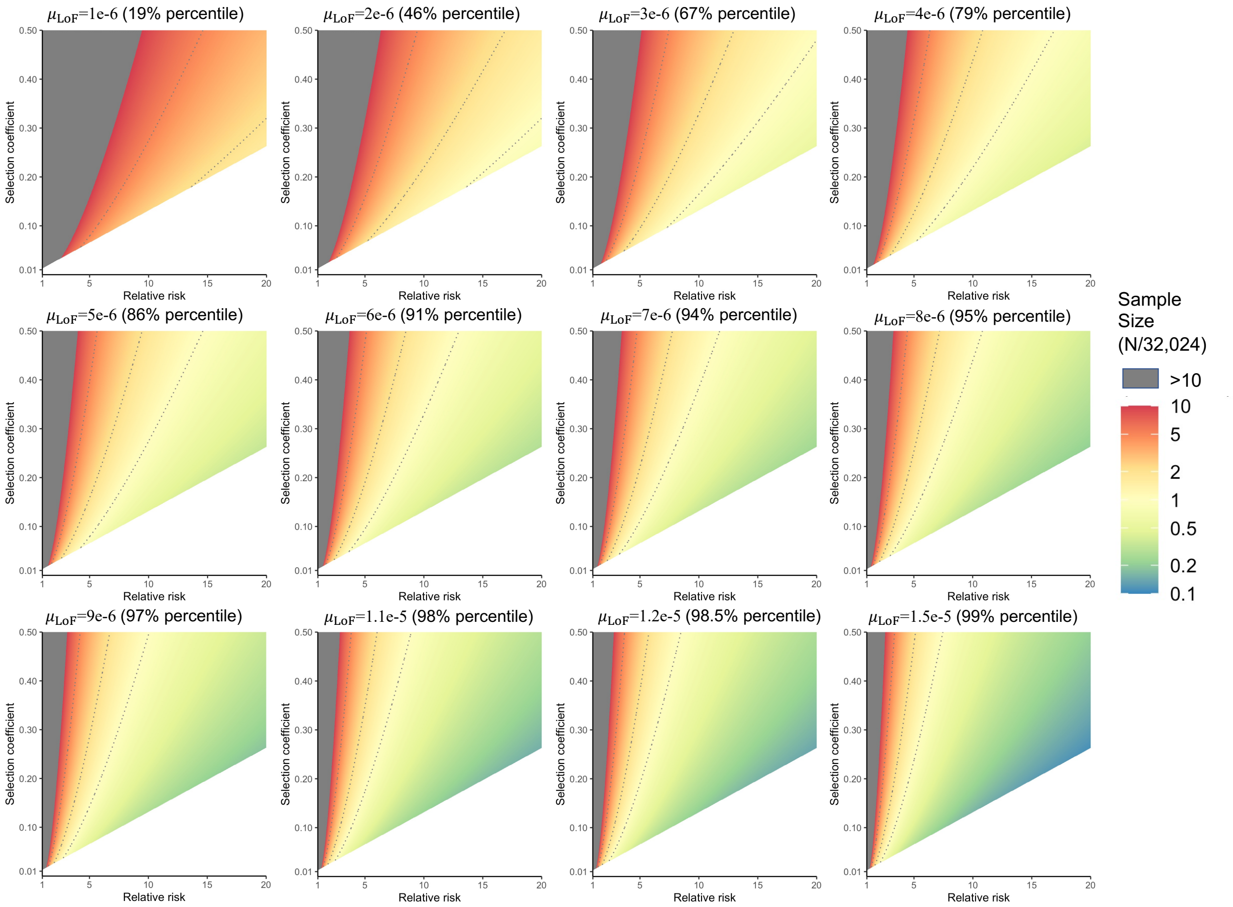


Using the same assumptions and experiment-wide error rate as power calculation, we calculated required sample size for 90% of power. Sample size is shown as a factor relative to the current sample size (32,024) and as a function of relative risk to ASD and selection coefficient across with different LoF mutation rates. Contours 1, 2 and 5 times of current size are shown as dashed lines. Regions of parameter space that require over 10 times current sample sizes are shown in gray.

### Supplementary Figure S21: SPARK sample QC: relatedness check and sex validation

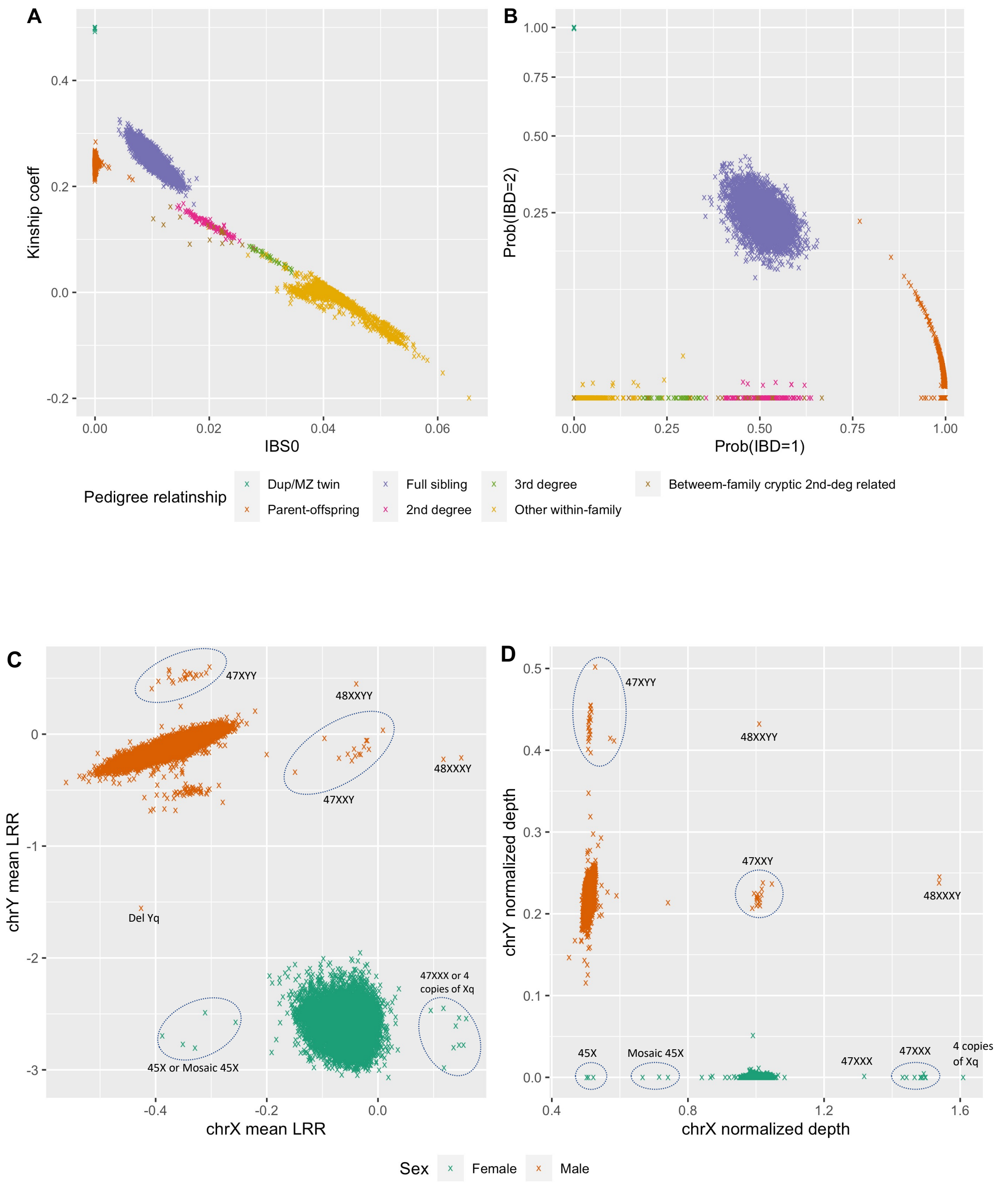

(A-B) Known pedigree relationships are verified by estimated kinship coefficients, proportion of SNPs with zero alleles shared identity by state (IBS0), and probabilities of 1 or 2 copies of chromosomes shared identity by descent (Prob(IBD=1) and Prob(IBD=2)). (C-D) Sample sexes are verified by log-R ratio (LRR) signals and normalized read depth of sex chromosomes. Samples with sex chromosome aneuploidies are highlighted.

### Supplementary Figure S22: SPARK principal component analysis (PCA) and ancestry inference

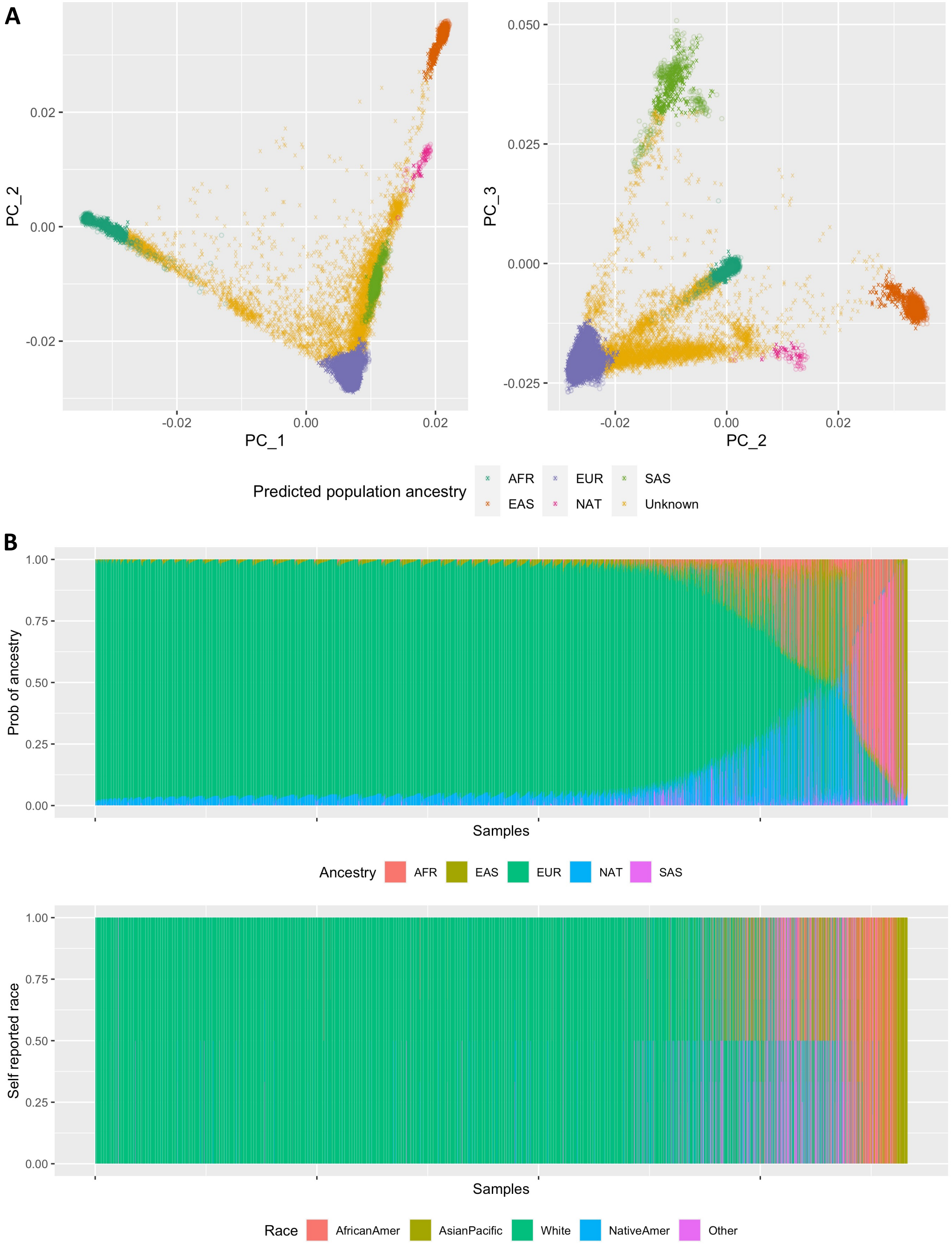

(A) PCA was first performed on samples from five reference populations including 650 Africans (AFR), 504 east Asians (EAS), 503 Europeans (EUR) and 486 south Asians (SAS) from 1000 Genomes Project and 48 native Americans (NAT) from HGDP-CEPH panel. SNP genotypes of 28,649 SPARK samples in the discovery cohort were then projected onto the principal axes of defined by the reference populations. The top three principal axes are shown: circle points are samples from reference populations, cross points are SPARK samples. The projected coordinates at top four axes are transformed to the probabilities of population ancestries using SNPweights method25. Sample is predicted to originate from one reference population if the corresponding probability >=0.8. Samples whose origin cannot be classified by the above criteria are labeled as unknown. (B) Comparing self-reported race(s) and inferred probabilities of population ancestries. For 7,176 offspring cases, self-reported race(s) are available, which include one or more of the following: black or African American (AfricanAmer), Asian or Native Hawaiian or other Pacific Islander (AsianPacific), White, American Indian or Alaska Native (NativeAmer), and Other. Inferred probability of five reference populations for each individual sum up to 1 and are visualized using a stacked bar plot. Individuals are ordered by the probability of EUR, AFR, ASI and NAT. Self-reported races for the same set of individuals are also visualized below as a bar plot. For individuals with multiple self-reported races, multiple races are shown as sub-bars with equal length. There is a general concordance of individuals with self-reported white and Asian with predicted EUR and ASI ancestries. Individuals with self-reported African American or native American are more likely to have recent admixture of EUR, AFR and NAT.

#### Supplementary Figure S23: SPARK self-reported cognitive impairment shows stronger correlation with Vineland score than full-scale IQ

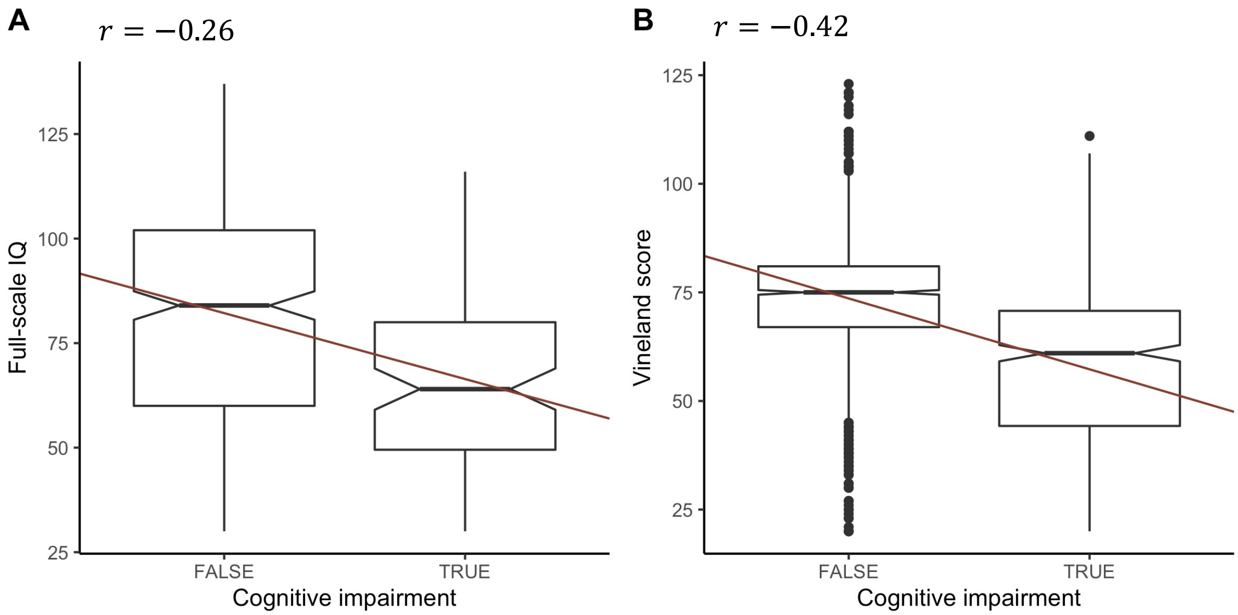


(A) In 478 samples with full-scale IQ, Pearson correlation between IQ and cognitive impairment is -0.26. (B) In 2183 samples with standardized Vineland score, correlation between Vineland scores and cognitive impairment is -0.42.

#### Supplementary Figure S24: Comparing phenotypes of samples from simplex and multiplex families in SPARK cohort

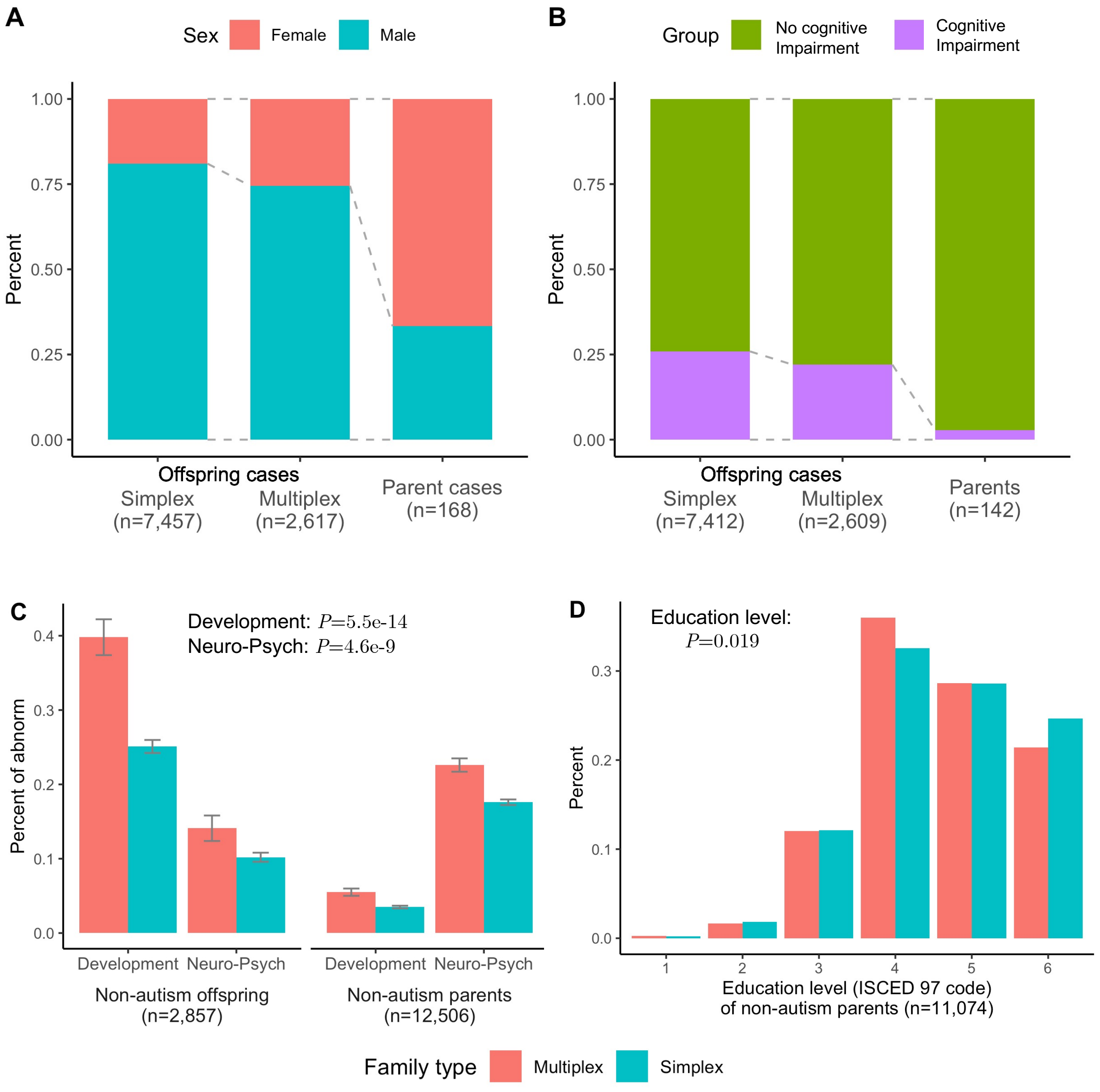


Multiplex families are defined as families with at least one pair of affected first degree relatives at recruitment or by self-report, all other families are simplex. (A) There is more female ASD cases in multiplex families, especially among affected parents. (B) Parent cases are also less likely to have cognitive impairment. Sample sizes shown below each group are the number of samples with non-missing information. (C) Unaffected family members in multiplex families are more likely to have other developmental or neuro-psychiatric issues than simplex families. Developmental issues include structural birth defects, learning or language disability, motor delays, social communication problems, etc. Neuro-psychiatric issues include seizure, schizophrenia or schizoaffective disorder, bipolar disorder, Tourette syndrome, etc. The association with family history was evaluated by logistic regression adjusting sex and role in the family. (D) Unaffected parents in multiplex families also have lower education attainment than unaffected parents in simplex families (P=0.019, by linear regression adjusting sex). Education level were coded by International Standard Classification of Education (1997).

### Supplementary Figure S25: Summary of final DNV call sets for SPARK and SSC discovery samples

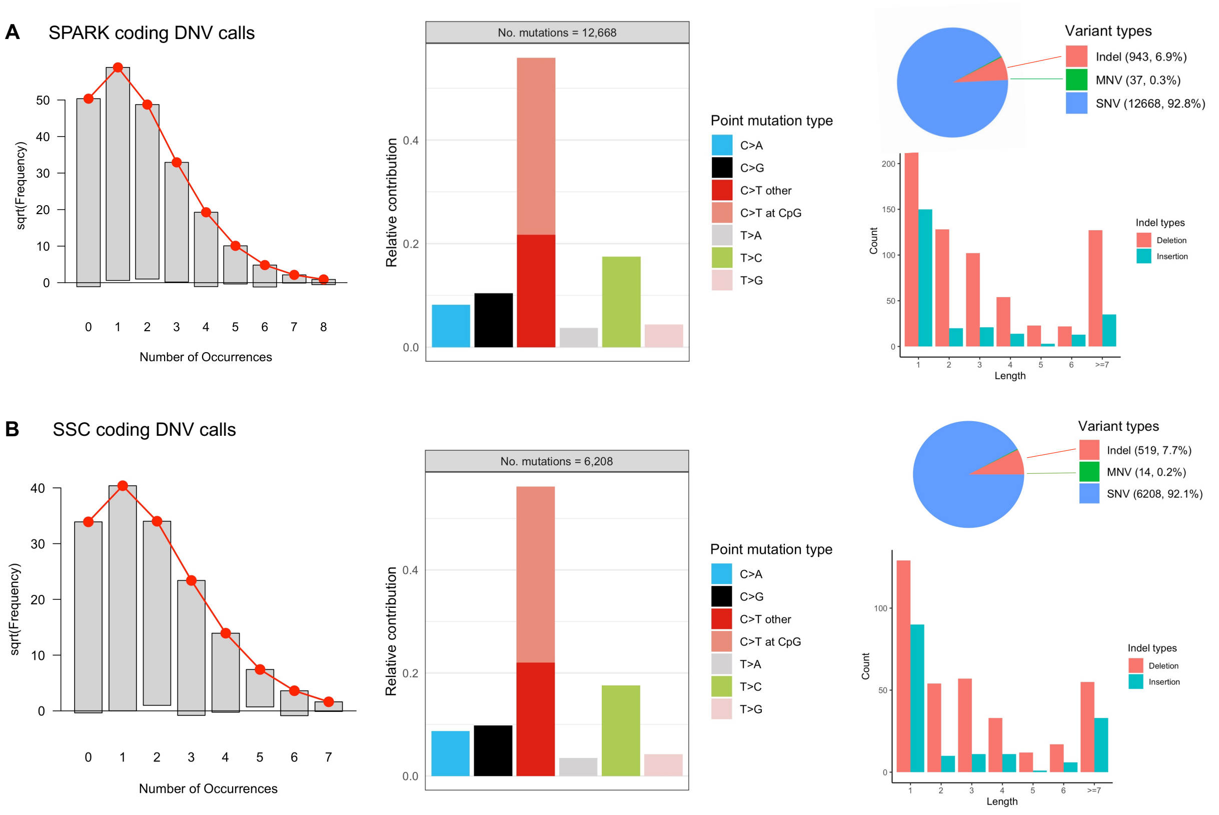


From left to right: identified coding DNVs per trio and fitted Poisson distribution, SNV mutation spectrum, indel length distribution and relative proportion of indels and SNVs.

### Supplementary Figure S26: Evidence of post-zygotic mosaicisms in the final DNV call set

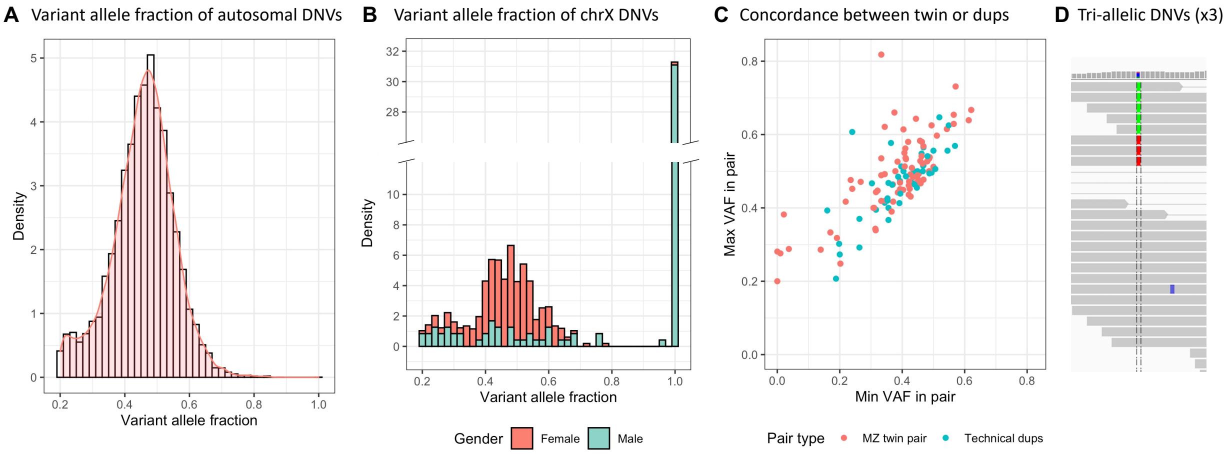


(A) Distribution of variant allele fractions (VAF) of autosomal DNVs shows a small peak at low VAF end. (B) Heterozygous non-PAR chrX DNVs were identified in males. (C) For samples with technical duplicate or MZ twin, VAFs are highly correlated between duplicate or twin pairs. But a small number DNVs with VAF between 0.2 and 0.4 were detected only in one of twin pairs. (D) A small number of DNVs are tri-allelic due to a second post-zygotic mutation.

### Supplementary Figure S27: Rare variant workflow and QC strategy

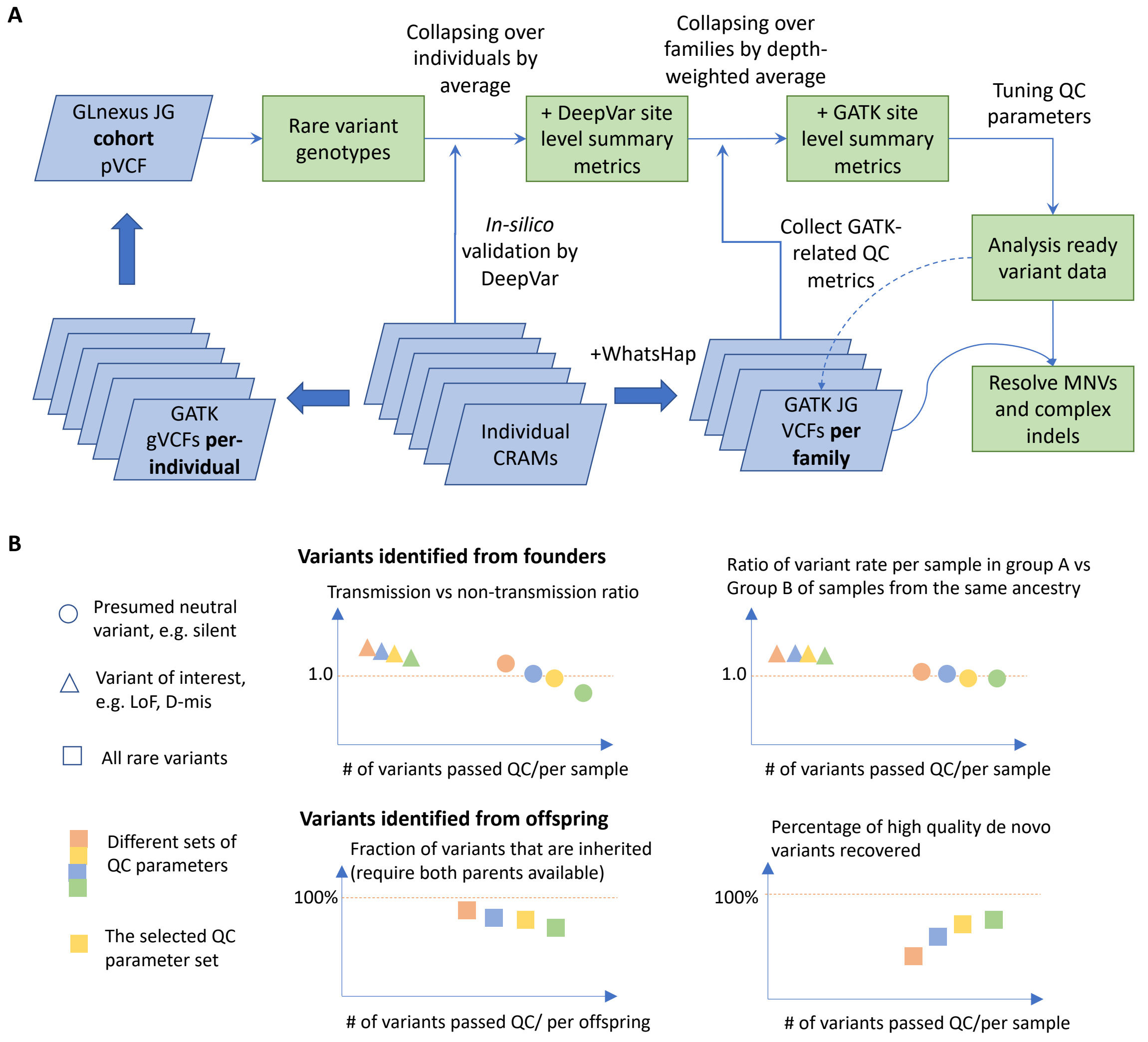


Details are described in Methods section.

### Supplementary Figure S28: Comparison of inhouse DenovoWEST results on NDD trios with published results

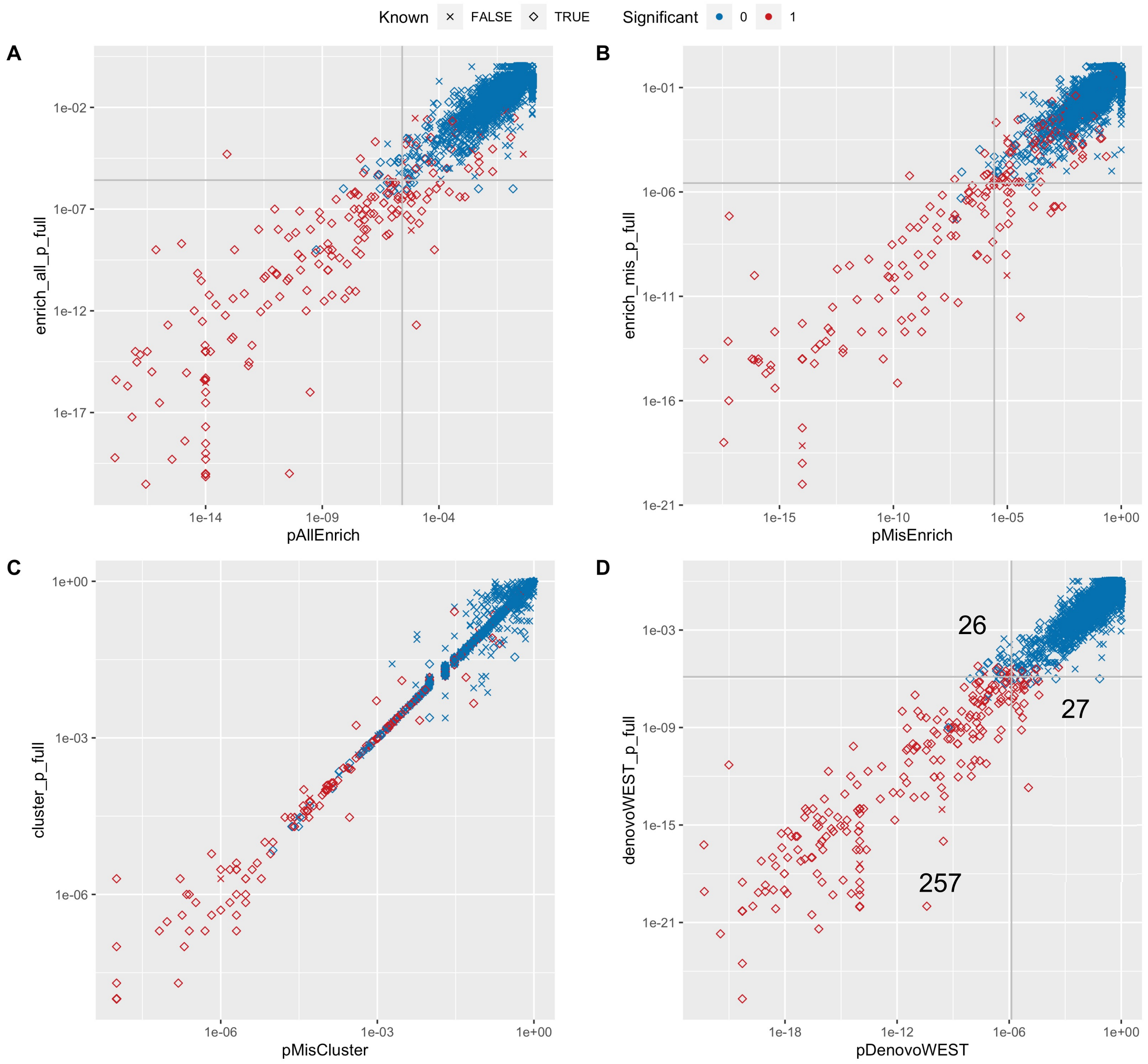


We reannotated DNVs of 31,058 NDD trios from the previous study2 and tested enrichment of DNVs in each coding gene by inhouse implementation of DenovoWEST. For each gene, four different p-values were generated and compared: (A) pAllEnrich: one-sided p-value for enrichment of all DNVs; (B) pMisEnrich: one-sided p-value for enrichment of missense DNVs; (C) pMisComb: combined p-value for enrichment and clustering of missense DNVs; (D) pDenovowEST: the final DenovoWEST p-value as the minimum of pAllEnrich and pMisComb. P-values from the reanalysis are shown on X-axes and compared with the published p-values on y-axes. Known developmental disease genes included in DDG2P58 (2020-02) are shown in diamond shape, and genes declared exome-wide significant in the previous study was highlighted in red.

#### Supplementary Figure S29: Illustration of pseudo cases and contributing sample sizes in different types of pedigrees

**Nuclear families**

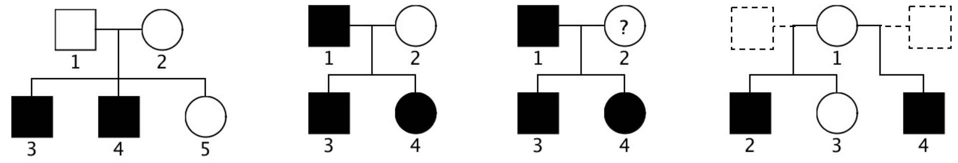


| | | | | |
|---|---|---|---|---|
| **De novo** | Two affected trios (3,4) | Two affected trios (3,4) | Two affected trios (3,4) | 0 |
| **TDT** | Two affected trios (3, 4) | Two affected duos (3,4) | 0: neither parent is unaffected | Two affected duo cases (2,4). |
| **Pseudo cases** | 0 | One affected parent (1) and 2 half affected offspring (3,4; after removing maternal alleles and de novo variants) | One affected parent (1), two full affected offspring (3,4). | Two half affected offspring (2,4; after removing maternal alleles) |

**Multi-generation families**

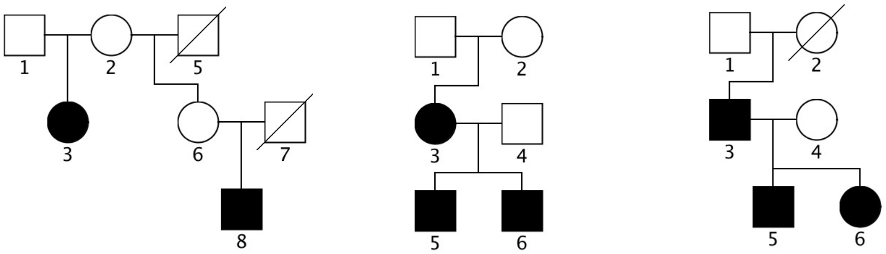


| | | | |
|---|---|---|---|
| **De novo** | Two affected trios (3,8) | Three affected trios (3,5,6) | One affected trio (5) |
| **TDT** | One affected trio (3), one affected duo (8) | One affected trio (3), two affected duos (5,6) | Three affected duos (3,5,6) |
| **Pseudo cases** | One half affected offspring (8; after removing maternal alleles) | 0: For two TDT duos (5,6), alleles not transmitted from unaffected father (4) have been analyzed in affected mother (3) and in their de novo variants. | Two ¼ affected offspring (5,6; after removing maternal alleles, de novo variants, and grand-parent alleles) |

Pseudo cases in family-based samples were created from cases to include variant genotypes that were not used by de novo or TDT analysis. Algorithms used create pseudo cases and determine their contributing sample sizes are described in Methods section.
